# Estimating COVID-19 Vaccine Protection Rates via Dynamic Epidemiological Models–A Study of Ten Countries

**DOI:** 10.1101/2022.08.08.22278571

**Authors:** Yuru Zhu, Jia Gu, Yumou Qiu, Song Xi Chen

## Abstract

The real-world performance of vaccines against COVID-19 infections is critically important to counter the pandemics. We propose a varying coefficient stochastic epidemic model to estimate the vaccine protection rates based on the publicly available epidemiological and vaccination data. To tackle the challenges posed by the unobserved state variables, we develop a multi-step decentralized estimation procedure that uses different data segments to estimate different parameters. A B-spline structure is used to approximate the underlying infection rates and to facilitate model simulation in obtaining an objective function between the imputed and the simulation-based estimates of the latent state variables, leading to simulation-based estimation of the diagnosis rate using data in the pre-vaccine period and the vaccine effect parameters using data in the post-vaccine periods. And the time-varying infection, recovery and death rates are estimated by kernel regressions. We apply the proposed method to analyze the data in ten countries which collectively used 8 vaccines. The analysis reveals that the average protection rate of the full vaccination was at least 22% higher than that of the partial vaccination and was largely above the WHO recognized level of 50% before November 20, 2021, including the Delta variant dominated period. The protection rates for the booster vaccine in the Omicron period were also provided.

## 1. Introduction

The COVID-19 pandemic has been raging around the globe for more than two years, which has caused waves of infections and deaths among countries. The pandemic has prompted the development of vaccines which have been clinically administrated in various countries since early 2021. Ten COVID-19 vaccines had been approved for public use by the World Health Organization (WHO) as of January 2022. Vaccine makers have provided clinical trial results on the efficacy of their products. The vaccine efficacy against the original SARS-Cov-2 strain from recent studies is reported in Table S1, which ranged from 50.7% to 95% for the two-dose vaccination. However, SARS-CoV-2 has undergone progressive changes. The Delta variant has caused global pandemics with a high transmission rate (Planas et al., 2021), resulting in considerable socioeconomic burden and pressure on hospital systems (Liu et al., 2021).

There is a great need to evaluate the vaccine effectiveness in the real-world situation. Clinical trials used to evaluate the vaccine efficacy exclude certain part of the population and are difficult to control all confounding factors that may cause infections in the population. The retrospective studies, such as Li et al. (2021), used the proportion of the vaccine breakthrough cases in all infected cases in specific institutions, which typically had a small sample size relative to the daily infections in a country. Moreover, most countries do not collect vaccine breakthrough statistics, which brings great challenges for evaluating vaccine effects in real-world situations. In addition, many studies have only provided data on effectiveness against symptomatic cases as including asymptomatic ones is difficult. Hence, the real-world performance against any infection is an important issue. In this study, we use the real-world vaccine protection rate (VPR), defined as one minus the percentage reduction in the infection rate of the vaccinated relative to the population without vaccine protection (unvaccinated and vaccine-expired) of a country, to measure the combined effectiveness of vaccines administrated in a country against SARS-Cov-2 infection at the population level.

Compartmental models, like SIR (Kermack and McKendrick, 1927) and SEIR (Anderson and May, 1982) models, are widely used for modeling the transmission of infectious diseases, which divide a population into compartments and specify the transition rates among compartments by ordinary differential equations (ODEs). Stochastic epidemic models (SEMs) extended from the SEIR model had been proposed to study the spread of COVID-19 before vaccines were available (Hao et al., 2020; Tian et al., 2021; Yan et al., 2021). As asymptomatic and pre-symptomatic infections in the COVID-19 pandemic are not observable, the likelihood functions based on the observed data are rather complex due to having to integrate out unobserved state variables. Bayesian methods which target the joint posterior distribution of the unobserved data and the model parameters use the Markov chain Monte Carlo (Auranen et al., 2000) or the sequential Monte Carlo (Dukic, Lopes and Polson, 2012) for parameter estimation. Quick, Dey and Lin (2021) built a multilevel regression model on the COVID-19 daily confirmed infection counts, polymerase chain reaction testing and serological survey data, and developed an EM algorithm to estimate the model parameters.

There are recent studies (Dashtbali and Mirzaie, 2021; Giordano et al., 2021) in evaluating COVID-19 vaccination strategies using a deterministic compartment model that assumes a portion of the vaccinated can achieve permanent and full immunity, which is quite restrictive. Moreover, deterministic models are inadequate to facilitate statistical inference on the characteristics of epidemics. Incorporating stochasticity into epidemic models is needed for real-world evaluation due to the randomness of the state variables. There has been no study using the publicly available data to estimate the real-world COVID-19 VPR without the restrictive permanent and full immunity assumption and incorporating the stochastic natures of the epidemics and the unobservable asymptomatic and pre-symptomatic cases.

We propose a new SEM for evaluating COVID-19 VPR based on publicly available epidemiological and vaccination data, which allows breakthroughs in fully and partially immunized people, and infection before clinical diagnosis and being asymptomatic. Its advantages over the traditional cohort or case-control studies are in its much-reduced data collection cost and timely assessment on the real-world performance of vaccines.

The unavailability of data on infections before clinical confirmation and vaccine breakthrough brings several challenges to estimating real-world vaccine effects. First, unobservable compartments in the model and the complexity of the model make the maximum likelihood estimation and the EM algorithm, which involves integrating out the latent variables in the conditional distribution, difficult to be implemented. Second, the approach of the leaveone-out cross-validation criteria with a kernel smoothing procedure for estimating parameters of the varying coefficient SEM built for modeling the pre-vaccine COVID epidemic in Yan et al. (2021) is no longer applicable with the vaccine-related compartments, since the model is more complex with more unobservable compartments. Third, although the conditional means of our model can be specified via the ODEs, the estimation methods used for the deterministic ODEs from partially observed data or data with additive measurement errors are not applicable for the proposed SEM with heterogeneity due to the conditional Poisson specification. The methods include the simulation-based estimation approach such as the single (Hicks and Ray, 1971) and multiple (Baake et al., 1992) shooting methods which minimize certain distance measures between the observed state variables and the simulated trajectories of the variables via the ODEs given the parameter values. The non-simulation-based estimation approaches for differential-equation-based models involved the generalized profiling estimation via B-splines expansions or the nonparametric regression estimate of model coefficients (Ramsay et al., 2007; Liang and Wu, 2008).

To tackle these challenges, this study proposes a decentralized estimation approach that utilizes different periods of the data series for estimating different parameters, taking advantage of long observations of the COVID-19 epidemics. Unobservable compartments are imputed by data on observed compartments based on the proposed stochastic model. A major part of the estimation is attained by minimizing certain contrast functions between the imputed and the simulation-based estimates for the infected to estimate (i) the diagnosis rate using the pre-vaccine period data, and (ii) the vaccine effect parameters using the post-vaccine period data. The final time-varying infection rate is estimated by the kernel smoothing method based on the model-implied imputation equations.

We apply the proposed model and estimation approach to analyze the data from ten countries to estimate the real-world VPRs and other key parameters. It is found that the VPRs of partial (one dose) vaccination ranged from 48% to 64% and 17.5% to 48% in the pre-Delta and the Delta-dominated periods, respectively. The VPRs of the full (usually two-dose) vaccination ranged from 68% to 95% in the pre-Delta era, which were reduced to 45% to 74% when the Delta variant dominated. The average VPRs from the full vaccination were at least 22% more than those of the partial vaccination, suggesting significant extra protection offered by the full vaccination. Furthermore, VPRs of full vaccination for the 10 countries up to November 20, 2021 (before the Omicron era) were largely above the WHO recognized 50% level with all 8 brands of the vaccines, including inactivated vaccines. Our results of the mixed effectiveness of vaccines being administrated simultaneously in a country were consistent with those of published studies via clinical trials and the retrospective studies reviewed in the supplementary material (SM).

The paper is organized as follows. Section 2 introduces the data and the periods with respect to vaccination and the pandemic. Section 3 presents the proposed SEM with vaccination compartments. The multi-step decentralized estimation procedure is given in Section 4. Section 5 reports simulation results to evaluate the proposed method. Sections 6, 7 and 8 provide the empirical analysis on the estimated vaccine effects in different periods of the pandemic, the sensitivity analysis of the proposed method, and the scenario analysis results for the no-vaccine, partial-vaccination and first-dose-priority scenarios, respectively. Section 9 extends the proposed approach to estimate the VPRs of booster vaccines and VPRs in the Omicron period. Section 10 offers a general discussion.

## 2. Data

Our analysis used the publicly available epidemiological data from February 23, 2020 to November 20, 2021 of 10 countries listed in Table S3 of the SM. The first date marked the start of the local transmission in these countries, while the second date represented the end of the pre-Omicron era as Omicron was first reported on November 24, 2021. We also extended the analysis to March 15, 2022 to cover the Omicron period. The daily epidemiological statistics were obtained from Johns Hopkins University Center for Systems Science and Engineering (JHU CSSE) COVID-19 Dashboard (Dong, Du and Gardner, 2020), and the information on the vaccine types and the cumulative numbers of people having received the partial and full vaccination was obtained from the official statistics of the countries. In this paper, the partial (full) vaccination means having not completed (having completed) the primary series of vaccination according to the definition of primary series and the prescribed number of vaccine doses of each vaccine product as specified in CDC (2022).

Some countries (Canada, Italy, Portugal, the UK, the US) under-reported their daily recovery cases almost from the beginning of the pandemics (Yan et al., 2021) as reflected in Figure S2 of the SM. Moreover, recoveries in all the 10 countries have not been reported since August, 2021. Thus, we used 14 days as the average time of recovery, as suggested by WHO and supported clinically by Guan et al. (2020), to impute the recovered cases for these countries. To reduce potential measurement errors, the daily data were smoothed using the procedure outlined in the SM with a bandwidth of 15 days.

For each country, we studied four non-overlapping periods: the pre-vaccine period from the start of the epidemic till the start of vaccination, the pre-Delta period till the Delta variant was first detected in the country, the intervening period till the Delta variant became predominant and the Delta-dominated period when the majority of the cases were caused by the Delta variant. India did not have the pre-Delta period as its first Delta case was reported before vaccination. The dates of the periods in the countries are provided in the SM.

## 3. Epidemiological Model

We propose a new SEM called varying coefficient susceptible-vaccinated-infected-diagnosed-removed (vSVIADR) model with ten compartments for a well-mixed population of size *M*. The proposed model with its compartments and key parameters is illustrated in Figure 1. It shows that in addition to the latent asymptomatic and pre-symptomatic compartments before diagnosis considered in Yan et al. (2021), we add the partially and fully vaccine immunized compartments to model the transmission after vaccination. This allows to estimate the VPR in real-world situations using daily statistics of epidemics and vaccination. As the vaccine supply and distribution vary from country to country, such an estimation would better reflect the actual vaccine effect in a given population than the estimates from the experimental studies using clinical trial data (Voysey et al., 2021; Polack et al., 2020) or the retrospective studies (Sheikh et al., 2021; Li et al., 2021).

**Fig 1:**
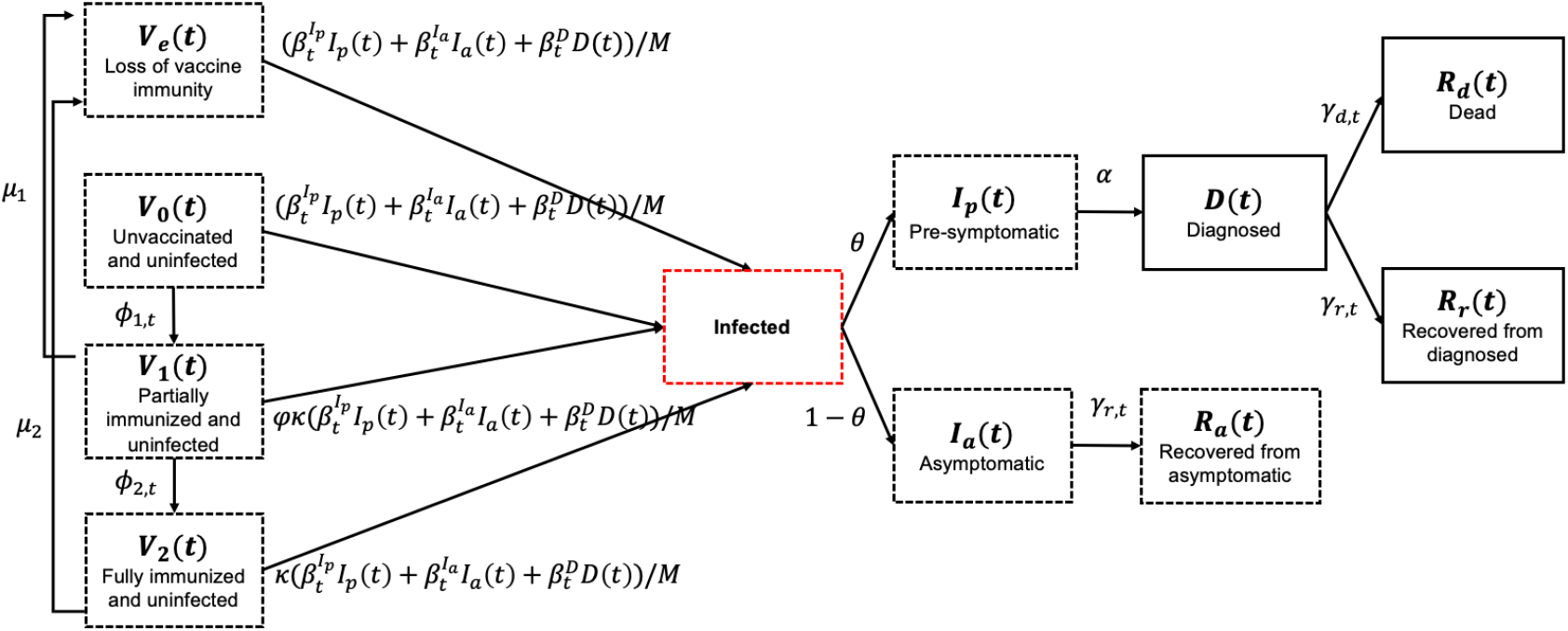
Compartments and state variables with a dynamic flow chart of the proposed vSVIADR model. The observable (unobservable) compartments are marked by solid (dashed) boxes. The red dashed infected box is a mix of pre-symptomatic and asymptomatic infections.

Let *V*_0_(*t*), *V*_1_(*t*), *V*_2_(*t*), *V*_*e*_(*t*) be counts at day *t* of four uninfected sub-populations having received no vaccine, with partial, full and expired vaccine immunity; and *I*_*a*_(*t*) and *I*_*p*_(*t*) be the counts of asymptomatic and pre-symptomatic infections, respectively, where the asymptomatic cases are never diagnosed, and the pre-symptomatic cases will be tested and confirmed in a future date, but not yet diagnosed at time *t*. The latter two compartments start two epidemiological pathways with that from asymptomatic *I*_*a*_(*t*) leading to the self recovered *R*_*a*_(*t*), and the symptomatic pathway from *I*_*p*_(*t*) to the diagnosed *D*(*t*), then to the recovered *R*_*r*_(*t*) and the dead *R*_*d*_(*t*). We combine *V*_0_ and *V*_*e*_ into one state *S*, and let *S*(*t*) = *V*_0_(*t*) + *V*_*e*_(*t*) be the counts of uninfected people without vaccine immunity whether due to receiving no vaccine or losing vaccine immunity at day *t*, ℱ_*t*_ = *σ*((*S*(*s*), *V*_1_(*s*), *V*_2_(*s*), *I*_*a*_(*s*), *I*_*p*_(*s*), *D*(*s*), *R*_*a*_(*s*), *R*_*r*_(*s*), *R*_*d*_(*s*)), *s* ≤ *t*) be the *σ*-algebra generated by the counts of all compartments up to time *t* and Δ*A*(*t*) = *A*(*t* + 1) − *A*(*t*) be the daily increment operator of a state variable *A* on day *t*.

Let 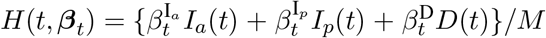 be the total infection loading at *t*, where 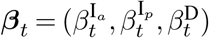. The vSVIADR model prescribes the conditional mean model

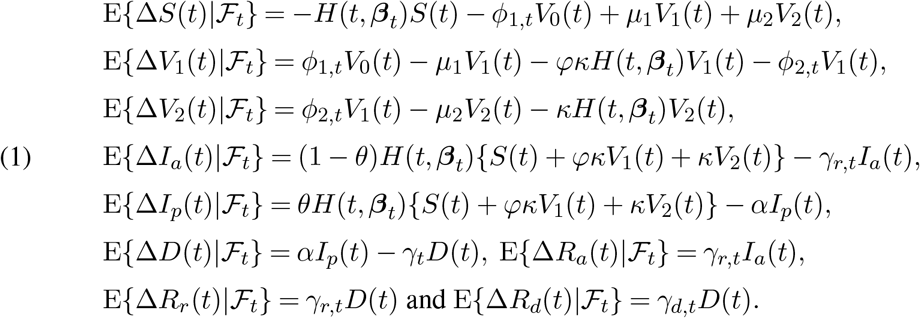

This form of the model connects well with the existing compartmental models defined via ODEs dating back to the SIR and SEIR models. However, as some compartments are latent, Model (1) is not enough to determine the joint distribution of the state variables.

Let 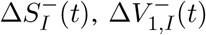 and 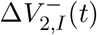 be the numbers of daily infected people from the three compartments *S, V*_1_ and *V*_2_ on day *t*, respectively; 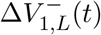 and 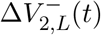 be the daily increments of people losing immunity from *V*_1_(*t*) and *V*_2_(*t*), respectively; and 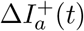 be the number of daily new asymptomatic cases on day *t*, where + (−) means inflow (outflow) from a compartment. Furthermore, let *N*(*t*) = *D*(*t*) + *R*_*d*_(*t*) + *R*_*r*_(*t*) be the accumulative confirmed cases, *G*_1_(*t*) and *G*_2_(*t*) be the accumulative numbers of people who have received at least one dose of vaccine and who are fully vaccinated, respectively. The following is the specification of the vSVIADR model via mutually independent Poisson distributions:

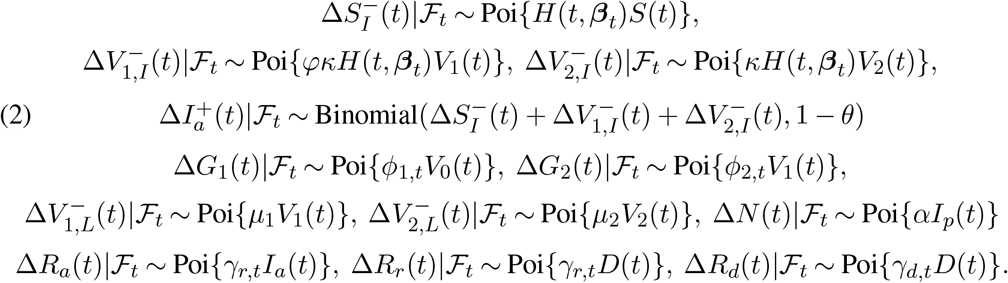

The Poisson assumptions can be relaxed into other distributions to accommodate potential overdispersion as discussed in Section 10. Given the initial {*S*(1), *V*_1_(1), *V*_2_(1), *I*_*a*_(1), *I*_*p*_(1), *D*(1), *R*_*a*_(1), *R*_*r*_(1), *R*_*d*_(1)} and based on the (2), the state variables progress according to (A.3) of the SM which are used to generate trajectories of the variables for parameter estimation. The relationship among Δ*G*_*i*_(*t*) and Δ*V*_*i*_(*t*), *i* = 1, 2, are explained by (A.3) in the SM; see Section S3 of the SM for details.

The infectious states in the vSVIADR model are the asymptomatic, pre-symptomatic, and diagnosed (*I*_*a*_, *I*_*p*_ and *D*) with time-varying infection rates 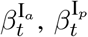 and 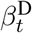, respectively. Asymptomatic and pre-symptomatic cases are not diagnosed at *t*, and asymptomatic cases are never confirmed through the infection. Thus, only the diagnosed state is observable. Since asymptomatic cases develop no symptom and diagnosed people are advised to isolate at home or hospitalized, their risks of transmission are lower than the pre-symptomatic cases in average at the population level. Therefore, we assumes the pre-symptomatic compartment to be more contagious than the other two compartments so that 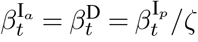 for a known constant *ζ* > 1, which was set as 5 according to the choice suggested in Yan et al. (2021). Results of a sensitivity analysis for different *ζ* are provided in Section 7.

Moreover, the vaccinated infected may have a lower transmission rate relative to the infected without vaccine protection, which implies heterogeneous infection rates among the non-vaccinated, partially vaccinated and fully vaccinated in each infected compartment. The current model has not taken this potential heterogeneity into account, partially due to unavailable data which keep track of the vaccination status of the infected.

Studies which evaluated neutralizing antibody titers, the durability of antibody responses, memory B-cell responses or cross-reactive T-cell responses after the vaccination showed the immune protection induced by the vaccination (Thiruvengadam et al., 2021; Davis et al., 2021; Barouch, 2022). Lee et al. (2022) reported the seroconversion rates for various types of immunocompromised patients, which showed the efficacy of COVID-19 vaccines in immunocompromised patients was less than that in immunocompetent controls (healthy people), but a second vaccine dose was associated with consistently improved seroconversion across various groups of immunocompromised patients. In the proposed vSVIADR model, vaccine effects of immunological barriers for the uninfected populations (*S, V*_1_, and *V*_2_) are reflected by the two vaccine effect parameters *φ* and *κ*. The full vaccination is assumed to reduce the infection rate by a factor *κ* ∈ [0, 1] relative to the unvaccinated, while the partial vaccination reduces the rate by *φκ* for a *φ* ∈ [0, 1*/κ*]. The VPR is 1 − *κ* and 1 − *φκ* for the fully and partially vaccinated under the model, respectively. If the full vaccination is fully effective, then *κ* = 0, while *κ* = 1 for complete failure.

Temporary immunity was assumed via *μ*_1_ and *μ*_2_ as 1*/μ*_1_ and 1*/μ*_2_ specify the respective average lengths of immunity after the partial and full vaccination. As the vaccinated people with lost or expired immunity are unobservable, *μ*_1_ and *μ*_2_ can not be estimated from daily epidemiological data, and their values have to be obtained from clinical studies. According to Doria-Rose et al. (2021) and Johnson & Johnson (2021), we set *μ*_1_ = 1*/*60 and *μ*_2_ = 1*/*240. It is noted that Dashtbali and Mirzaie (2021) and Giordano et al. (2021) assumed *μ*_1_ = *μ*_2_ = 0 (permanent immunity) and a regular flow from susceptibles with full efficacy of vaccines. The time-varying rate *ϕ*_1,*t*_ (*ϕ*_2,*t*_) of receiving the partial (full) vaccination for the susceptible (the partially vaccinated) people *S*(*t*) (*V*_1_(*t*)) can be estimated by the kernel smoothing on the daily number of unvaccinated (partially vaccinated) people receiving the partial (full) vaccination Δ*G*_1_(*t*) (Δ*G*_2_(*t*)).

Under the vSVIADR model, *θ* ∈ (0, 1) is the daily proportion of the pre-symptomatic cases and *α* is the diagnostic rate from the pre-symptomatic state *I*_*p*_ to the diagnosed state *D*. Implied from (2), the number of newly asymptomatic cases 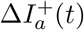 given the daily new infections follows Binomial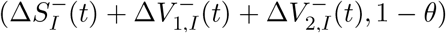. As the asymptomatic and pre-symptomatic cases are latent, similar as *μ*_1_ and *μ*_2_, we determine the value of *θ* based on existing studies. The meta-analysis of Buitrago-Garcia et al. (2020) found 20% (CI: 17% - 25%) of COVID-19 infections remained asymptomatic in 79 published studies, which led our setting *θ* = 0.8 in the analysis.

The effective reproduction number *R*_*t*_ is a key indicator and quantifies the mean number of secondary infections generated per primary infection at *t*. Derivation in the SM shows that

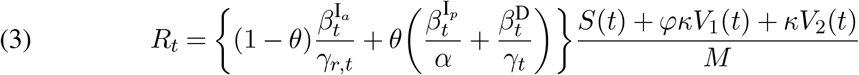

under the vSVIADR model. When *R*_*t*_ *>* 1(*<* 1), the epidemic is increasing (decreasing). From (3), *R*_*t*_ is conventionally driven positively by the three infection rates 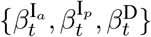, and negatively by the diagnosis rate *α*, the removal rate *γ*_*t*_ and the VPRs represented by 1 −*κ* and 1 − *φκ*. That vaccine slowing down *R*_*t*_ is seen by relocating *φκV*_1_(*t*) + *κV*_2_(*t*) from the susceptible population, as more people move from the group *S*(*t*) without vaccine immunity to the partially or fully vaccinated groups *V*_1_(*t*) and *V*_2_(*t*) with enhanced immunity.

Due to the unobservable states, identification for the stochastic vSVIADR model (2) is challenging. We provide a justification for identifiability under a deterministic version (1) of the vSVIADR model in Section S5 in the SM and conduct the sensitivity analyses of the estimates with respect to the choice of *θ* and *ζ* which are discussed in Section 7.

## 4. Estimation and inference

We consider the estimation of the model parameters *α*, 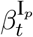, *γ*_*d,t*_, *γ*_*r,t*_, *φ* and *κ* for a country, which leads to the estimation of the effective reproduction number *R*_*t*_ by (3) as well as the VPRs. For each country, we denote its start date of the pandemic as *t* = 1, the start date of vaccination as *T*_1_, and the ending date as *T*. The observed data are 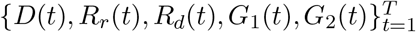.

We propose a multi-step decentralized estimation procedure as displayed by the flow chart in Figure 2, that estimates the constant *α, φ* and *κ* by minimizing certain criterion functions, and estimates the time-varying parameters by the nonparametric regression method. The decentralization implies using different periods of data to estimate different parameters.

**Fig 2:**
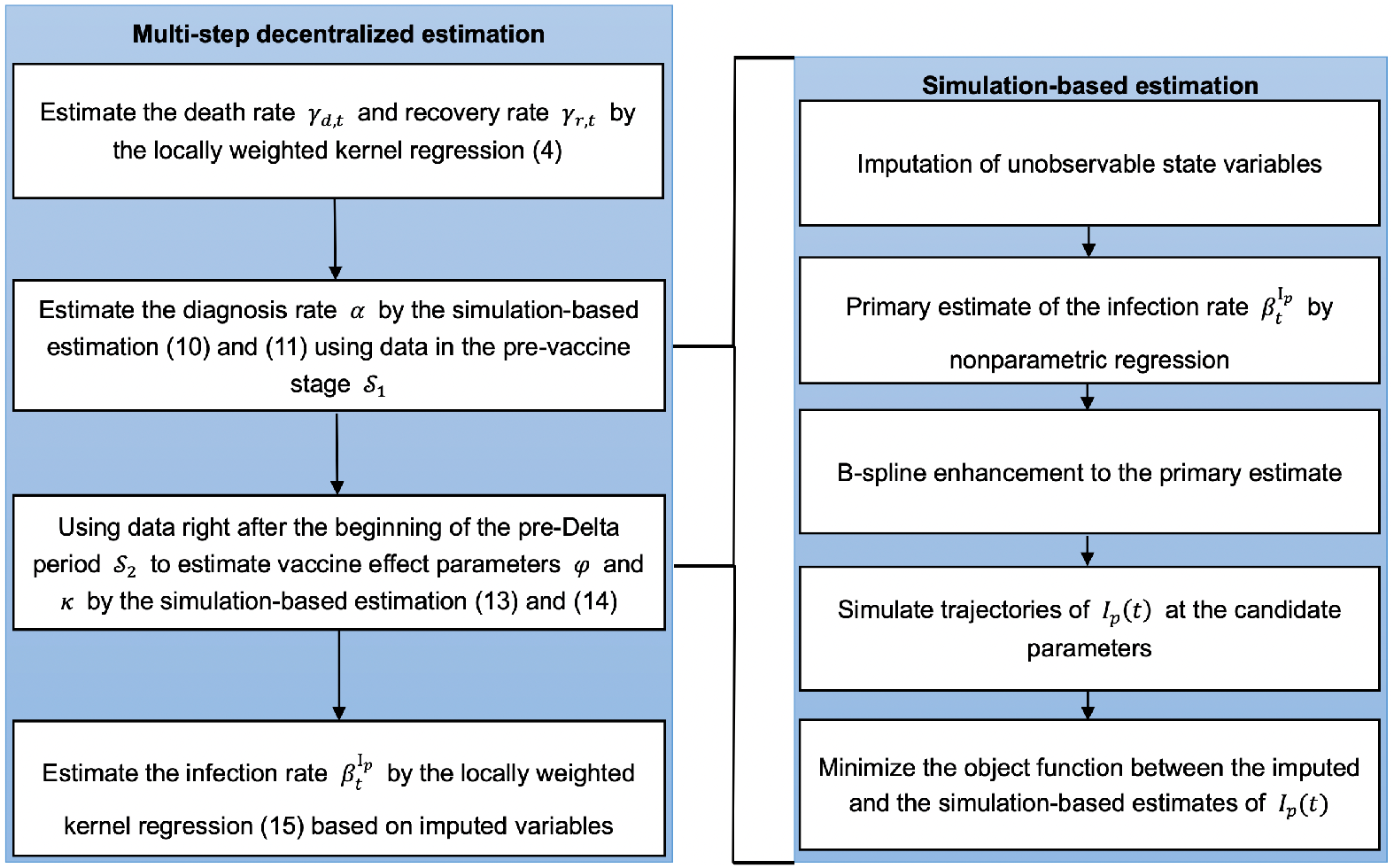
The flowchart of the multi-step decentralized estimation procedure, whose main steps are outlined in the left panel. And the right panel provides the detailed procedure of the simulation-based estimation method used in the second and third steps in the left panel.

### 4.1. Estimation of removal rates γ_d,t_ and γ_r,t_

The estimation of the two removal parameters is the most straightforward among all the parameters, as the three compartments *D*(*t*), *R*_*r*_(*t*) and *R*_*d*_(*t*) involving the recovery process are observable largely because they are located at the end of the epidemiological process. From the Poisson increments of the daily new deaths Δ*R*_*d*_(*t*) and daily new recoveries Δ*R*_*r*_(*t*) specified in (2), we have E{Δ*R*_*d*_(*t*)|*D*(*t*)} = *γ*_*d,t*_*D*(*t*) and E{Δ*R*_*r*_(*t*)|*D*(*t*)} = *γ*_*r,t*_*D*(*t*). The time-varying nature of the parameters suggests the locally weighted kernel smoothing estimator of *γ*_*r,t*_ and *γ*_*d,t*_ by regressing Δ*R*_*r*_(*t*) and Δ*R*_*d*_(*t*) on *D*(*t*) without intercept, respectively. Specifically, the estimators are in the form

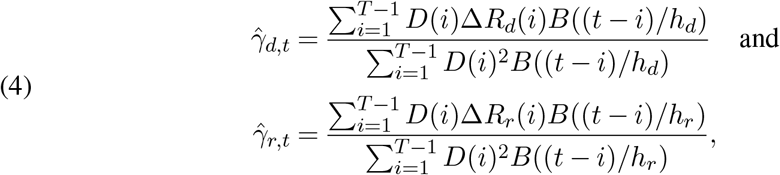

where *B*(*·*) is a boundary kernel modified from a symmetric kernel, and *h*_*d*_ and *h*_*r*_ are the temporal smoothing bandwidths, whose expression is given in the SM. The boundary kernel (Jones, 1993) is used to account for the boundary bias associated with the non-parametric regression estimation near the ending time of the analysis. It is a standard method to counter the discontinuity at a boundary in nonparametric curve estimation.

### 4.2. Estimation of diagnosis rate α

We consider using data in the pre-vaccine stage 𝒮_1_ = {*t*_1_, …, *t*_2_} ∈ {1, …, *T*_1_} to estimate the diagnosis rate *α*, which avoids the interference from estimating the vaccine effect parameters *φ* and *κ*. The challenge in estimating *α* lies in the pre-symptomatic compartment *I*_*p*_(*t*) being latent. Our strategy is to minimize a contrast measure with respect to *α* and 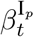 in the form of

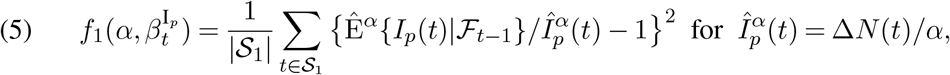

where 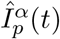 is an imputed value of *I*_*p*_(*t*) using the daily increment Δ*N*(*t*) of confirmed cases at a given *α* by noting E{Δ*N*(*t*)|ℱ_*t*_} = *αI*_*p*_(*t*), and Ê^*α*^{*I*_*p*_(*t*)|ℱ_*t*−1_} is a simulation-based estimate of E{*I*_*p*_(*t*)|ℱ_*t*−1_} by averaging the simulated trajectories according to (2) and (A.3) at the given *α* and 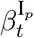. The construction of Ê^*α*^{*I*_*p*_(*t*)|ℱ_*t*−1_} requires (i) obtaining initial values of all compartments at *t*_1_ via imputation; (ii) a preliminary estimate of the time-varying 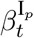 by the kernel method based on imputed variables; and (iii) a B-spline enhancement to the preliminary estimate of 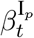 for better and more stable fitting of 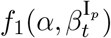. Throughout the section, the superscript *α* indicates the computed quantities’ dependence on *α*.

The observable *D*(*t*), *R*_*r*_(*t*) and *R*_*d*_(*t*) can be used as the initial values. For those unobservable state variables, 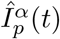 can serve as a replacement for *I*_*p*_(*t*). To impute *I*_*a*_(*t*), since

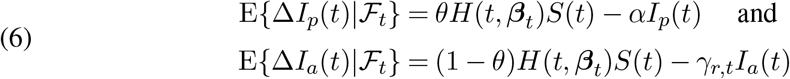

in the pre-vaccine period, replacing *θH*(*t*, ***β***_*t*_)*S*(*t*) by Δ*I*_*p*_(*t*) + *αI*_*p*_(*t*), we have Δ*I*_*a*_(*t*) ≈ {Δ*I*_*p*_(*t*) + *αI*_*p*_(*t*)}(1 − *θ*)*/θ* − *γ*_*r,t*_*I*_*a*_(*t*). Thus, {*I*_*a*_(*t*)} can be imputed sequentially by

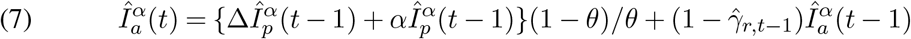

in the pre-vaccine period, where 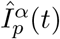 is given in (5) and 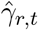 is the estimator attained in (4). Furthermore, as E{Δ*R*_*a*_(*t*)|ℱ_*t*_} = *γ*_*r,t*_*I*_*a*_(*t*), *R*_*a*_(*t*) in the pre-vaccine period is imputed by

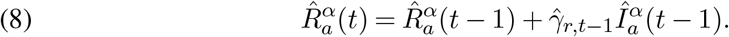

Then, *S*(*t*) can be imputed by 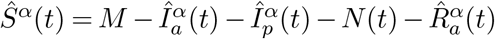, giving the initial values 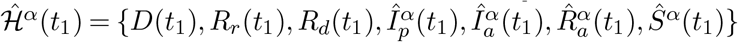 at *t*_1_ for given *α*.

To minimize the objective function 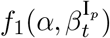 with respective to *α* and the varying coefficient 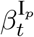 in (5), we approximate 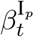 by the B-spline

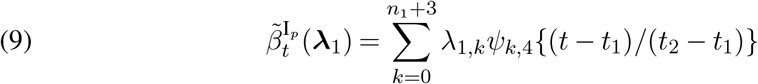

for *t* ∈ 𝒮_1_, where 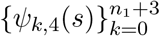 are the order four basis functions supported on [0, 1], 0 ≤ *n*_1_ *< t*_2_ − *t*_1_ − 3 is the number of equally spaced internal knots within [0, 1], and 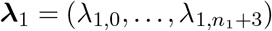 is the coefficient vector of the splines. This makes the objective function (5) take the form 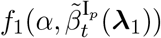, which makes the optimization seemingly parametric. A data driven procedure is proposed in Section S6 in the SM to determine a plausible range Λ_*α*_ of the parameter ***λ***_1_ for minimizing 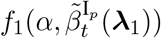, where Λ_*α*_ is given in (A.9) and (A.10).

Given initial values 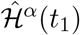 at *t*_1_, the candidate parameters *α* and 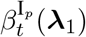 with ***λ***_1_ ∈ Λ_*α*_, and the estimated removal rates 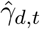 and 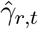, we can simulate all nine states in the proposed vSVIADR model for *t* ∈ 𝒮_1_ based on (2) and (A.3), with 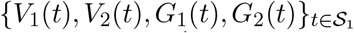 being set as zero for the pre-vaccine period. This leads to the estimator Ê^*α*^{*I*_*p*_(*t*)|ℱ_*t*−1_} and the evaluation of 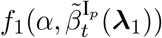. Specifically, let 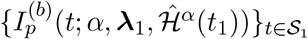 be the *b*th simulated trajectory of the pre-symptomatic infections *I*_*p*_(*t*) over 𝒮_1_ for *b* = 1, …, *B* for a reasonably large integer *B*, which was set as 300 in the empirical analysis. All *B* trajectories are independently generated. We use the average simulated value 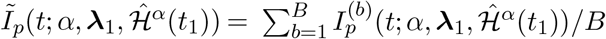 to estimate E{*I*_p_(*t*)|ℱ_*t−1*_} given the candidate *α* and ***λ***_1_. Then, the goodness-of-fit criterion function in (5) can be formulated by profiling out ***λ***_1_ at a given *α* as

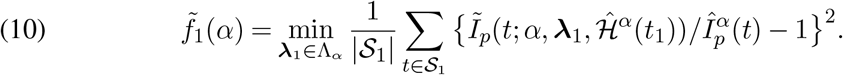

The estimation of *α* is obtained by minimizing 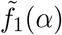 over 𝒜 so that

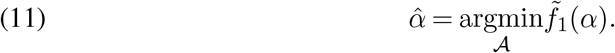

Based on the clinical information (Guan et al., 2020), we chose 𝒜 = [0.1, 0.2] implying the average diagnosis time is from 5 to 10 days before the start of public vaccination. By discretizing the search domain, the optimization problems (10) and (11) turn into the grid search in the space formed by *n*_1_ +5 parameters 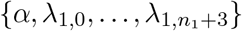. Other numerical algorithms, such as the genetic algorithm (Holland, 1992) may be applied to solve the optimization problems. We chose the grid search to ensure more accuracy. The detailed optimization procedure is provided in Section S6 in the SM.

We chose 𝒮_1_ to be a 30-day period. The relatively short period permitted less number of spline basis functions to model 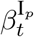 in order to save computational cost. From the regression estimation equation (A.7) in the SM, larger *I*_*p*_(*t*) and *D*(*t*) would make the estimate of 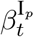 more stable, we intend to choose 𝒮_1_ around the peak 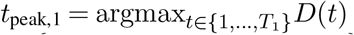 of *D*(*t*) before *T*_1_. Specifically, if *t*_peak,1_ + 15 *< T*_1_, we set 𝒮_1_ = {*t*_peak,1_ − 15, …, *t*_peak,1_ + 15}, otherwise, we set 𝒮_1_ = {*T*_1_ − 31, …, *T*_1_ − 1}. As shown in Figure S3 in the SM, the peak infection times *t*_peak,1_ of the 10 countries were within 55 days from *T*_1_ except India and Peru, which were, respectively, 113 and 161 days ahead of their respective vaccine start date *T*_1_.

### 4.3. Estimation of vaccine effects φ and κ

We consider data in a period 𝒮_2_ = {*t*_3_, …, *t*_4_} ⊂ {*T*_1_, …, *T*_1_ + *l*_1_} right after the start of vaccination to estimate the vaccine effect parameters *φ* and *κ* in the pre-Delta period. Similar to the objective function 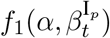 for estimating *α* in (5), we minimize the contrast measure

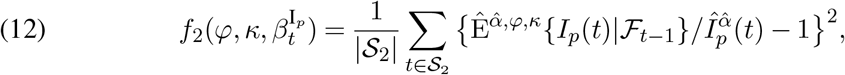

where 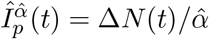 is the imputed value of *I*_*p*_(*t*) by the estimated diagnosis rate 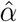 obtained in (11), and 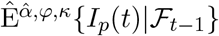 is the simulation-based estimate of E{*I*_*p*_(*t*)|ℱ_*t*−1_} at the given *φ, κ* And 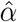 by averaging the simulated trajectories of the proposed vSVIADR model, in the same way as the construction of 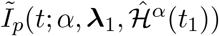 for the pre-vaccine period.

Let 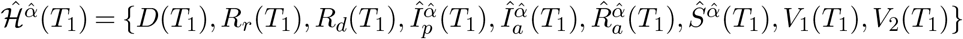 be the imputed state variables at time *T*_1_, where 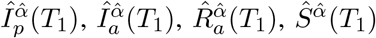 are imputed by (5), (7) and (8) with the estimated diagnosis rate 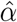, and *V*_1_(*T*_1_) = *V*_2_(*T*_1_) = 0. To obtain the trajectory of *I*_*p*_(*t*), similar as Section 4.2, we also use the B-spline model 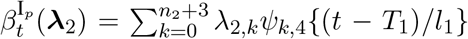 to approximate the infection rate 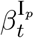 for *t* ∈ {*T*_1_, …, *T*_1_ + *l*_1_} right after the start of vaccination, where *n*_2_ is the number of internal knots and 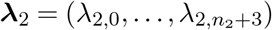 is the coefficient vector. Then, the objective function in (12) becomes 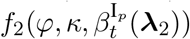. A data driven procedure is proposed in Section S7 in the SM to determine a plausible range Λ_2_ of the parameter ***λ***_2_.

Let Θ be the candidate set of the vaccine parameters (*φ, κ*), which is set to {(*φ, κ*) : 0.5 ≤ *φ* ≤ 10, 0.01 ≤ *κ* ≤ 1, *φκ* ≤ 1} based on the reported empirical VEs in recent studies summarized in Tables S1 and S2 in the SM. Then, the simulation of trajectories is generated from the proposed model. Specifically, given the initial values 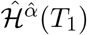 at time *T*_1_, and the candidate model parameters (*φ, κ*) ∈ Θ, 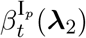 with ***λ***_2_ ∈ Λ_2_ together with the estimated diagnosis rate 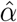 and the estimated removal rates 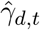 and 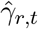, we simulate all nine state variables for *t* ∈ {*T*_1_, …, *T*_1_ + *l*_1_} according to the specification in (2) and (A.3). However, we do not generate *G*_1_(*t*) and *G*_2_(*t*) by the Poisson increments shown in (2) but directly use their observations, since they are irrelevant to the parameters of our interests, *φ, κ* and 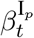.

Let 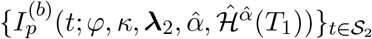 be the *b*th simulated trajectory of the infected and pre-symptomatic cases over the target interval 𝒮_2_ for *b* = 1, …, *B*, where *B* was 300 in the empirical analysis, and all the trajectories are independently generated. The average 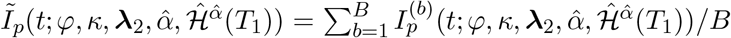 is used estimate E{*I*_*p*_(*t*)|ℱ_*t*−1_} after the start of vaccination at the candidate parameters *φ, κ* and ***λ***_2_. Then, the criterion function (12) can be formulated by profiling out ***λ***_2_ at the given *φ* and *κ* as

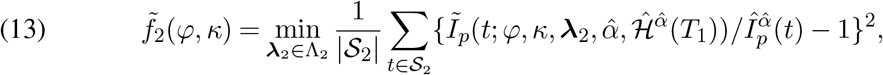

and the estimator of (*φ, κ*) is

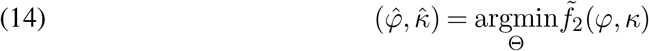

by conducting a grid search in the space formed by *n*_2_ +6 parameters 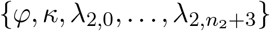. The detailed optimization procedure is provided in Section S7 in the SM.

The vaccine takes time to be effective. Usually, two weeks are required to form the protective effects against SARS-CoV-2 infections after receiving a dose. We chose a 30-day period 18 days after the start of the vaccination {*T*_1_ + 18, …, *T*_1_ + 48} as 𝒮_2_ in the estimation of the vaccine effect. The 18 days were slightly more than the 14 days recognised by the WHO for full vaccine effect, which was based on a slightly better performance in the simulation studies reported in Table S5 in the SM.

The estimation of (*φ, κ*) in the intervening and the Delta-dominated periods uses the same procedure as outlined above except that the starting date and the initial conditions need to be adjusted. The lengths of the target periods and the number of B-spline knots may be different to suit each period’s situation.

### 4.4. Estimation of infection rate 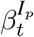

After having estimated the vaccine effects *φ* and *κ* and the diagnosis rate *α*, the smoothing estimator 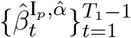 evaluated at 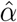 in Equation (A.8) in the SM can be used to estimate the infection rates in the pre-vaccine period. We need to estimate the infection rate function in the post-vaccine periods. We do not use the spline estimate for 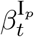 used in Sections 4.2 and 4.3, since after obtaining 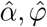 and 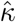, a more tangible estimator based on the kernel regression estimator similar to (A.8) can be formulated in the post-vaccine era. However, we need to update the imputation by incorporating the vaccine compartments *V*_1_(*t*) and *V*_2_(*t*) to suit the post-vaccine situation as shown below.

From the conditional mean of Δ*I*_*p*_(*t*) in (1), we have the approximation Δ*I*_*p*_(*t*)+*αI*_*p*_(*t*) ≈*θH*(*t*, ***β***_*t*_){*S*(*t*) + *φκV*_1_(*t*) + *κV*_2_(*t*)}. Therefore, the conditional-mean-based imputation of *I*_*p*_(*t*), *I*_*a*_(*t*) and *R*_*a*_(*t*) via (5), (7) and (8) would remain the same, which is free of *V*_1_(*t*), *V*_2_(*t*), *φ* and *κ* regardless being before or after vaccination. Let 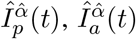 and 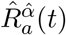 be such imputed values for *t > T*_1_ with the estimated diagnosis rate 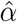. To impute *V*_1_(*t*) and *V*_2_(*t*), note that {Δ*I*_*p*_(*t*) + *αI*_*p*_(*t*)}*/*[*θ*{*S*(*t*) + *φκV*_1_(*t*) + *κV*_2_(*t*)}] can serve as a substitution for the total infection loading *H*(*t*, ***β***_*t*_). From Equation (A.3), we can impute *V*_1_(*t*) and *V*_2_(*t*) for *t > T*_1_ by

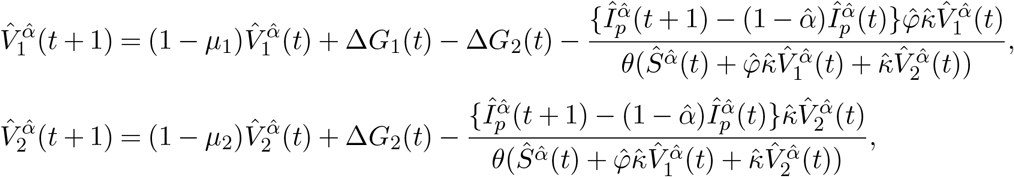

with the initial values *V*_1_(*T*_1_) = *V*_2_(*T*_1_) = 0. Then, 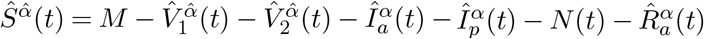 is the imputed value of *S*(*t*).

With the updated definitions of 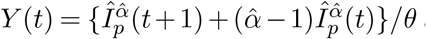 and 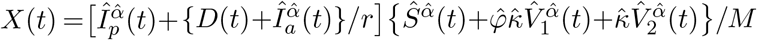 in the post-vaccine period (*t > T*_1_), for *t* = *T*_1_ + 1, …, *T* − 2, the infection rate 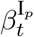 can be estimated by kernel regression,

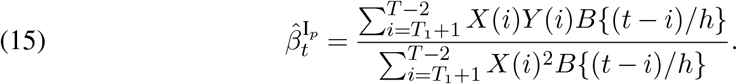

The regression use *Y*(*t*) and *X*(*t*) until *T* − 2 since the imputed values 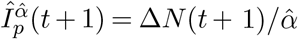 used in *Y*(*t*) can only be obtained for *t* ≤ *T* − 2. Note that the estimator 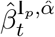 in (A.8) for the infection rates in the pre-vaccine period is a special case of (15) with 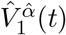 and 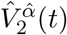 set to zero for *t* ∈ {1, …, *T*_1_}. The estimation procedure of 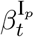 in the intervening and Delta-dominated period is the same except using each period’s 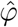 and 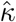.

### 4.5. Parametric bootstrap inference

Let 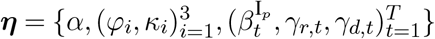 be the epidemiological parameters of our concern in the whole study period, where 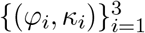 are the vaccine effect parameters for the pre-Delta, intervening and Delta-dominated periods, respectively. And we denote the generation process by our proposed SEM based on (2) and (A.3) at the given ***η*** as vSVIADR(***η***). To obtain an uncertainty measure for these epidemiological parameters, we consider using the parametric bootstrap procedure under the vSVIADR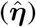 model with the estimated parameters 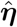.

Specifically, let 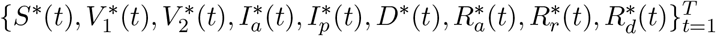 be a resampled trajectory of the entire state variables from the bootstrap. Given 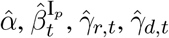, and the observed and imputed initial values 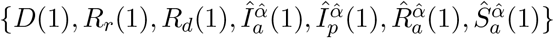, bootstrap resampled trajectories for the first *T*_1_ days in the pre-vaccine period were generated by substituting these estimates into (2) and (A.3) with setting 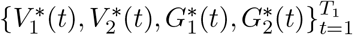 to zero. Then, the bootstrap resampling for the pre-Delta period was conducted via vSVIADR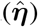 with the initial value 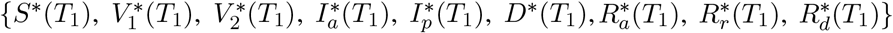 according to (2) and (A.3). For the resampling in this period, we did not resample *G*_1_(*t*) and *G*_2_(*t*) but used their original observations since they are irrelevant to ***η*** which can be seen in (2). The following bootstrap resampled data in the intervening and Delta-dominated period can be generated in the same way with the estimates for the corresponding estimated parameters in 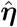.

The parameter ***η*** was re-estimated based on the bootstrap resamples. The resampling was replicated for a large number (*B*) of times to obtain *B* independent bootstrap estimates and 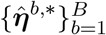 for the parameter. The sample standard deviation of the bootstrap estimates can be used to estimate the standard error of 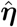. Let 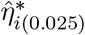 and 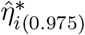 be, respectively, the 2.5th and 97.5th percentile of the bootstrap estimates for *η*_*i*_, the *i*th element of ***η***. Then, the 95% percentile bootstrap confidence interval of *η*_*i*_ is 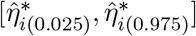. Furthermore, let the vaccine effect parameter for full vaccination in country *j* in period *i* be 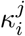. Then, we can use the 95% one-sided bootstrap confidence interval of 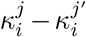 to formulate a pairwise bootstrap test for the difference in the VPRs of the two countries *j* and *j*′. Specifically, let 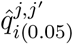 and 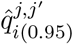 be the 5th and 95th percentile of the bootstrap distribution for 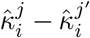, respectively, then the VPR of the full vaccination in period *i* in country *j* is significantly lower (higher) than that in country *j*′ if 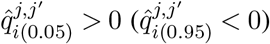. Similar tests can be conducted for VPRs of the partial vaccination.

## 5. Simulation experiments

To illustrate the performance of the proposed estimation method and the asymptotic performance of the estimators as the population size *M* increases, we designed a simulation in which the length of data before and after the start of vaccination was *T*_1_ = 300 and *T*_2_ = 50 days, respectively. To mimic the COVID-19 reality, we set *r* = 5, *α* = 0.15, *μ*_1_ = 1*/*60, *μ*_2_ = 1*/*240, *θ* = 0.8, (*φ, κ*) ∈ {(2.5, 0.1), (1.5, 0.4)}, *M* ∈ {5 × 10^8^, 1 × 10^9^, 1.5 × 10^9^}, *I*_*a*_(1)*/M* = 5 × 10^−8^, *I*_*p*_(1)*/M* = 2 × 10^−7^, *D*(1)*/M* = 3 × 10^−8^, *R*_*r*_(1)*/M* = 4 × 10^−8^, *R*_*a*_(1) = *R*_*d*_(1) = *G*_1_(1) = *G*_2_(1) = 0, 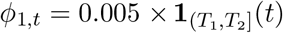 and 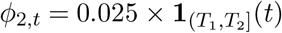, where **1**_*A*_(*t*) is the indicator function with value 1 if *t* ∈ *A*, and 0 otherwise. For the varying coefficient parameters, to make the simulation in line with the data in real-world situations, we mimicked the pattern of the estimated infection, recovery and death rates of the US in the pre-vaccine period. Specifically, we set 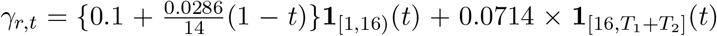 for the recovery rate, 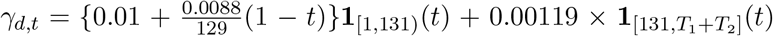 for the death rate which linearly decreases in the first 130 days and then remains constant, and 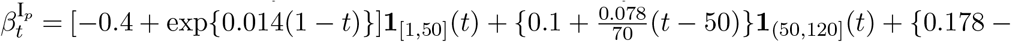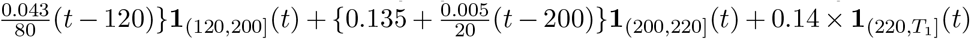 for the pre-vaccine period *t* ≤ *T*_1_, which consists of segments of exponential decrease, linear increase, linear decrease and constant functions. See Figure S4 in the SM for the comparison of the piece-wise curves in our setting with those of the US estimates.

For each combination of the parameter settings, we considered three different settings of the infection rate after the vaccination: (i) constant trend 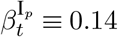; (ii) increasing trend 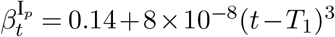; and (iii) decreasing trend 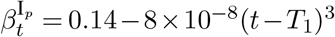. For each parameter setting, the trajectories of all compartments were generated by adding up the Poisson increments over time according to (2) and (A.3), and this process was independently repeated 100 times. For each repetition, we use the estimation method described in Section 4 to obtain 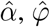 and 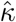 by (11) and (14), and then the infection rate by (15).

Table 1 reports the estimation results for *α* and the vaccine effects (1 − *φκ*, 1 − *κ*) for the partial and fully vaccinated group, which are equal to (0.75, 0.9) and (0.4, 0.6) under the settings of (*φ, κ*) = (2.5, 0.1) and (*φ, κ*) = (1.5, 0.4), respectively. Figures 3 and S5-S9 in the SM display the estimation results of the time-varying coefficients 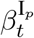, *γ*_*d,t*_, *γ*_*r,t*_ and *R*_*t*_, with the comparison to their true values. These results reveal generally satisfactory performance of the estimation procedure, since all the confidence intervals of the parameters covered the true values although some were quite narrow due to the large infection size. For each setting of parameters, the standard errors of the estimates over 100 replications decreased (Table 1) and the confidence intervals of the parameters became smaller (by comparison among the three panels of Figure 3) with the increase of the population size. Generally, the bias of the estimates also decreased as the population size increased. From Figure 3, the differences between the estimates 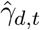 and 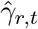s and their true values suggest that their variation decreased with time *t* due to the increasing number of infected people (sample size). There was no obvious trend in the variations of 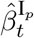 and 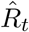 over time since our multi-step decentralized estimation made the variation of these estimates be also influenced by the variation of the other parameters, including 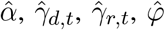 and 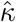. We also considered the simulation with another set of initial values. The results are reported in Table S6 in the SM, which were close to the estimated VPRs in Table 1.

**Table 1.**
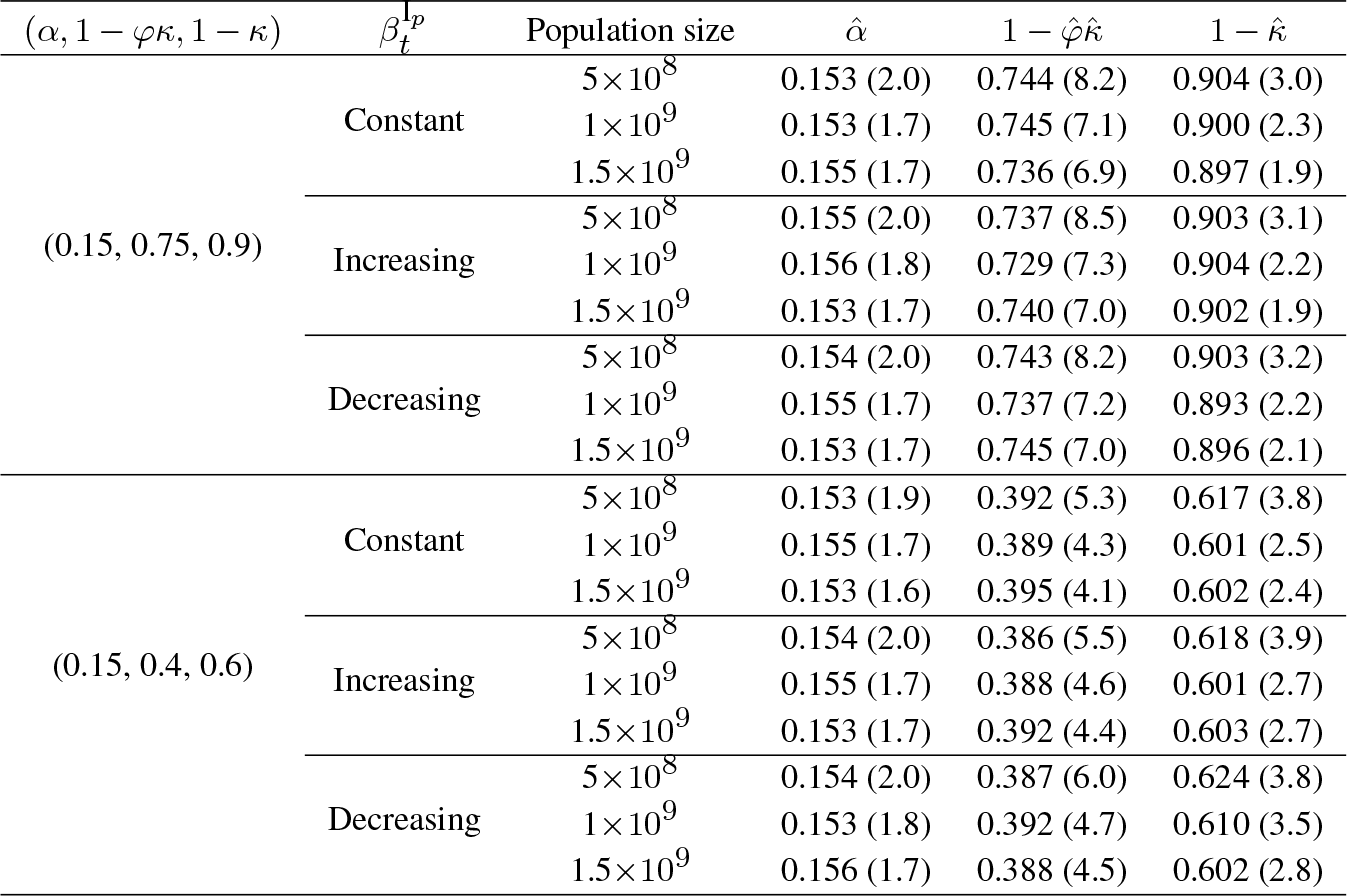
Average estimates (10^3^× the standard errors) of the diagnosis rate α, the vaccine protection rates 1 − φκ and 1 − κ for the partial and full vaccination, respectively, under different parameter settings based on 100 simulations.

**Fig 3:**
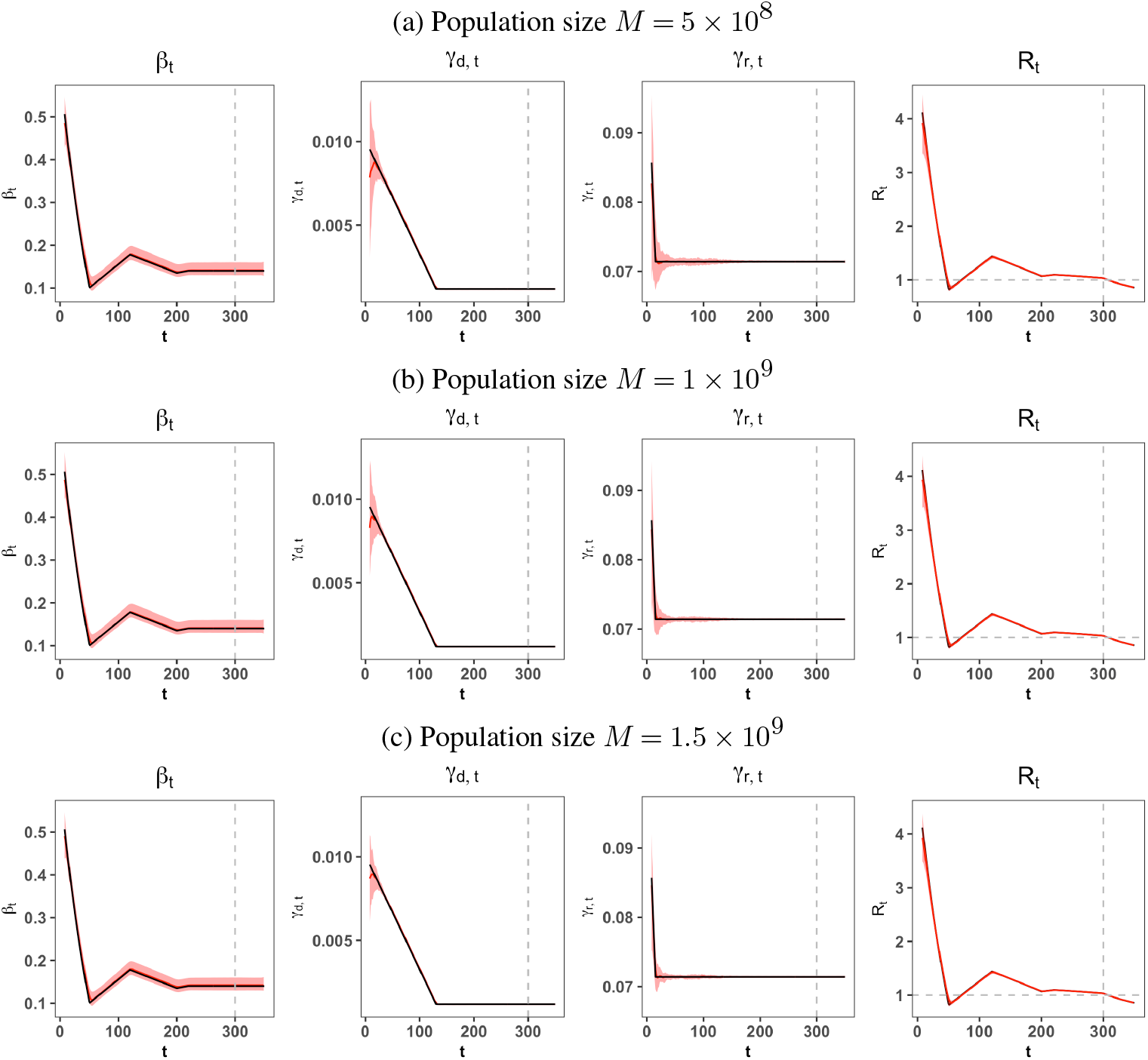
Curves of true (black) and estimated (red) 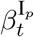, *γ*_*r,t*_, *γ*_*d,t*_ and *R*_*t*_ with the colored 2.5%-97.5% quantile bands for three population sizes. The dashed vertical line marks the start of vaccination. The true values of infection rates after the start of the vaccination were 0.14, and *α, φ* and *κ* were 0.15, 2.5 and 0.1, respectively.

## 6. Results on vaccine protection rates

We applied the proposed SEM and the inference procedures presented in Section 4 to evaluate the real-world VPRs in the ten countries, which are represented by 1 − *φκ* and 1 − *κ* for the partial and full vaccination under our model, respectively. We estimated the VPRs in the three periods (the pre-Delta, intervening and Delta-dominated) after vaccination for the ten countries. The results are reported in Table 2. India did not have the pre-Delta period after vaccination as stated in Section 2. It is noted that the VPR estimated in the Delta period was not against the Delta variant only, but against all strains that occurred in this period.

**Table 2.** The estimated vaccine protection rates of the partial and full vaccination in the pre-Delta, the intervening and the Delta-dominated periods for the 10 countries. The standard errors obtained by the bootstrap method are reported in the parentheses.

| (a) Pre-Delta period |  |  |  |  |
| --- | --- | --- | --- | --- |
| Country | Time | Vaccine | Partial | Full |
| Brazil | 2021-01-16 ~ 2021-05-19 | AstraZeneca, Sinovac | 0.625 (0.07) | 0.75 (0.03) |
| Canada | 2020-12-13 ~ 2021-03-14 | Moderna, AstraZeneca, Pfizer | 0.58 (0.07) | 0.86 (0.02) |
| Germany | 2020-12-26 ~ 2021-02-28 | Janssen, Moderna, AstraZeneca, Pfizer | 0.56 (0.07) | 0.89 (0.02) |
| Italy | 2020-12-26 ~ 2021-04-01 | Janssen, Moderna, AstraZeneca, Pfizer | 0.58 (0.07) | 0.94 (0.01) |
| Peru | 2021-02-07 ~ 2021-06-09 | AstraZeneca, Pfizer, Sinopharm | 0.64 (0.06) | 0.76 (0.04) |
| Portugal | 2020-12-26 ~ 2021-04-04 | Moderna, AstraZeneca, Pfizer | 0.52 (0.06) | 0.68 (0.04) |
| Turkey | 2021-02-11 ~ 2021-04-27 | Sinovac, Pfizer | 0.48 (0.05) | 0.74 (0.04) |
| UK | 2021-01-09 ~ 2021-02-21 | AstraZeneca, Pfizer | 0.52 (0.04) | 0.68 (0.03) |
| US | 2020-12-12 ~ 2021-02-22 | Moderna, Pfizer | 0.55 (0.07) | 0.95 (0.01) |
| Average (SE) |  |  | 0.562 (0.017) | 0.806 (0.035) |

| (b) Intervening period |  |  |  |  |
| --- | --- | --- | --- | --- |
| Country | Time | Vaccine | Partial | Full |
| Brazil | 2021-05-20 ~ 2021-08-15 | Pfizer, AstraZeneca, Sinovac | 0.52 (0.05) | 0.68 (0.03) |
| Canada | 2021-03-15 ~ 2021-07-04 | Moderna, AstraZeneca, Pfizer | 0.45 (0.05) | 0.78 (0.03) |
| Germany | 2021-03-01 ~ 2021-07-04 | Janssen, Moderna, AstraZeneca, Pfizer | 0.505 (0.05) | 0.67 (0.03) |
| India | 2021-01-15 ~ 2021-04-11 | Covaxin, AstraZeneca | 0.46 (0.05) | 0.64 (0.04) |
| Italy | 2021-04-02 ~ 2021-07-04 | Janssen, Moderna, AstraZeneca, Pfizer | 0.55 (0.05) | 0.7 (0.02) |
| Peru | 2021-06-10 ~ 2021-09-12 | AstraZeneca, Pfizer, Sinopharm | 0.46 (0.06) | 0.73 (0.04) |
| Portugal | 2021-04-05 ~ 2021-05-23 | Janssen, Moderna, AstraZeneca, Pfizer | 0.38 (0.06) | 0.69 (0.03) |
| Turkey | 2021-04-28 ~ 2021-06-20 | Sinovac, Pfizer | 0.235 (0.05) | 0.49 (0.03) |
| UK | 2021-02-22 ~ 2021-05-23 | Moderna, AstraZeneca, Pfizer | 0.46 (0.06) | 0.64 (0.04) |
| US | 2021-02-23 ~ 2021-06-20 | Janssen, Moderna, Pfizer | 0.675 (0.05) | 0.87 (0.02) |
| Average (SE) |  |  | 0.470 (0.036) | 0.689 (0.031) |

| (c) Delta-dominated period |  |  |  |  |
| --- | --- | --- | --- | --- |
| Country | Time | Vaccine | Partial | Full |
| Brazil | 2021-08-16 ~ 2021-11-20 | Janssen, Pfizer, AstraZeneca, Sinovac | 0.385 (0.05) | 0.59 (0.03) |
| Canada | 2021-07-05 ~ 2021-11-20 | Moderna, AstraZeneca, Pfizer | 0.28 (0.05) | 0.64 (0.03) |
| Germany | 2021-07-05 ~ 2021-11-20 | Janssen, Moderna, AstraZeneca, Pfizer | 0.40 (0.04) | 0.60 (0.03) |
| India | 2021-04-12 ~ 2021-11-20 | Covaxin, AstraZeneca | 0.325 (0.05) | 0.55 (0.03) |
| Italy | 2021-07-05 ~ 2021-11-20 | Janssen, Moderna, AstraZeneca, Pfizer | 0.355 (0.02) | 0.57 (0.01) |
| Peru | 2021-09-13 ~ 2021-11-20 | AstraZeneca, Pfizer, Sinopharm | 0.415 (0.04) | 0.61 (0.03) |
| Portugal | 2021-05-24 ~ 2021-11-20 | Janssen, Moderna, AstraZeneca, Pfizer | 0.37 (0.05) | 0.58 (0.03) |
| Turkey | 2021-06-21 ~ 2021-11-20 | Sinovac, Pfizer | 0.175 (0.04) | 0.45 (0.03) |
| UK | 2021-05-24 ~ 2021-11-20 | Moderna, AstraZeneca, Pfizer | 0.34 (0.05) | 0.56 (0.03) |
| US | 2021-06-21 ~ 2021-11-20 | Janssen, Moderna, Pfizer | 0.48 (0.05) | 0.74 (0.03) |
| Average (SE) |  |  | 0.353 (0.026) | 0.589 (0.023) |

Table 2 shows that the overall VPRs of the full vaccination during the pre-Delta period in the 9 countries without India ranged from 68% to 95% (Average: 81%, SE: 4%) while those of the partial vaccination ranged from 48% to 64% (Average: 56%, SE: 2%), which suggested that the full vaccination would bring 12%-40% (Average:24%, SE: 4%) more VPR than the partial vaccination. Hence, the full vaccination’s VPRs regardless of the brands and their combinations all passed the 50% threshold for being effective according to the WHO guideline (WHO, 2021), while the partial vaccination’s VPRs largely passed the threshold except Turkey whose was at 48%.

The full vaccination’s VPRs of the countries in the intervening period ranged from 49% to 87% (Average: 69%, SE 3%) and those of the partial vaccination ranged from 23.5% to 67.5% (Average: 47%, SE 4%). The paired t-test for the mean VPRs in the nine countries in the two periods showed that the mean VPRs in the intervening period were significantly lower than those in the Pre-Delta period for the partial vaccination (p-value 0.016) and the full vaccination (p-value 0.005). And the full vaccination offered 15%-33% (Average: 22%, SE 2%) more protection than the partial one. When the Delta variant was prevalent, the partial vaccination showed even lower VPRs ranging from 17.5% to 48% (Average: 35%, SE 3%).

Only the US’s and Peru’s partial VPRs stayed above 40%. Thus, the partial vaccination was not sufficiently protective against the Delta strain in the ten countries, which was consistent with the findings in Li et al. (2021) based on a small size retrospective study and Planas et al. (2021) via investigating the neutralising capacity of sera from vaccine recipients. The VPRs of the full vaccination during the Delta period ranged from 45% to 74% (Average: 59%, SE: 2%), with the highest being 74% (CI: 69% to 79%) in the US and the lowest 45% (CI: 40% to 50%) in Turkey. The full vaccination during the Delta period had 19.5%-36% (average 24%, SE 2%) premium beyond the partial vaccination, indicating the boosting effect of the full vaccination against the Delta variant.

Table 2 indicates waning VPRs as the Delta variant gradually became the dominant strain, whether for partial or full vaccination. Compared to the pre-Delta period, the VPRs of full vaccination in the Delta-dominated period for the 9 countries without India decreased by 10%-37% (average 21%, SE 3%), while those for partial vaccination decreased by 7%-30.5% (average 21%, SE 2%). Relative to the intervening period, the VPRs of full vaccination in the Delta-dominated period were reduced by 4%-14% (average 10%, SE 1%), and those of the partial vaccination by 1%-19.5% (average 12%, 2%). The paired t-test for the mean VPRs in the ten countries in the two periods showed that the mean VPRs in the Delta-dominated period were significantly lower than those in the intervening period for the partial vaccination (p-value 1.2×10^−4^) and the full vaccination (p-value 1.8×10^−6^). The average effectiveness of partial vaccination in the three periods was 23% (SE: 1%) less than that of the full vaccination, which was consistent with the findings in a test-negative case-control study in England (Bernal et al., 2021) and also the sera neutralising capacity study in Planas et al. (2021).

The ten countries used a mixed brand of vaccines. Our results suggested that the estimated VPRs were similar among countries administrated with the same type of vaccines in a period. For example, the estimated VPRs in Germany and Italy were close over the three periods for both partial and full vaccination. In addition to vaccine types, the VPRs were also affected by the distribution strategy of vaccines and the SARS-Cov-2 variants in a country. For the two European nations Italy and Portugal, the differences of the VPRs in the two countries were not significant in the intervening and Delta-dominated periods. The larger difference in the two countries’ VPRs over the Pre-Delta period might be due to the proportions of different types of vaccines administrated and the dosing interval between the first and second shots. Notice that Portugal extended the vaccine dosing interval from 21 to 28 days in February 2021 (Kislaya et al., 2022), and the proportion of mRNA vaccines distributed in Italy was higher as of July 21, 2022 (Statista, 2021), although we did not have data to compare the vaccine distribution of the two countries by April 1, 2021. If information on the numbers of intakes for different vaccine brands was available, we would conduct a regression analysis on the relationship between the estimated VPRs of each country and the intake proportions of different vaccine brands in this country. However, this information was not available in the national level statistics, which prevents us comparing the protection rate across vaccine brands. Although the VPR of a specific brand can not be directly measured due to the lack of data, the country-level results can reflect the protection rates of different vaccine brands to some extent. For instance, results of the US which mainly took Pfizer and Moderna may be used to show the effect of mRNA vaccines; Canada and the four European countries mainly used a mixture of non-replicating viral vector vaccines (AstraZeneca) and mRNA vaccines.

To evaluate the vaccine performance in different countries, we can test for the differences in the VPRs of full dose vaccination among the ten countries over the post-vaccine periods based on the one-sided pairwise test outlined in Section 4.5 with the results reported in Figure 5 (a) and Figures S10-S11 in the SM. The figures show that over the three periods, the US had the significantly highest VPRs among the 10 countries, where the two mRNA-based vaccines Moderna and Pfizer were mainly applied. Turkey where an inactivated vaccine Sinovac was administrated had the significantly lowest VPRs in the intervening and Delta-dominated periods. The VPRs of the countries (Brazil, Canada, Germany, Italy, Peru, Portugal, UK) using AstraZeneca and mRNA vaccines were immediately below the US’s, but higher than those of India which took Covaxin and AstraZeneca. Our results were highly consistent with the analyses of Cai et al. (2021a,b) on the efficacy of COVID-19 vaccines using the published test-negative designs and clinical trials, which found RNA-based vaccines’ effectiveness ranked first, followed by the viral vector vaccines and then the inactivated vaccines.

## 7. Sensitivity analysis

The estimated VPRs depend on assigning values for two tuning parameters *ζ* and *θ*, which took values 5 and 0.8 in our analysis according to external studies. To gain information on the sensitivity of the VPRs on the two tuning parameters, we conducted the sensitivity analysis with *ζ* = 2, 10 under *θ* = 0.8 and *θ* = 0.6 under *ζ* = 5. Estimates for the diagnosis rate and the VPRs in the three post-vaccine periods for all countries are reported in Table S7 in the SM. By comparing with the results at the chosen *ζ* as reported in Tables 2 and S4, the differences between the values of the diagnosis rates and the VPRs in the main analysis and those in sensitivity analyses were quite small. Specifically, the maximum differences in the diagnosis rates and the VPRs by altering the values of *ζ* were 0.03 and 0.085 respectively, and the average absolute differences were 0.013 (SE: 0.002) for the diagnosis rates and 0.021 (SE: 0.002) for the VPRs. For the sensitivity analysis of *θ*, the results in Table S7 show that the differences between the values of the diagnosis rates and the VPRs with *θ* being 0.8 as in the main analysis and those with *θ* being 0.6 were at most 0.025 and 0.095, respectively. And the average absolute differences were 0.014 (SE: 0.002) for the diagnosis rates and 0.022 (SE: 0.003) for the VPRs. These indicated that the estimated diagnosis rates and VPRs were robust to different values of *θ* and *ζ*.

## 8. Scenario analysis

To evaluate community transmission of COVID-19 in the ten countries, we estimate the effective reproduction number *R*_*t*_ via (3) under the vSVIADR model. The estimated *R*_*t*_ curves are displayed in Figure 4 (a), which shows a strong correlation between *R*_*t*_ *>* 1 and the substantial increase of the newly confirmed cases Δ*N*(*t*). Figure 4 (b) shows that the estimated *R*_*t*_ was highly correlated with the estimated infection rates 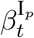, with the generally higher infection rates in the Delta-dominated period as compared to the pre-Delta and intervening periods. For many countries, *R*_*t*_ gradually dropped below 1 after vaccination in the pre-Delta period, but rose above 1 as the Delta variant became prevalent.

**Fig 4:**
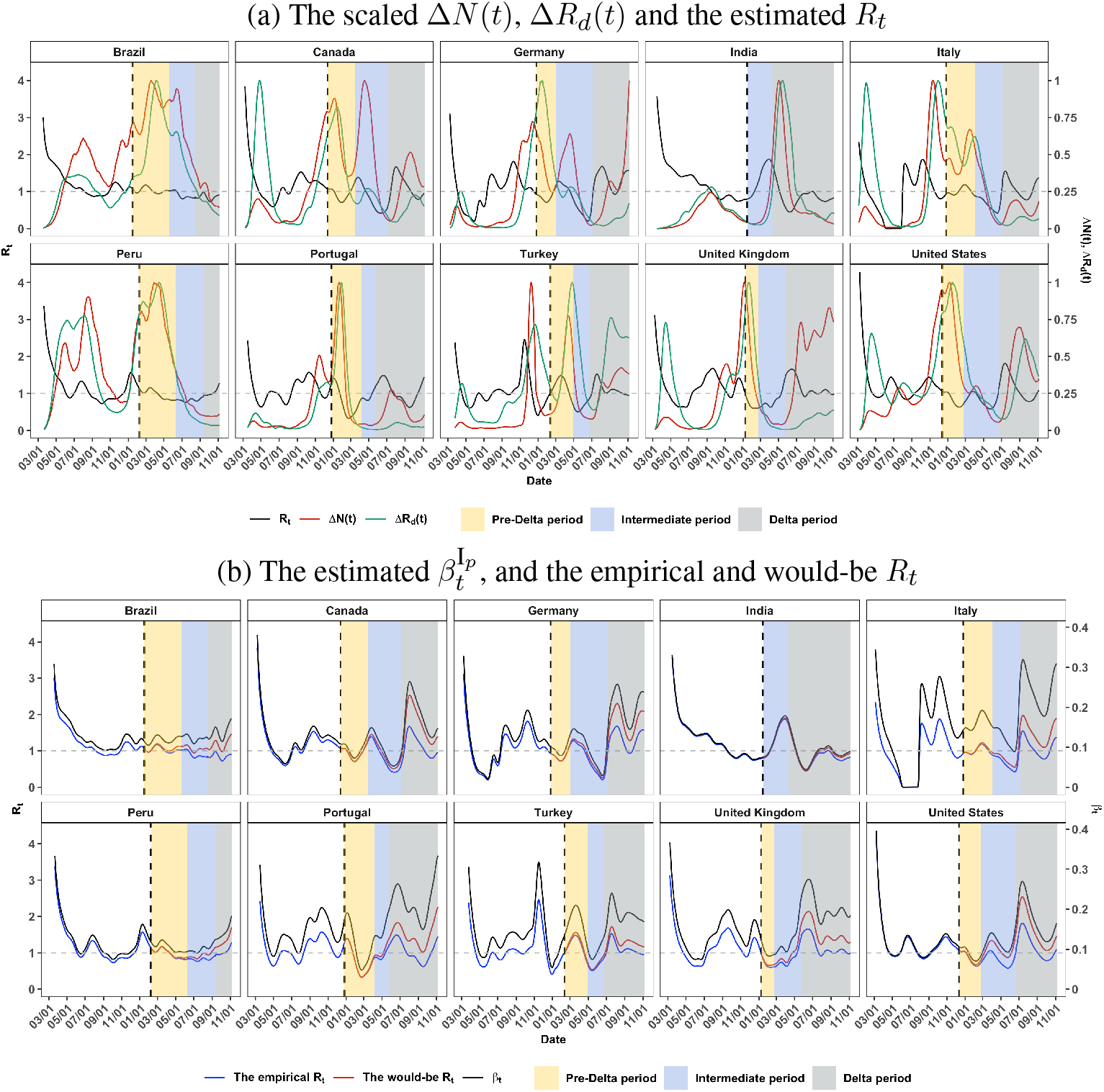
(a) Estimated *R*_*t*_ (black), and the daily increase of confirmed cases Δ*N*(*t*) (red) and death Δ*R*_*d*_(*t*) (green) rescaled to [0, 1] by their respective maximums from March 5, 2020 to November 5, 2021. (b) Estimates of infection rates 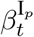, the empirical (blue) and the would-be (red) *R*_*t*_. The yellow, light blue and gray colored areas mark the pre-Delta, intervening and Delta periods, respectively. The dashed vertical lines indicate the vaccination start dates.

**Fig 5:**
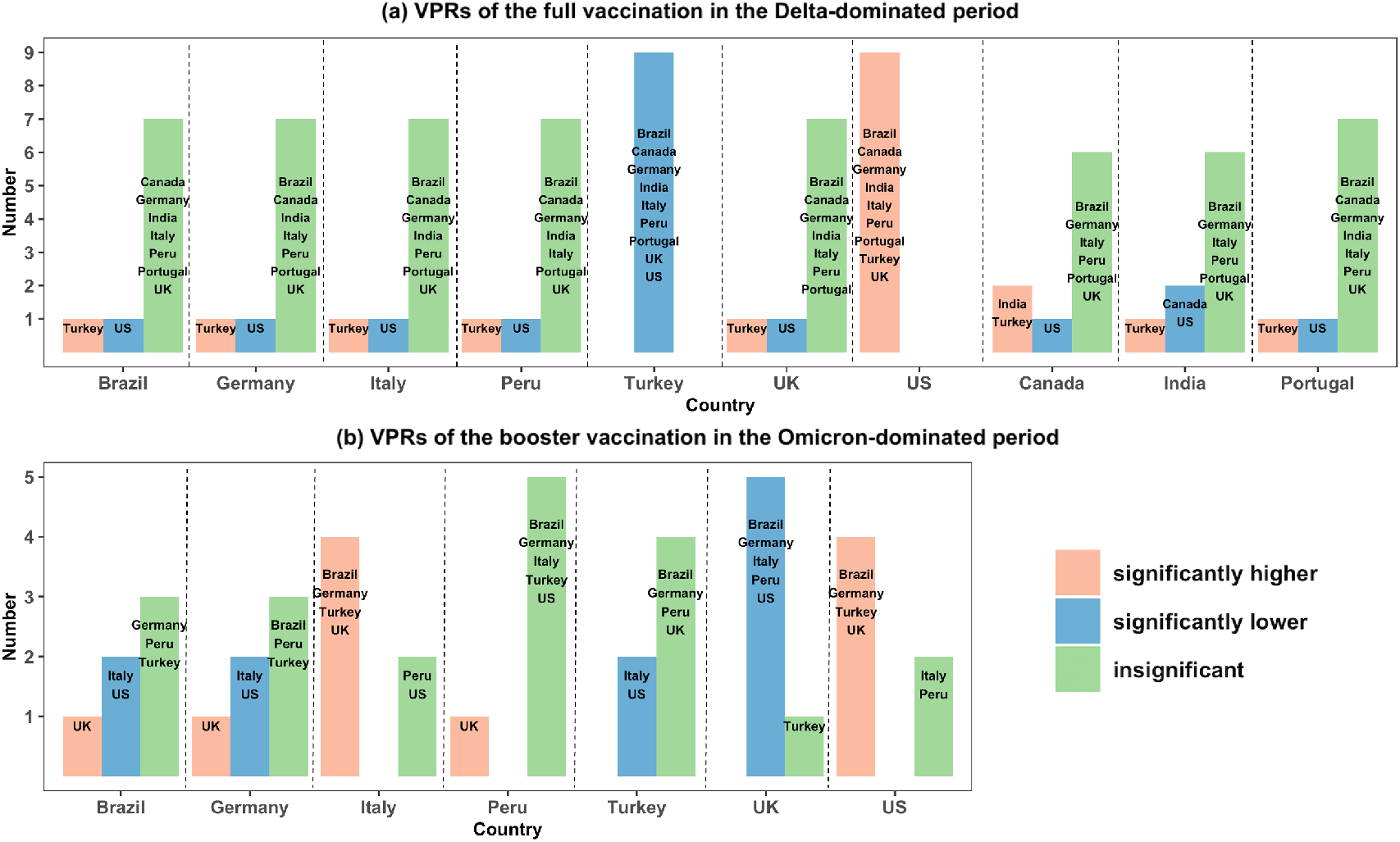
Frequency bar-charts on the pairwise VPR comparison of the country on the horizontal coordinate whose VPR was significantly higher than (red), significantly lower than (blue) or insignificantly different from (green) the other countries by conducting the pairwise testing on the VPRs via the bootstrap method. Panel (a) for the full vaccination among the ten countries in the Delta-dominated period, and Panel (b) for the booster vaccination among the seven countries in the Omicron-dominated period.

The effective reproduction number without vaccines is 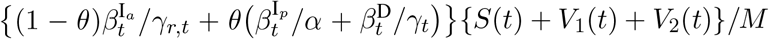. Figure 4 (b) shows that the would-be *R*_*t*_ curves were higher than those of the empirically observed *R*_*t*_ defined in (3) after the start of vaccination, especially when the Delta variant was dominated. This suggests that vaccines contributed to reducing the effective reproduction number *R*_*t*_ substantially in the Delta-dominated period. The average empirically observed *R*_*t*_ in the post-vaccine period for the 10 countries was 20% less than that of the would-be *R*_*t*_ without vaccination.

Another way to evaluate the vaccine effects is to calculate the would-be confirmed cases and deaths under no vaccination at all and the partial vaccination only without going for the full vaccination. For each country, the two scenarios were created by dynamically generating the nine states from the proposed vSVIADR model under each scenario. Since the vaccine effects are accounted by 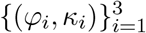, we used the empirically estimated diagnosis rate 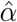, the infection rates 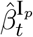, the recovery rates 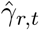 and the death rates 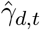 shown in Tables 2 and S4, and Figures 4 (b) and S12. We set 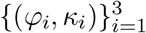 and 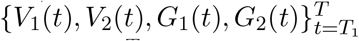 to zero for the no-vaccination scenario, used the observed 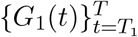 and the estimates 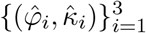 as well as making 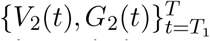 to zero for the partial-vaccination scenario. We also designed a first-dose-priority vaccination scenario which gave priority to the first dose, and then the second dose if there were remaining vaccines left for it. This design can be realized by using the estimates 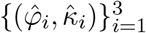 and changing the vaccination functions 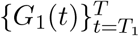 to 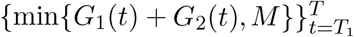 and 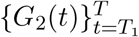 to 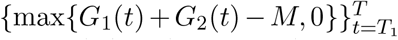. The simulated trajectories under the three scenarios were made based on (2) and (A.3) with the initial values 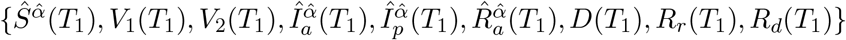. And the simulations of the three scenarios were repeated 100 times for each country under each scenario.

Table 3 provides the would-be increases in confirmed cases and deaths under the no-vaccine, partial-vaccine and first-dose-priority scenarios relative to the observed values. Comparing to the observations up to October 31, 2021, there would be 259 million increase in confirmed cases and 2.6 million increase in deaths for the 10 countries combined in the absence of vaccination, which would amount to 194% and 102% increase over the observed confirmed cases and deaths, respectively. Under the partial-vaccination scenario, there would be 117% and 62% increases in confirmed cases and deaths, respectively. The first-dose-priority scenario would lead to the respective increases of 49% and 24% in the total confirmed cases and deaths, indicating it would not be a better strategy than the existing one used by the countries, although the increases in Brazil, Italy and Peru were not significant. The details on the number of the would-be increases are reported in Table S8 in the SM. The increase in the cases and deaths would have been particularly phenomenal for Canada, Germany and the USA, with 333-769% increases in confirmed cases and 109-376% increases in deaths under the no-vaccine scenario. In a sharp contrast, the would-be increases in both the cases and deaths in India and Peru were rather small. These were due to, as shown in Figure S13, the much lower vaccination rates in Peru (46%) and India (23%), as compared with the other countries.

**Table 3.** The increases (the 95% confidence intervals) and the percentages of increase in the confirmed cases and deaths on October 31, 2021 for the 10 countries under the no (No), partial (Part) and first-dose-priority (First) vaccination scenarios.

| Country | Confirmed cases (thousand) |  |  |  |  | Death (thousand) |  |  |  |  |
| --- | --- | --- | --- | --- | --- | --- | --- | --- | --- | --- |
|  | Increase (95% CI) |  | Percentage % |  |  | Increase (95% CI) |  | Percentage % |  |  |
|  | No |  | No | Part | First | No |  | No | Part | First |
| Brazil | 14915.6 | (14801.6, 15040.1) | 68 | 13 | 0 | 312.4 | (308.4, 315.7) | 51 | 9 | 0 |
| Canada | 13265.8 | (13141.7, 13422.1) | 769 | 411 | 77 | 109.2 | (107.7, 110.9) | 376 | 188 | 34 |
| Germany | 21600.6 | (21363.6, 21833.3) | 465 | 188 | 36 | 104.5 | (102.6, 106.1) | 109 | 37 | 5 |
| India | 6184.0 | (5829.7, 6534.8) | 18 | 6 | 4 | 75.3 | (70.2, 79.8) | 16 | 6 | 3 |
| Italy | 10817744 | (10683.9, 10954.7) | 227 | 61 | 7 | 95.4 | (93.7, 97.0) | 72 | 17 | 0 |
| Peru | 276.9 | (255.3, 295.6) | 13 | 4 | 0 | 10.9 | (9.6, 12.2) | 5 | 2 | 0 |
| Portugal | 2654.8 | (2599.0, 2713.4) | 243 | 88 | 51 | 13.0 | (12.6, 13.4) | 72 | 24 | 11 |
| Turkey | 10647.1 | (10521.0, 10724.6) | 132 | 74 | 34 | 76.1 | (75.1, 77.0) | 108 | 57 | 26 |
| UK | 25284.4 | (25214.1, 25349.7) | 278 | 214 | 134 | 74.7 | (74.1, 75.4) | 53 | 44 | 25 |
| US | 153210.8 | (153132.6, 153294.3) | 333 | 232 | 98 | 1688.6 | (1686.1, 1691.8) | 226 | 166 | 70 |
| Total | 258857.7 | (257542.3, 260162.9) | 194 | 117 | 49 | 2560.1 | (2540.1, 2579.3) | 102 | 62 | 24 |

The actual and the average would-be confirmed cases and deaths generated under the three scenarios are displayed in Figure S14 in the SM, which shows that the gaps between the would-be and the observed grew with respect to time for most countries and scenarios especially in the Delta-dominated period due to the higher would-be *R*_*t*_ in almost all countries. These are another reflection on the vaccines’ effect in reducing the epidemics.

## 9. Extension to booster shot and Omicron period

The vaccination of booster shots started since June 2021 and the circulation of the Omicron variant began since late November 2021. The influence of booster vaccines on the estimated VPRs and the estimation of VPRs in the Omicron era are important for investigating the necessity of booster vaccination for pandemic response. The proposed vSVIADR model can be extended to include an additional compartment *V*_3_(*t*) for those having received the booster shot and not infected at time *t* with an additional vaccine effect parameter to reflect the VPR of the booster vaccination.

A similar estimation approach can be applied for the extended model, in which a trivariate function with respect to the three vaccine effect parameters (partial, full and booster) similar to the criterion function (13) is minimized by the simulation-based estimation method. We extend the analysis to March 15, 2022 to cover the Omicron period. Based on the data quality of the publicly available epidemiological and vaccination data in the extended period, three out of the ten countries were not considered in the empirical analysis due to the missing rates in the vaccination data (48% in Portugal) and insufficient number of confirmed cases (less than 10% of the total population) in India and Canada.

The estimated VPRs with the booster vaccine are provided in Table 4. Compared with Table 2 (c), Table 4 (a) reports the VPRs with the booster vaccine in the Delta-dominated period after the start of booster shots. Notice that, relative to the start of the Delta-dominated period, the booster vaccination started 34 days, 31 days and 8 days later in Brazil, Peru and Turkey, respectively, and more than 51 days later for the other countries. The differences between the estimated VPRs of the partial and full vaccination when Delta was dominated in Tables 2 (c) and 4 (a) were quite small with the average absolute difference in the seven countries being 0.05 (SE: 0.01) for the VPRs of the partial vaccination and 0.03 (SE: 0.006) for the full vaccination. Therefore, the influence of the booster vaccines on the estimated VPRs of full and partial vaccination in Table 2 was very small for the Delta-dominated period. From Table 4 (b) and (c), the Omicron variant reduced the average VPR of the full vaccination in the seven countries to 38% (SE: 2%). The pattern of relative VPRs of full vaccination of those countries in the Omicron-dominated period was similar to that in the Delta-dominated period. Turkey still had the lowest rate at 27%, and the US achieved the highest rate at 43%. The booster dose restored the VPR to 63% (SE: 1.2%) in the Omicron-dominated period, averaged over the seven countries. The VPR of Turkey was much improved by the booster shots, from 27% to 56.2%. These suggested the necessity of receiving booster doses to acquire additional immunity. Testing results for the differences in the VPRs among the seven countries in the same post-vaccine period were shown in Figure 5 (b) and Figures S15-S17 in the SM, which show that the booster VPRs of the US and Italy in the Omicron-dominated period were significantly higher than those of most of the other countries.

**Table 4.**
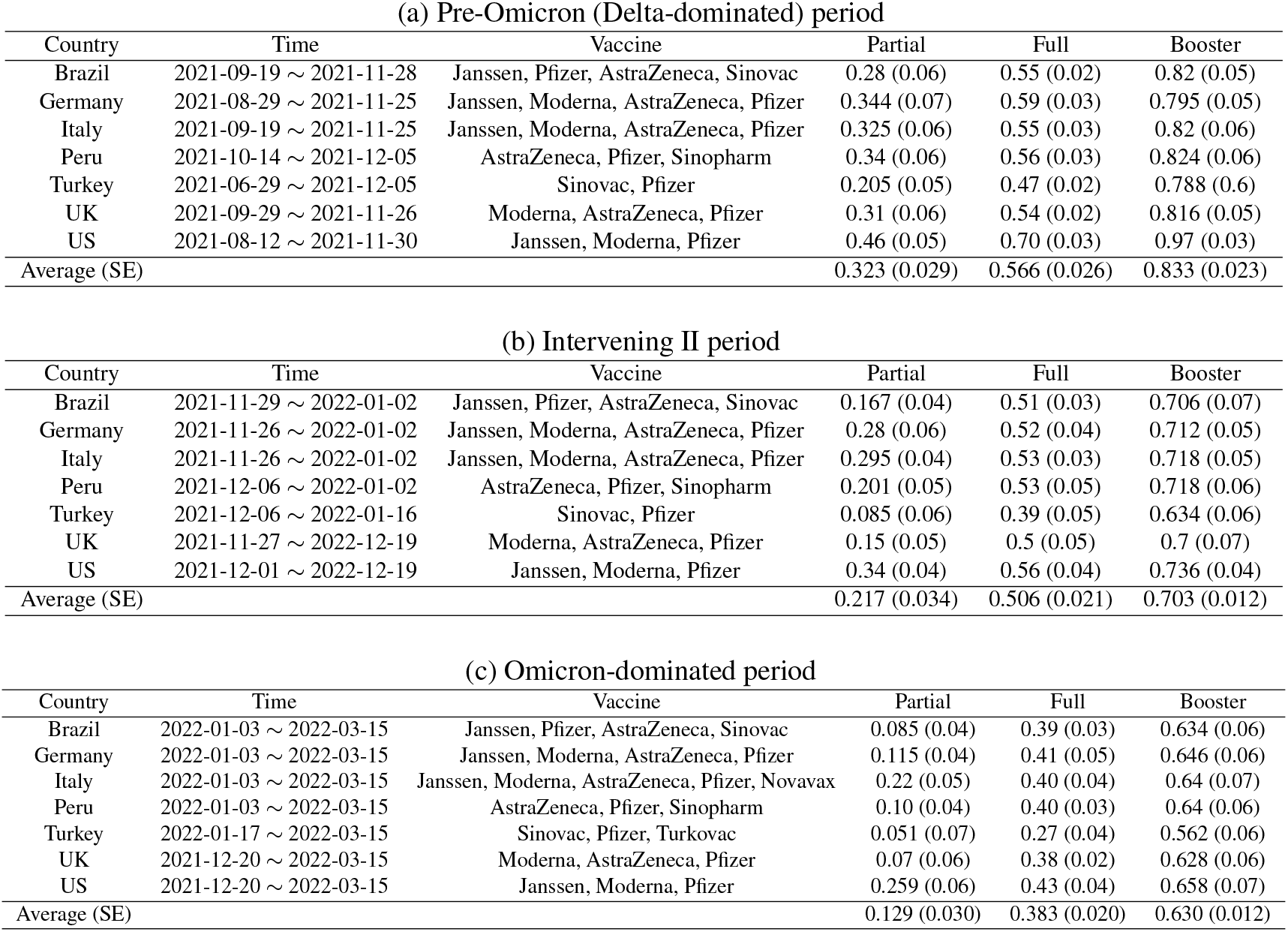
The estimated vaccine protection rates of the partial, full and booster vaccination against COVID-19 infection in the pre-Omicron (Delta-dominated) period from the start of booster shots till the Omicron variant wasfirst detected, the intervening period from the first detection of Omicron till Omicron became predominant and the Omicron-dominated period when the majority of the cases were caused by Omicron for seven countries.

| Country | Time | Vaccine | Partial | Full | Booster |
| --- | --- | --- | --- | --- | --- |
| Brazil | 2021-09-19 ~ 2021-11-28 | Janssen, Pfizer, AstraZeneca, Sinovac | 0.28 (0.06) | 0.55 (0.02) | 0.82 (0.05) |
| Germany | 2021-08-29 ~ 2021-11-25 | Janssen, Moderna, AstraZeneca, Pfizer | 0.344 (0.07) | 0.59 (0.03) | 0.795 (0.05) |
| Italy | 2021-09-19 ~ 2021-11-25 | Janssen, Moderna, AstraZeneca, Pfizer | 0.325 (0.06) | 0.55 (0.03) | 0.82 (0.06) |
| Peru | 2021-10-14 ~ 2021-12-05 | AstraZeneca, Pfizer, Sinopharm | 0.34 (0.06) | 0.56 (0.03) | 0.824 (0.06) |
| Turkey | 2021-06-29 ~ 2021-12-05 | Sinovac, Pfizer | 0.205 (0.05) | 0.47 (0.02) | 0.788 (0.6) |
| UK | 2021-09-29 ~ 2021-11-26 | Moderna, AstraZeneca, Pfizer | 0.31 (0.06) | 0.54 (0.02) | 0.816 (0.05) |
| US | 2021-08-12 ~ 2021-11-30 | Janssen, Moderna, Pfizer | 0.46 (0.05) | 0.70 (0.03) | 0.97 (0.03) |
| Average (SE) |  |  | 0.323 (0.029) | 0.566 (0.026) | 0.833 (0.023) |

(b) Intervening II period
| Country | Time | Vaccine | Partial | Full | Booster |
| --- | --- | --- | --- | --- | --- |
| Brazil | 2021-11-29 ~ 2022-01-02 | Janssen, Pfizer, AstraZeneca, Sinovac | 0.167 (0.04) | 0.51 (0.03) | 0.706 (0.07) |
| Germany | 2021-11-26 ~ 2022-01-02 | Janssen, Moderna, AstraZeneca, Pfizer | 0.28 (0.06) | 0.52 (0.04) | 0.712 (0.05) |
| Italy | 2021-11-26 ~ 2022-01-02 | Janssen, Moderna, AstraZeneca, Pfizer | 0.295 (0.04) | 0.53 (0.03) | 0.718 (0.05) |
| Peru | 2021-12-06 ~ 2022-01-02 | AstraZeneca, Pfizer, Sinopharm | 0.201 (0.05) | 0.53 (0.05) | 0.718 (0.06) |
| Turkey | 2021-12-06 ~ 2022-01-16 | Sinovac, Pfizer | 0.085 (0.06) | 0.39 (0.05) | 0.634 (0.06) |
| UK | 2021-11-27 ~ 2022-12-19 | Moderna, AstraZeneca, Pfizer | 0.15 (0.05) | 0.5 (0.05) | 0.7 (0.07) |
| US | 2021-12-01 ~ 2022-12-19 | Janssen, Moderna, Pfizer | 0.34 (0.04) | 0.56 (0.04) | 0.736 (0.04) |
| Average (SE) |  |  | 0.217 (0.034) | 0.506 (0.021) | 0.703 (0.012) |

(c) Omicron-dominated period
| Country | Time | Vaccine | Partial | Full | Booster |
| --- | --- | --- | --- | --- | --- |
| Brazil | 2022-01-03 ~ 2022-03-15 | Janssen, Pfizer, AstraZeneca, Sinovac | 0.085 (0.04) | 0.39 (0.03) | 0.634 (0.06) |
| Germany | 2022-01-03 ~ 2022-03-15 | Janssen, Moderna, AstraZeneca, Pfizer | 0.115 (0.04) | 0.41 (0.05) | 0.646 (0.06) |
| Italy | 2022-01-03 ~ 2022-03-15 | Janssen, Moderna, AstraZeneca, Pfizer, Novavax | 0.22 (0.05) | 0.40 (0.04) | 0.64 (0.07) |
| Peru | 2022-01-03 ~ 2022-03-15 | AstraZeneca, Pfizer, Sinopharm | 0.10 (0.04) | 0.40 (0.03) | 0.64 (0.06) |
| Turkey | 2022-01-17 ~ 2022-03-15 | Sinovac, Pfizer, Turkovac | 0.051 (0.07) | 0.27 (0.04) | 0.562 (0.06) |
| UK | 2021-12-20 ~ 2022-03-15 | Moderna, AstraZeneca, Pfizer | 0.07 (0.06) | 0.38 (0.02) | 0.628 (0.06) |
| US | 2021-12-20 ~ 2022-03-15 | Janssen, Moderna, Pfizer | 0.259 (0.06) | 0.43 (0.04) | 0.658 (0.07) |
| Average (SE) |  |  | 0.129 (0.030) | 0.383 (0.020) | 0.630 (0.012) |

## 10. Discussion

This paper proposes a stochastic epidemiological model and an estimation procedure to evaluate vaccine protection rates against COVID-19 infection based on publicly available data. The real-world evaluation of vaccine protection is operated under stochasticity, non-permanent immunity and breakthrough infections which were not considered in Dashtbali and Mirzaie (2021) and Giordano et al. (2021). The proposed model and estimation procedure can be applied to study the spread of other infectious diseases.

Our analyses on the 10 countries’ data show significant real-world vaccine benefits in slowing down the COVID-19 infection, largely meeting the WHO standard for 50% VPRs even in the Delta-dominated periods, despite the VPRs waning over time as the Delta variant dominated. Our results on real-world VPR were largely agreeable to those of the published studies via clinical trials and retrospective studies with smaller sample size and higher costs. As demonstrated in the scenario analysis in Section 8, the proposed estimation approach can be used to simulate outcomes under various vaccination strategies, for instance doubling the vaccination rates of the first dose for a more intensive version of the first-dose-priority, in an effort to find better strategies than the one having been carried out by the countries. Our finding of higher effectiveness of the booster vaccination would support efforts for the boost vaccine uptake in the population in the Omicron era.

It is also noted that the imputation of various compartments is only based on the mean specification of Model (2). The Poisson regression in (2) implicitly assumes the means of the daily increments are equal to their variances. If overdispersion exists in the observed data, negative binomial regression with the same mean specification can be used to replace the Poisson assumption in (2). Given the mean *μ*_*t*_, the variance of the negative binomial distribution is equal to 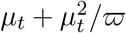 for an overdispersion parameter 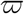. A similar estimation procedure can be carried out to estimate the model parameters by including the additional parameter 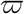 in the objective functions (5) and (12), and simulating the trajectories of the proposed model under the negative binomial distribution.

Furthermore, studying the age and racial effects on the VPRs is an important issue. The proposed model (2) can be extended to include covariates by building parametric regression models on the infection rates and VPRs for countries or states. Specifically, after obtaining the estimated infection rates and VPRs by fitting the epidemiological and vaccination data using the proposed method for each country or state, we estimate the covariate effects by fitting regressions of the estimated infection rates and VPRs on the covariates. Another approach is via constructing a multi-cluster vSVIADR model, where the total population is divided to estimate VPRs for each sub-population, if the daily data on vaccinations and confirmed cases of each sub-population are available. The detailed formulations of the two approaches to studying covariate effects are provided in Section S8 of the SM.

Our study shows the effectiveness of vaccines in reducing the sizes of infection and death. As we do not have access to the data of seriously ill patients, the vaccine impacts on COVID-19 to prevent incidence of serious illness could not be investigated. However, upon having access to these data, the model and analysis can be extended to evaluate such effects. Reinfections are not considered in our study due to the reinfection information in the ten countries is not available in the public domain. This is a limitation of the proposed model. An additional compartment for the reinfection cases may be added to the proposed model. Nevertheless, it is noted that SARS-CoV-2 reinfections were uncommon (less than 1% of the total confirmed infections) until the end of 2021 (Medić et al., 2022) due to the high effectiveness (at approximately 90%) of previous infection in preventing reinfection before the emergence of Omicron variant with good durability as the median time to reinfection was 16 months (Altarawneh et al., 2022; Townsend et al., 2021). Hence, not considering the reinfection would have little effect on the evaluation of the vaccine effects in the pre-omicron eras, which was the main study period of our study. However, the more reinfection cases in the Omicron era would affect the estimation of the vaccine effects against the omicron variant.

## Supporting information

This supplementary material provides the additional details, tables and figures to the main paper.

## Data Availability

All data produced are available online at "https://covid.ourworldindata.org/".

## Funding

The research was partially supported by NSFC Grants 12026607 and 12071013.

SUPPLEMENTARY MATERIAL

## SUPPLEMENTARY MATERIAL

The supplementary material provides additional details, tables and figures.

