## Supplementary material for "Estimating COVID-19 Vaccine Protection Rates via Dynamic Epidemiological Models–A Study of Ten Countries": This supplementary material provides the additional details, tables and figures to the main paper.

This supplementary material provides the additional details, tables and figures to the main paper.

**S1. Data smoothing.** Let $S(t)$, $V_1(t)$, $V_2(t)$, $I_a(t)$, $I_p(t)$, $D(t)$, $R_a(t)$, $R_r(t)$ and $R_d(t)$ be the counts of the uninfected without vaccine immunity, partially vaccine immunized and uninfected, fully vaccine immunized and uninfected, infected but asymptomatic, infected and pre-symptomatic, diagnosed, recovered from asymptomatic, recovered from diagnosed and dead people in a country at day $t$, respectively. Let $R(t)$ be the sum of the recovered $R_r(t)$ and the death $R_d(t)$ as the total removal number at time $t$, and $G_1(t)$ and $G_2(t)$ be the accumulative numbers of people who have received at least one dose of vaccine and who are fully vaccinated, respectively. The reported numbers of vaccinated, infected and removed people are subject to measurement errors. To reduce the error, we apply a weighted moving average filter on the reported counts. Let $N(t) = D(t) + R(t)$ and $\Delta N(t) = N(t+1) - N(t)$ be the accumulative and daily increase numbers of confirmed cases at time $t$. We smooth $\Delta N(t)$ by a boundary kernel $B(t)$ (Jones, 1993) and obtain the smoothed $N(t)$ by summing $\Delta N(s)$ for $s < t$. Let $K(t)$ be the Epanechnikov kernel (Epanechnikov, 1969). The boundary kernel $B(t)$ is constructed as

$$B(t) = \frac{\{a_2(p_t) - a_1(p_t)t(\mathbb{I}_{t\geq T/2} - \mathbb{I}_{t<T/2})\}K(t)}{a_2(p_t)a_0(p_t) - a_1^2(p_t)}, \tag{A.1}$$

where $a_l(p) = \int_p^1 u^l K(u)du$ for $l = 0, 1, 2$ are the moments of the symmetrical kernel $K(t)$, $p_t = \max\{\frac{1-t}{h}, \frac{t-T}{h}, -1\}$ and $h$ denotes the bandwidth. We smooth $\Delta N(t)$ via

$$\Delta\bar{N}(t) = \frac{\sum_{i=1}^{T} N(i)B(\frac{t-i}{h})}{\sum_{i=1}^{T} B(\frac{t-i}{h})} \tag{A.2}$$

for a bandwidth $h = 15$ in our analysis. Note that if $t$ is at least one bandwidth away from $1$ or $T$, $a_0(p) = 1$, $a_1(p) = 0$, and $B(t) = K(t)$. The boundary kernel $B(t)$ corrects the bias of smoothing by the symmetric kernel $K(t)$ at the boundary $t = 1$ and $t = T$. The same boundary kernel $B(t)$ is used for the parameter estimation for the proposed vSVIADR model in equations (4) in the main paper.

Let $\bar{N}(t)$ be the smoothed total infection number by summing $\Delta\bar{N}(s)$ for $s < t$. Applying the same method, we can obtain the smoothed dead $\bar{R}_d(t)$ and recovered numbers $\bar{R}_r(t)$, and smoothed numbers for vaccinated people having received at least one dose $\bar{G}_1(t)$ and full doses $\bar{G}_2(t)$. Let $\bar{R}(t) = \bar{R}_d(t) + \bar{R}_r(t)$ and $\bar{D}(t) = \bar{N}(t) - \bar{R}(t)$ be the smoothed total removal number and active infected number at time $t$, respectively. To simplify the notations, we drop the bar notations, and write $\bar{N}(t)$, $\bar{R}(t)$, $\bar{D}(t)$, $\bar{G}_1(t)$ and $\bar{G}_2(t)$ as $N(t)$, $R(t)$, $D(t)$, $G_1(t)$ and $G_2(t)$ when there is no confusion. All the following analysis is based on the smoothed data.

**S2. Data sources to determine the starting dates for the four periods.** The starting date for the vaccination and the daily vaccine statistics were acquired from Our World in Data, while the dates of the earliest reported cases of the Delta variant for the ten countries were collected in Wikipedia (2021). The timing when the Delta variant began to dominate was defined as the first date when the proportion of the Delta variant in the sequenced SARS-CoV-2 viruses was at least 50%, available in University of Bern (2021). However, as the variant proportions were updated every two weeks, we choose the middle date of the two weeks when the proportion of delta variants first exceeded 50% as the approximated start date. These starting dates in the 10 countries are provided in Table S3.

**S3. The progression of the state variables in the vSVIADR model.** Given the initial values $\{S(1), V_1(1), V_2(1), I_a(1), I_p(1), D(1)\}$ and based on the transitions among different compartments generated by Equation (2) in the main paper, the state variables satisfy the following equations which will be used to generate trajectory in the calculation of the criterion function when we estimate $\alpha$, $\varphi$ and $\kappa$.

$$
\begin{aligned}
S(t+1) &= S(t) - \Delta S_I^-(t) - \Delta G_1(t) + \Delta V_{1,L}^-(t) + \Delta V_{2,L}^-(t),\\
V_1(t+1) &= V_1(t) - \Delta V_{1,I}^-(t) - \Delta V_{1,L}^-(t) + \Delta G_1(t) - \Delta G_2(t),\\
V_2(t+1) &= V_2(t) + \Delta G_2(t) - \Delta V_{2,I}^-(t) - \Delta V_{2,L}^-(t),\\
I_a(t+1) &= I_a(t) + \Delta I_a^+(t) - \Delta R_a(t),\\
I_p(t+1) &= I_p(t) + \Delta S_I^-(t) + \Delta V_{1,I}^-(t) + \Delta V_{2,I}^-(t) - \Delta I_a^+(t) - \Delta N(t),\\
D(t+1) &= D(t) + \Delta N(t) - \Delta R_r(t) - \Delta R_d(t),\\
R_a(t+1) &= R_a(t) + \Delta R_a(t), R_r(t+1) = R_r(t) + \Delta R_r(t), R_d(t+1) = R_d(t) + \Delta R_d(t).
\end{aligned}
\tag{A.3}
$$

It is noted that $\Delta G_1(t)$ ($\Delta G_2(t)$) is the number of unvaccinated (partially vaccinated) people receiving the partial (full) vaccination on day $t$. From (A.3), $\Delta V_1(t)$ is the change in the number of the partially vaccine immunized and uninfected compartment $V_1$ on day $t$, which is made from an inflow of unvaccinated people receiving the partial vaccination $\Delta G_1(t)$ and three outflows of partially vaccine immunized people receiving the full vaccination $\Delta G_2(t)$, being infected $\Delta V_{1,I}^-(t)$ and losing vaccine immunity $\Delta V_{1,L}^-(t)$. And $\Delta V_2(t)$ is the change in the number of the fully vaccine immunized and uninfected compartment $V_2$ on day $t$, which results from an inflow of the partially vaccine immunized people receiving the full vaccination $\Delta G_2(t)$, and two outflows of fully vaccine immunized people being infected $\Delta V_{2,I}^-(t)$ and losing the vaccine immunity $\Delta V_{2,L}^-(t)$. Thus, $\Delta G_1(t) \geq \Delta V_1(t)$ and $\Delta G_2(t) \geq \Delta V_2(t)$. The time-varying rate $\phi_{1,t}$ ($\phi_{2,t}$) of receiving the partial (full) vaccination for the susceptible (the partially vaccinated) people $S(t)$ ($V_1(t)$) can be estimated by the kernel smoothing on $\Delta G_1(t)$ ($\Delta G_2(t)$).

**S4. Derivation of the reproduction number.** A disease free equilibrium (DFE) is

$$
(S, V_1, V_2, I_a, I_p, D, R) = \frac{M}{\phi_{1,0}\phi_{2,0} + \mu_2\phi_{1,0} + \mu_1\mu_2 + \mu_2\phi_{2,0}} (\mu_1\mu_2 + \mu_2\phi_{2,0}, \mu_2\phi_{1,0}, \phi_{1,0}\phi_{2,0}, 0, 0, 0, 0).
$$

The infected states in our models are $(I_a, I_p, D)$. Following the procedure of the computation of $R_0$ by the next generation matrix method (Van den Driessche, 2017), we have

$$F = \begin{bmatrix} (1-\theta)\beta_t^{\mathrm{I}_a} & (1-\theta)\beta_t^{\mathrm{I}_p} & (1-\theta)\beta_t^{\mathrm{D}} \\ \theta\beta_t^{\mathrm{I}_a} & \theta\beta_t^{\mathrm{I}_p} & \theta\beta_t^{\mathrm{D}} \\ 0 & 0 & 0 \end{bmatrix} \left\{ \frac{S(t)}{M} + \varphi\kappa\frac{V_1(t)}{M} + \kappa\frac{V_2(t)}{M} \right\},$$

and

$$V = \begin{bmatrix} \gamma_{r,t} & 0 & 0 \\ 0 & \alpha & 0 \\ 0 & -\alpha & \gamma_t \end{bmatrix}, \quad V^{-1} = \begin{bmatrix} \gamma_{r,t}^{-1} & 0 & 0 \\ 0 & \alpha^{-1} & 0 \\ 0 & \gamma_t^{-1} & \gamma_t^{-1} \end{bmatrix}.$$

Thus the next generation matrix is

$$FV^{-1} = \begin{bmatrix} (1-\theta)\frac{\beta_t^{\mathrm{I}_a}}{\gamma_{r,t}} & (1-\theta)(\frac{\beta_t^{\mathrm{I}_p}}{\alpha} + \frac{\beta_t^{\mathrm{D}}}{\gamma_t}) & (1-\theta)\frac{\beta_t^{\mathrm{D}}}{\gamma_t} \\ \theta\frac{\beta_t^{\mathrm{I}_a}}{\gamma_{r,t}} & \theta(\frac{\beta_t^{\mathrm{I}_p}}{\alpha} + \frac{\beta_t^{\mathrm{D}}}{\gamma_t}) & \theta\frac{\beta_t^{\mathrm{D}}}{\gamma_t} \\ 0 & 0 & 0 \end{bmatrix} \left\{ \frac{S(t)}{M} + \varphi\kappa\frac{V_1(t)}{M} + \kappa\frac{V_2(t)}{M} \right\}.$$

By setting the characteristic polynomial

$$\det(\lambda I - FV^{-1}) = \lambda^2 \Bigg[ \lambda - \left\{ (1-\theta)\frac{\beta_0^{\mathrm{I}_a}}{\gamma_{r,0}} + \theta(\frac{\beta_0^{\mathrm{I}_p}}{\alpha} + \frac{\beta_0^{\mathrm{D}}}{\gamma_0}) \right\} \frac{\mu_1\mu_2 + \mu_2\phi_{2,0} + \varphi\kappa\mu_2\phi_{1,0} + \kappa\phi_{1,0}\phi_{2,0}}{\mu_1\mu_2 + \mu_2\phi_{2,0} + \mu_2\phi_{1,0} + \phi_{1,0}\phi_{2,0}} \Bigg]$$

to be zero, we obtain the eigenvalues and then the basic reproduction number

$$R_0 = \rho(FV^{-1}) = \left\{ (1-\theta)\frac{\beta_0^{\mathrm{I}_a}}{\gamma_{r,0}} + \theta(\frac{\beta_0^{\mathrm{I}_p}}{\alpha} + \frac{\beta_0^{\mathrm{D}}}{\gamma_0}) \right\} \frac{\mu_1\mu_2 + \mu_2\phi_{2,0} + \varphi\kappa\mu_2\phi_{1,0} + \kappa\phi_{1,0}\phi_{2,0}}{\mu_1\mu_2 + \mu_2\phi_{2,0} + \mu_2\phi_{1,0} + \phi_{1,0}\phi_{2,0}},$$

where $\rho$ is the spectral radius. Thus, the effective reproduction number is $R_t = \{(1-\theta)\frac{\beta_t^{\mathrm{I}_a}}{\gamma_{r,t}} + \theta(\frac{\beta_t^{\mathrm{I}_p}}{\alpha} + \frac{\beta_t^{\mathrm{D}}}{\gamma_t})\}\{\frac{S(t)}{M} + \varphi\kappa\frac{V_1(t)}{M} + \kappa\frac{V_2(t)}{M}\}$.

**S5. Identification of parameters.** Identifiability under the stochastic vSVIADR model is quite challenging, which requires the expression of the joint distribution function of $\{S(t)$, $V_1(t)$, $V_2(t)$, $I_a(t)$, $I_p(t)$, $D(t)$, $R_a(t)$, $R_r(t)$, $R_d(t)\}$, and integrating out all the unobserved compartments. Nevertheless, we are able to show that the parameters are identifiable in the system of equations (1) in the paper for the deterministic vSVIADR model under parametric specification for the infection rates $\beta_t^{\mathrm{I}_a}$, $\beta_t^{\mathrm{I}_p}$ and $\beta_t^{\mathrm{D}}$.

Firstly, the removal rates $\gamma_{r,t}$ and $\gamma_{d,t}$ can be identified and estimated since $D(t)$, $R_r(t)$ and $R_d(t)$ are observable. Secondly, as $I_a(t)$ and $I_p(t)$ are unobservable, $\beta_t^{\mathrm{I}_a}$, $\beta_t^{\mathrm{I}_p}$ and $\beta_t^{\mathrm{D}}$ are not identifiable. However, under a specification

(A.4) $$\beta_t^{\mathrm{I}_a} = \beta_t^{\mathrm{I}_p}/\zeta, \quad \beta_t^{\mathrm{D}} = \beta_t^{\mathrm{I}_p}/\zeta$$

for a tuning parameter $\zeta > 0$ and $\beta_t^{\mathrm{I}_p} = f(t, \boldsymbol{\tau})$, where $f(\cdot,\cdot)$ is a function of time $t$ and $d$-dimensional parameter $\boldsymbol{\tau}$, the three infection rates $\beta_t^{\mathrm{I}_a}$, $\beta_t^{\mathrm{I}_p}$ and $\beta_t^{\mathrm{D}}$ can be identified. To illustrate this, we consider the time range before vaccination when $V_1(t) = V_2(t) = 0$, $\phi_{1,t} =$

$\phi_{2,t} = 0$, and the deterministic portion of the vSVIADR model in (1) becomes

$$\text{(A.5)}\qquad \begin{aligned} \Delta S(t) &= -H(t, \boldsymbol{\tau}, \zeta) S(t), \\ \Delta I_a(t) &= (1-\theta) H(t, \boldsymbol{\tau}, \zeta) S(t) - \gamma_{r,t} I_a(t), \\ \Delta I_p(t) &= \theta H(t, \boldsymbol{\tau}, \zeta) S(t) - \alpha I_p(t), \\ \Delta D(t) &= \alpha I_p(t) - \gamma_t D(t), \\ \Delta R_a(t) &= \gamma_{r,t} I_a(t), \\ \Delta R_r(t) &= \gamma_{r,t} D(t) \text{ and } \Delta R_d(t) = \gamma_{d,t} D(t) \end{aligned}$$

where $H(t, \boldsymbol{\tau}, \zeta) = \{\beta_t^{\mathrm{I}_p} I_a(t)/\zeta + \beta_t^{\mathrm{I}_p} I_p(t) + \beta_t^{\mathrm{I}_p} D(t)/\zeta\}/M$ is the total infection loading at $t$. Let $R(t) = R_a(t) + R_r(t) + R_d(t)$ be the total removal. As $S(t) = M - I_a(t) - I_p(t) - D(t) - R(t)$, the second to fifth equations in (A.5) can be written as

$$\text{(A.6)}\qquad \begin{aligned} \Delta I_a(t) &= (1-\theta) H(t, \boldsymbol{\tau}, \zeta)\{M - I_a(t) - I_p(t) - D(t) - R(t)\} - \gamma_{r,t} I_a(t), \\ \Delta I_p(t) &= \theta H(t, \boldsymbol{\tau}, \zeta)\{M - I_a(t) - I_p(t) - D(t) - R(t)\} - \alpha I_p(t), \\ \Delta D(t) &= \alpha I_p(t) - \gamma_t D(t) \text{ and} \\ \Delta R_a(t) &= \gamma_{r,t} I_a(t). \end{aligned}$$

For $t = 1, \ldots, T$, there are $4T$ equations in (A.6). By treating the unobserved compartments $I_a(t)$, $I_p(t)$ and $R_a(t)$ as additional unknown parameters, there are $3(T+1) + 3 + d = 3T + d + 6$ distinct "parameters" in (A.6), where $d$ is the number of parameters in the model for $\beta_t^{\mathrm{I}_p}$. If $T > d + 6$, the number of equations is larger than the number of parameters, and the parameters can be identified under the deterministic model before vaccination.

Similar arguments can be applied to the vSVIADR model after the start of vaccination. From (1) in the paper, there are two additional parameters $\phi$ and $\kappa$ for vaccine effects, and two additional unobservable compartments $V_1(t)$ and $V_2(t)$. Therefore, for $t = T_1 + 1, \ldots, T$, there are $6(T - T_1)$ equations and $5(T - T_1 + 1) + 5 + d = 5(T - T_1) + d + 10$ distinct parameters, where $T_1$ is the start date of vaccination. If $T - T_1 > d + 10$, these parameters can be identified.

Although the parameters are identifiable under the deterministic vSVIADR model in (A.5) and the parametric specification of the infection rates, this does not directly imply the identifiability under the stochastic vSVIADR model. Particularly, from the second and third equations in (A.5), since $\alpha$ and $\gamma_{r,t}$ are typically small, the correlation between $I_a(t)$ and $I_p(t)$ could be quite high, especially for large values of $I_a(t)$ and $I_p(t)$ when the daily infections are high. This can cause collinearity issues in the estimation. Therefore, in the proposed estimation procedure, we fix the values of the daily proportion $\theta$ of the pre-symptomatic cases and the ratio parameter $\zeta$ for the three infection rates, and optimize with respect to the other parameters. This simplifies the optimization algorithm and avoids the collinearity issues. Also note that the parametric models for the infection rates $\beta_t^{\mathrm{I}_a}$, $\beta_t^{\mathrm{I}_p}$ and $\beta_t^{\mathrm{D}}$ are only required for establishing the identifiability. In the estimation, we do not need to know the parametric form $f(\cdot, \cdot)$ for $\beta_t^{\mathrm{I}_p}$. Instead, we use nonparametric methods to estimate the parametric model of $\beta_t^{\mathrm{I}_p}$, which is robust to model specification.

The results of sensitivity analyses with respect to the two pre-specified parameters $\theta$ and $\zeta$ in Table S7 show that the differences between the values of the estimated diagnosis rate $\alpha$ and the estimated VPRs in the sensitivity analyses and those in the main analysis in Table 2 in the main paper and Table S4 were quite small. Specifically, the largest absolute difference in the diagnosis rates by altering the values of $\theta$ or $\zeta$ was 0.03 and the average of the absolute differences was 0.013 (SE: 0.001). And for the estimated VPRs, the largest absolute difference was 0.095 and the average of the absolute differences was 0.022 (SE: 0.001). These results indicated that the estimated diagnosis rates and VPRs were robust to different values of $\theta$ and $\zeta$.

**S6. Optimization algorithm to estimate $\boldsymbol{\alpha}$ in Section 4.2.** In this section, we provide the optimization algorithm for the objective function $f_1(\alpha, \beta_t^{\mathrm{I}_p})$ in Equation (5) with $\beta_t^{\mathrm{I}_p}$ approximated by $\tilde{\beta}_t^{\mathrm{I}_p}(\boldsymbol{\lambda}_1)$ in Equation (9). We first determine a plausible range for the parameter $\boldsymbol{\lambda}_1$ for minimizing $f_1(\alpha, \tilde{\beta}_t^{\mathrm{I}_p}(\boldsymbol{\lambda}_1))$. To this end, we first construct a kernel smoothing estimator for $\beta_t^{\mathrm{I}_p}$ in the pre-vaccine period based on the imputed values $\{\hat{I}_p^\alpha(t), \hat{I}_a^\alpha(t), \hat{S}^\alpha(t)\}$. From the first equation in Equation (6) in the main paper,

$$\{I_p(t+1) + (\alpha-1)I_p(t)\}/\theta \approx \{\beta_t^{\mathrm{I}_a} I_a(t) + \beta_t^{\mathrm{I}_p} I_p(t) + \beta_t^{\mathrm{D}} D(t)\}S(t)/M. \tag{A.7}$$

Let $Y(t) = \{\hat{I}_p^\alpha(t+1) + (\alpha-1)\hat{I}_p^\alpha(t)\}/\theta$ and $X(t) = \left[\hat{I}_p^\alpha(t) + \{D(t) + \hat{I}_a^\alpha(t)\}/r\right]\hat{S}^\alpha(t)/M$. Under the setting $\beta_t^{\mathrm{I}_a} = \beta_t^{\mathrm{D}} = \beta_t^{\mathrm{I}_p}/r$, from Equation (A.7), $\beta_t^{\mathrm{I}_p}$ in the pre-vaccine stage can be estimated by the kernel regression of $Y(t)$ on $X(t)$ as

$$\hat{\beta}_t^{\mathrm{I}_p,\alpha} = \frac{\sum_{i=1}^{T_1} X(i)Y(i)B\{(t-i)/h\}}{\sum_{i=1}^{T_1} X(i)^2 B\{(t-i)/h\}} \tag{A.8}$$

for $t < T_1$, where $B(\cdot)$ is the same boundary kernel used in Equation (4) and $h$ is a bandwidth parameter. This regression estimate of $\beta_t^{\mathrm{I}_p}$ can be interpreted by the dynamics of $I_p(t)$ from the fifth equation in Equation (1). Note that $\alpha\hat{I}_p^\alpha(t)$ is the estimated out-flow from $I_p(t)$ to $D(t)$. Therefore, $\hat{I}_p^\alpha(t+1) - \hat{I}_p^\alpha(t) + \alpha\hat{I}_p^\alpha(t)$ is the estimated daily new pre-symptomatic cases (the inflow to $I_p(t)$). Since $\theta$ proportion of the total new infections are pre-symptomatic in the vSVIADR model, $Y(t)$ approximates the total newly infected cases (asymptomatic and pre-symptomatic combined) at time $t$. Meanwhile, $\hat{I}_p^\alpha(t) + \{D(t) + \hat{I}_a^\alpha(t)\}/r$ represents the estimated size of the infection group at $t$, where $\{D(t) + \hat{I}_a^\alpha(t)\}/r$ is the rescaled infectious stock in the $D$ and $I_a$ compartments in terms of $\beta_t^{\mathrm{I}_p}$. In a well-mixed population before the vaccination, the newly infected cases were caused by the contact of the infectious and susceptible people (estimated by $\hat{S}^\alpha(t)$) with the infection rate $\beta_t^{\mathrm{I}_p}$. Therefore, $\beta_t^{\mathrm{I}_p} X(t)$ can be viewed as the expected value of the newly infected number $Y(t)$.

Regressing $\hat{\beta}_t^{\mathrm{I}_p,\alpha}$ on the B-spline basis functions, we obtain a preliminary estimate of $\boldsymbol{\lambda}_1$

$$\tilde{\boldsymbol{\lambda}}_1^\alpha = (\tilde{\lambda}_0^\alpha, \ldots, \tilde{\lambda}_{n_1+3}^\alpha) = \operatorname*{argmin}_{\boldsymbol{\lambda}_1} \sum_{t \in \mathcal{S}_1} \left[\hat{\beta}_t^{\mathrm{I}_p,\alpha} - \sum_{k=0}^{n_1+3} \lambda_{1,k}\psi_{k,4}\{(t-t_1)/(t_2-t_1)\}\right]^2. \tag{A.9}$$

Let the cartesian product set

$$\Lambda_\alpha = [\tilde{\lambda}_0^\alpha - \delta_1, \tilde{\lambda}_0^\alpha + \delta_1] \times \ldots \times [\tilde{\lambda}_{n_1+3}^\alpha - \delta_1, \tilde{\lambda}_{n_1+3}^\alpha + \delta_1] \tag{A.10}$$

be the candidate set of $\boldsymbol{\lambda}_1$ in the objective function $f_1(\alpha, \tilde{\beta}_t^{\mathrm{I}_p}(\boldsymbol{\lambda}_1))$, where $\delta_1 > 0$ is a search window size. Instead of directly minimizing $f_1(\alpha, \tilde{\beta}_t^{\mathrm{I}_p}(\tilde{\boldsymbol{\lambda}}_1^\alpha))$ with respect to $\alpha$, we consider profile minimization of $f_1(\alpha, \tilde{\beta}_t^{\mathrm{I}_p}(\boldsymbol{\lambda}_1))$ for $\boldsymbol{\lambda}_1 \in \Lambda_\alpha$ at each $\alpha$, where

$$\tilde{f}_1(\alpha) = \min_{\boldsymbol{\lambda}_1 \in \Lambda_\alpha} \frac{1}{|\mathcal{S}_1|} \sum_{t \in \mathcal{S}_1} \left\{\tilde{I}_p(t; \alpha, \boldsymbol{\lambda}_1, \hat{\mathcal{H}}^\alpha(t_1))/\hat{I}_p^\alpha(t) - 1\right\}^2.$$

And then, minimize $\tilde{f}_1(\alpha)$ over $\alpha \in \mathcal{A}$, where $\mathcal{A}$ is a candidate set for $\alpha$. The profile optimization can lead to a better fitting of the objective function $f_1(\alpha, \tilde{\beta}_t^{\mathrm{I}_p}(\boldsymbol{\lambda}_1))$.

In the real data analysis, to minimize $\tilde{f}_1(\alpha)$ with respect to $\alpha \in \mathcal{A}$, we set a grid resolution of 0.005 for the candidate set $\mathcal{A}$ of $\alpha$, and chose the number of internal knots $n_1$ such that the maximum root mean squared relative error (RMSRE)

$$\max_{\alpha\in\mathcal{A}}[\sum_{t\in\mathcal{S}_1}\{\beta_t^{\mathrm{I}_p}(\tilde{\boldsymbol{\lambda}}_1^{\alpha})/\hat{\beta}_t^{\mathrm{I}_p,\alpha}-1\}^2/(\#\{\mathcal{S}_1\})]^{1/2}$$

of the regression (A.9) over $\alpha \in \mathcal{A}$ is less than 0.001 to guarantee enough basis functions to model the unknown infection rate function $\{\beta_t^{\mathrm{I}_p}\}_{t\in\mathcal{S}_1}$. The selected values of $n_1$ for the 10 countries are listed in Table S4 in the SM. Since the maximal coefficients of variation of the estimated coefficients $\{\tilde{\lambda}_k^{\alpha}\}_{k=0}^{n_1+3}$ from (A.9) for the 10 countries were less than 0.003, we set the search window $\delta_1$ for the candidate set $\Lambda_\alpha$ to be 0.02 and a grid resolution of 0.01 for each component of $\Lambda_\alpha$. Implemented in C++ using 100 cores on a server with Intel Xeon Gold 6132 2.6GHz CPUs, the time of calculating $\hat{\alpha}$ for a country based on the average $\{\tilde{I}_p(t;\alpha,\boldsymbol{\lambda}_1,\hat{\mathcal{H}}^{\alpha}(t_1))\}_{t\in\mathcal{S}_1}$ of 300 trajectories for the 30-day time interval $\mathcal{S}_1$ was about 2 hours.

**S7. Optimization algorithm to estimate $\varphi$ and $\kappa$ in Section 4.3.** In this section, we provide the optimization algorithm for the objective function $f_2(\varphi,\kappa,\beta_t^{\mathrm{I}_p})$ in Equation (12) with $\beta_t^{\mathrm{I}_p}$ approximated by $\tilde{\beta}_t^{\mathrm{I}_p}(\boldsymbol{\lambda}_2)$. Similar as the construction of the candidate set $\Lambda_\alpha$ in Section S6, we obtain the candidate set of $\boldsymbol{\lambda}_2$ by considering two sets of regression $\{\hat{\beta}_t^{\mathrm{I}_p,\hat{\alpha}}\}_{t=T_1-l_1}^{T_1}$ before vaccination and $\{\hat{\beta}_{t,*}^{\mathrm{I}_p,\hat{\alpha}}\}_{t=T_1}^{T_1+l_1}$ after vaccination on the B-spline basis functions, and obtaining

$$\tilde{\boldsymbol{\lambda}}^{\text{before}}=(\tilde{\lambda}_0^{\text{before}},\ldots,\tilde{\lambda}_{n_2+3}^{\text{before}})=\operatorname*{argmin}_{\boldsymbol{\lambda}_2}\textstyle\sum_{t=T_1-l_1}^{T_1}\left[\hat{\beta}_t^{\mathrm{I}_p,\hat{\alpha}}-\sum_{k=0}^{n_2+3}\lambda_{2,k}\psi_{k,4}\{(t-T_1+l_1)/l_1\}\right]^2,$$

$$\tilde{\boldsymbol{\lambda}}^{\text{after}}=(\tilde{\lambda}_0^{\text{after}},\ldots,\tilde{\lambda}_{n_2+3}^{\text{after}})=\operatorname*{argmin}_{\boldsymbol{\lambda}_2}\textstyle\sum_{t=T_1}^{T_1+l_1}\left[\hat{\beta}_{t,*}^{\mathrm{I}_p,\hat{\alpha}}-\sum_{k=0}^{n_2+3}\lambda_{2,k}\psi_{k,4}\{(t-T_1)/l_1\}\right]^2,$$

where $\{\hat{\beta}_t^{\mathrm{I}_p,\hat{\alpha}}\}_{t=T_1-l_1}^{T_1}$ is calculated from (A.8) with the estimated diagnosis rate $\hat{\alpha}$, and $\{\hat{\beta}_{t,*}^{\mathrm{I}_p,\hat{\alpha}}\}_{t=T_1}^{T_1+l_1}$ is calculated in the same way via updating the formulas leading to (A.8) for $t > T_1$. Note that $\{\hat{\beta}_{t,*}^{\mathrm{I}_p,\hat{\alpha}}\}_{t=T_1}^{T_1+l_1}$ is not a valid estimator for the infection rate $\{\beta_t^{\mathrm{I}_p}\}_{t=T_1}^{T_1+l_1}$ after the start of vaccination, as the estimator (A.8) is for the pre-vaccine stage without considering the vaccine effects. Our proposal here is to find a reasonable range of $\boldsymbol{\lambda}_2$ for minimizing $f_2(\varphi,\kappa,\beta_t^{\mathrm{I}_p}(\boldsymbol{\lambda}_2))$, and $\{\hat{\beta}_{t,*}^{\mathrm{I}_p,\hat{\alpha}}\}_{t=T_1}^{T_1+l_1}$ serve as a lower bound for $\{\beta_t^{\mathrm{I}_p}\}_{t=T_1}^{T_1+l_1}$ since the vaccine would have reduced the transmission resulting in a lowered than the should-be estimate of $\beta_t^{\mathrm{I}_p}$. In this sense, $\{\hat{\beta}_t^{\mathrm{I}_p,\hat{\alpha}}\}_{t=T_1-l_1}^{T_1}$ and $\{\hat{\beta}_{t,*}^{\mathrm{I}_p,\hat{\alpha}}\}_{t=T_1}^{T_1+l_1}$ provide approximate upper and lower bounds for $\{\beta_t^{\mathrm{I}_p}\}_{t=T_1}^{T_1+l_1}$, and the region between $\tilde{\boldsymbol{\lambda}}^{\text{before}}$ and $\tilde{\boldsymbol{\lambda}}^{\text{after}}$ should contain the true coefficient $\boldsymbol{\lambda}_2$. Like $\Lambda_1$, for a search window $\delta_2 > 0$, let $\Lambda_2$ be the cartesian product of $[\min(\tilde{\lambda}_i^{\text{before}},\tilde{\lambda}_i^{\text{after}})-\delta_2,\max(\tilde{\lambda}_i^{\text{before}},\tilde{\lambda}_i^{\text{after}})+\delta_2]$ for $i=0,\ldots,n_2+3$. Similar as $\delta_1$, we set $\delta_2$ to 0.02 based on the variation of $\tilde{\lambda}_i^{\text{before}}$ and $\tilde{\lambda}_i^{\text{after}}$.

In the real data analysis, we set $l_1 = 50$, and chose the number of internal knots $n_2$ such that the RMSREs of the B-spline fitting of $\boldsymbol{\lambda}_2$ are less than 0.005. The selected $n_2$ of the 10 countries for the pre-Delta, intervening and Delta-dominated periods are listed in Table S4. To compute $\tilde{f}_2(\varphi,\kappa)$ in Equation (13) and minimize it with respect to $\varphi$ and $\kappa$, we set a grid resolution of $0.5\times 0.01$ for $\Theta$ and 0.01 for each component $\lambda_{2,k}$ of $\Lambda_2$ in the grid search algorithm. The grid search algorithm used to optimize (14) to find $(\hat{\varphi},\hat{\kappa})$ took about 8 hours for one country, which could be substantially reduced by applying the genetic algorithm.

**S8. Including covariates in the model.** Studying the age and racial effects on the VPRs is an important issue. The proposed model (2) in the main paper can be extended to include covariates. Let $\beta_{i,t}^{\mathrm{I}_p}$, $\varphi_i$ and $\kappa_i$ be the infection rate of the pre-symptomatic compartment, and the vaccine effect parameters for the $i$th country/state. As in the proposed approach stated in Section 3 in the main paper, we may set $\beta_{i,t}^{\mathrm{I}_a} = \beta_{i,t}^{\mathrm{D}} = \beta_{i,t}^{\mathrm{I}_p}/\zeta$ to reduce the model complexity. Let $\mathbf{X}_i$ denote the demographic profile of the $i$th country/state. We could consider parametric models

$$\beta_{i,t}^{\mathrm{I}_p} = g_\beta(\mathbf{X}_i^{\mathrm{T}}\pi_\beta) + \epsilon_{\beta,i}, \quad \varphi_i = g_\varphi(\mathbf{X}_i^{\mathrm{T}}\pi_\varphi) + \epsilon_{\varphi,i} \quad \text{and}$$

$$\kappa_i = g_\kappa(\mathbf{X}_i^{\mathrm{T}}\pi_\kappa) + \epsilon_{\kappa,i}$$

for the impact of the covariate $\mathbf{X}_i$ on $\beta_{i,t}^{\mathrm{I}_p}$, $\varphi_i$ and $\kappa_i$, where $g_\beta(\cdot)$, $g_\varphi(\cdot)$ and $g_\kappa(\cdot)$ are some known functions, $\epsilon_{\beta,i}$, $\epsilon_{\varphi,i}$ and $\epsilon_{\kappa,i}$ are errors of the models, and $\pi_\beta$, $\pi_\varphi$ and $\pi_\kappa$ represent the covariate effects of interest on the infection rate and vaccine protection rates. Here, the covariates could also include the intake proportions of different vaccine brands (if available), in order to provide comparative protection rates across vaccination brands. The estimated infection rates and VPRs, $\hat{\beta}_{i,t}^{\mathrm{I}_p}$, $\hat{\varphi}_i$ and $\hat{\kappa}_i$, can be obtained by fitting the epidemiological and vaccination data by the proposed method for each country or state. Then, we estimate the covariate effects $\pi_\beta$, $\pi_\varphi$ and $\pi_\kappa$ by fitting regressions of the estimated infection rates and VPRs, $\hat{\beta}_{i,t}^{\mathrm{I}_p}$, $\hat{\varphi}_i$ and $\hat{\kappa}_i$, on the covariates $\mathbf{X}_i$.

Another approach is constructing a multi-cluster vSVIADR model, where the total population is divided into $m$ sub-populations of interest, if the daily data on vaccinations and confirmed cases of each sub-population are available. In that case, the model can be extended to a multi-cluster model to achieve the task of estimating VPRs for each sub-population. Specifically, the epidemic transitions of the states can be described by the following system of equations:

$$\begin{aligned}
&\mathrm{E}\{\Delta S_i(t)|\mathcal{F}_t\} = -H(t,\boldsymbol{\beta}_{i,t})S_i(t) - \phi_{1,i,t}S_i(t) + \mu_{1,i}V_{1,i}(t) + \mu_{2,i}V_{2,i}(t),\\
&\mathrm{E}\{\Delta V_{1,i}(t)|\mathcal{F}_t\} = \phi_{1,i,t}S_i(t) - \mu_{1,i}V_{1,i}(t) - \varphi_i\kappa_i H(t,\boldsymbol{\beta}_{i,t})V_{1,i}(t) - \phi_{2,i,t}V_{1,i}(t),\\
&\mathrm{E}\{\Delta V_{2,i}(t)|\mathcal{F}_t\} = \phi_{2,i,t}V_{1,i}(t) - \mu_{2,i}V_{2,i}(t) - \kappa_i H(t,\boldsymbol{\beta}_{i,t})V_{2,i}(t),\\
&\mathrm{E}\{\Delta I_{a,i}(t)|\mathcal{F}_t\} = (1-\theta_i)H(t,\boldsymbol{\beta}_{i,t})\{S_i(t) + \varphi_i\kappa_i V_{1,i}(t) + \kappa_i V_{2,i}(t)\} - \gamma_{r,i,t}I_{a,i}(t),\\
&\mathrm{E}\{\Delta I_{p,i}(t)|\mathcal{F}_t\} = \theta_i H(t,\boldsymbol{\beta}_{i,t})\{S_i(t) + \varphi_i\kappa_i V_{1,i}(t) + \kappa_i V_{2,i}(t)\} - \alpha_i I_{p,i}(t),\\
&\mathrm{E}\{\Delta D_i(t)|\mathcal{F}_t\} = \alpha_i I_{p,i}(t) - \gamma_{i,t}D_i(t),\ \mathrm{E}\{\Delta R_{a,i}(t)|\mathcal{F}_t\} = \gamma_{r,i,t}I_{a,i}(t),\\
&\mathrm{E}\{\Delta R_{r,i}(t)|\mathcal{F}_t\} = \gamma_{r,i,t}D(t) \text{ and } \mathrm{E}\{\Delta R_{d,i}(t)|\mathcal{F}_t\} = \gamma_{d,i,t}D(t),
\end{aligned}$$

where $\beta_{i,t}^{\mathrm{I}_a} = \beta_{i,t}^{\mathrm{D}} = \beta_{i,t}^{\mathrm{I}_p}/\zeta$ and $H(t,\boldsymbol{\beta}_{i,t}) = \beta_{i,t}^{\mathrm{I}_p}\sum_{j=1}^{m}\{I_{a,j}(t)/\zeta + I_{p,j}(t) + D_j(t)/\zeta\}/M$. Here, the notations with the subscript $i$ inherit the same meaning as those notations of the vSVIADR model for the overall population defined in Section 3 in the main paper, and the subscript $i$ indicates the $i$th sub-population for $i \in \{1,\ldots,m\}$. Then, using the confirmed cases and vaccination data for each group, the infection rates and VPRs of each sub-population can be estimated by the proposed method constructed for the overall population. Note that the parameters of the overall population can be viewed as a weighted average of the corresponding parameters in each sub-population. As the sub-population infection data are not available, we are only able to estimate the VPRs at the population level in this paper.

**S9. Supplemental Tables.**

TABLE S1

*The vaccine efficacy and corresponding 95% confidence interval (in parentheses) against the original SARS-Cov-2 strain obtained in recent studies. A "–" indicates the study did not report the vaccine efficacy for one dose.*

| Vaccine name | Vaccine type | Full dose | VE for one dose (%) | VE for two doses (%) |
|---|---|---|---|---|
| Pfizer-BioNTech (BNT162b2) | mRNA | 2 | 52 (29.5 - 68.4) | 95 (90.3 - 97.6) |
| | | | A multinational, placebo-controlled, observer-blinded trial (Polack et al., 2020) | |
| Moderna (mRNA-1273) | mRNA | 2 | 95.2 (91.2 - 97.4) | 94.1 (89.3 - 96.8) |
| | | | A phase 3 randomized, observer-blinded, placebo-controlled trial (Baden et al., 2021) | |
| Janssen (Ad26.COV2.S) | viral vector | 1 | 66.9 (59.0 - 73.4) | |
| | | | A randomized, double-blind, placebo-controlled, phase 3 trial (Sadoff et al., 2021) | |
| AstraZeneca-Oxford (AZD1222) | viral vector | 2 | 76.0 (59.3 - 85.9) | 66.7 (57.4 - 74.0) |
| | | | Three single-blind randomised controlled trials (Voysey et al., 2021a) | |
| | | | 64.1 ( 50.5 - 73.9) | 62.1 (41.0 - 75.7) |
| | | | Four blinded, randomised, controlled trials (Voysey et al., 2021b) | |
| | | | – | 64.3 (56.1 - 71.0) |
| | | | A double-blind, randomized, placebo-controlled clinical trial (Falsey et al., 2021) | |
| | | | – | 76 (68 - 82) |
| | | | A randomised, double-blind, placebo-controlled Phase III trial (AstraZeneca PLC, 2021) | |
| Sinopharm-Beijing (BBIBP-Corv) | inactivated virus | 2 | 65.5 (52.0 - 75.1) | 78.1 (64.8 - 86.3) |
| | | | A randomized, double-blind, phase 3 trial (Al Kaabi et al., 2021) | |
| Sinovac (CoronaVac) | inactivated virus | 2 | 46.4 (0.4 - 71.2) | 83.5 (65.4 - 92.1) |
| | | | A double-blind, randomised, placebo-controlled phase 3 trial (Tanriover et al., 2021) | |
| | | | 15.5 (14.2 - 16.8) | 65.9 (65.2 - 66.6) |
| | | | A prospective national cohort (Jara et al., 2021) | |
| | | | 57.9 (46.4 - 66.9) | 50.7 (36.0 - 62.0) |
| | | | A randomised, double-blind, placebo-controlled phase 3 clinical trial (Palacios et al., 2021) | |
| Covaxin (BBV152) | inactivated virus | 2 | – | 77.8 (65.2 - 86.4) |
| | | | A double-blind, randomised, phase 3 clinical trial (Ella et al., 2021) | |

TABLE S2

*The vaccine efficacy and corresponding 95% confidence interval (in parentheses) against the Delta strain obtained in recent studies. A "–" indicates the study did not report the vaccine efficacy for one dose. Li et al. (2021) reported the mixed vaccine efficacy of two inactivated SARS-CoV-2 vaccines, Sinopharm-Beijing and Sinovac.*

| Vaccine name | Vaccine type | Full dose | VE for one dose (%) | VE for two doses (%) |
|---|---|---|---|---|
| Pfizer-BioNTech (BNT162b2) | mRNA | 2 | 35.6 (22.7 - 46.4) | 88.0 (85.3 - 90.1) |
| | | | A test-negative casecontrol design in England (Bernal et al., 2021) | |
| | | | – | 52.4 (48.0 - 56.4) |
| | | | An observational study in United States (Nanduri et al., 2021) | |
| | | | – | 79 (75 - 82) |
| | | | A test-negative design in Scotland (Sheikh et al., 2021) | |
| | | | – | 75 (71 - 78) |
| | | | A retrospective cohort study in the USA (Tartof et al., 2021) | |
| | | | 45.3 (22.0 - 61.6) | 51.9 (47.0 - 56.4) |
| | | | A matched test-negative casecontrol study in Qatar (Tang et al., 2021) | |
| | | | 58 (51 - 63) | 82 (79 - 85) |
| | | | A large, community-based survey in the United Kingdom (Pouwels et al., 2021) | |
| Moderna (mRNA-1273) | mRNA | 2 | – | 50.6 (45.0 - 55.7) |
| | | | An observational study in United States (Nanduri et al., 2021) | |
| | | | 73.7 (58.1 - 83.5) | 73.1 (67.5 - 77.8) |
| | | | A matched test-negative casecontrol study in Qatar (Tang et al., 2021) | |
| | | | 77.0 (60.7 - 86.5) | 86.7 (84.3 - 88.7) |
| | | | A test negative case-control study in Southern California (Bruxvoort et al., 2021) | |
| Janssen (Ad26.COV2.S) | viral vector | 1 | 60 | |
| | | | Reported by Dr. Scott Gottlieb, former Food and Drug Administration commissioner (NBC Boston, 2021) | |
| AstraZeneca-Oxford (AZD1222) | viral vector | 2 | 30.0 (24.3 - 35.3) | 67.0 (61.3 - 71.8) |
| | | | A test-negative casecontrol design in England (Bernal et al., 2021) | |
| | | | – | 60 (53 - 66) |
| | | | A test-negative design in Scotland (Sheikh et al., 2021) | |
| | | | 46.2 (31.6 - 57.7) | 63.1 (51.5 - 72.1) |
| | | | A test-negative, case-control study in Faridabad, India (Thiruvengadam et al., 2021) | |
| | | | 43 (31 - 52) | 67 (62 - 71) |
| | | | A large, community-based survey in the United Kingdom (Pouwels et al., 2021) | |
| Sinopharm-Beijing (BBIBP-Corv)<br>Sinovac (CoronaVac) | inactivated virus | 2 | 13.8 (-60.2 - 54.8) | 59.0 (16.0 - 81.6) |
| | | | A test-negative casecontrol study in Guangzhou (Li et al., 2021) | |
| Covaxin (BBV152) | inactivated virus | 2 | – | 65.2 (33.1 - 83.0) |
| | | | A double-blind, randomised, phase 3 clinical trial in India (Ella et al., 2021) | |
| | | | 1 (30 - 25) | 47 (29 - 61) |
| | | | A test-negative, case-control study in India (Desai et al., 2022) | |

TABLE S3

*The start dates of vaccination, the first detection of the Delta variant and the dates when the Delta variant began to dominate in the ten countries. The fourth column was reported as the middle date of the two weeks when the proportion of delta variants in all SARS-CoV-2 virus genome sequences first exceeded 50% reported in the last column with the explicit proportion reported in the parentheses.*

| Country | Start of vaccination | Start of Delta | Date Delta Dominate | Two Weeks when Delta Dominate (Proportion) |
|---|---|---|---|---|
| Brazil | 2021-01-16 | 2021-05-20 | 2021-08-16 | 2021-08-09 ~ 2021-08-23 (65%) |
| Canada | 2020-12-13 | 2021-03-15 | 2021-07-05 | 2021-06-28 ~ 2021-07-12 (51%) |
| Germany | 2020-12-26 | 2021-03-01 | 2021-07-05 | 2021-06-28 ~ 2021-07-12 (80%) |
| India | 2021-01-15 | 2020-10-05 | 2021-04-12 | 2021-04-05 ~ 2021-04-19 (56%) |
| Italy | 2020-12-26 | 2021-04-02 | 2021-07-05 | 2021-06-28 ~ 2021-07-12 (70%) |
| Peru | 2021-02-07 | 2021-06-10 | 2021-09-13 | 2021-09-06 ~ 2021-09-20 (57%) |
| Portugal | 2020-12-26 | 2021-04-05 | 2021-05-24 | 2021-05-17 ~ 2021-05-31 (70%) |
| Turkey | 2021-02-11 | 2021-04-28 | 2021-06-21 | 2021-06-14 ~ 2021-06-28 (51%) |
| United Kingdom | 2021-01-09 | 2021-02-22 | 2021-05-24 | 2021-05-17 ~ 2021-05-31 (73%) |
| United States | 2020-12-12 | 2021-02-23 | 2021-06-21 | 2021-06-14 ~ 2021-06-28 (51%) |

TABLE S4

*The internal knots of the order four B-spline basis functions used for the estimation procedure presented in Section 4 in 10 countries in the pre-vaccine, pre-Delta, intervening and Delta-dominated periods. The sixth column was reported as the estimated diagnosis rates for the 10 countries with the standard errors obtained by the bootstrap method in the parentheses.*

| Country | Pre-vaccine | Pre-Delta | Intervening | Delta dominated | Estimated diagnosis rates |
|---|---|---|---|---|---|
| Brazil | 4 | 1 | 2 | 2 | 0.115 (0.014) |
| Canada | 2 | 2 | 1 | 1 | 0.110 (0.010) |
| Germany | 2 | 2 | 1 | 1 | 0.120 (0.012) |
| India | 2 | | 2 | 1 | 0.100 (0.008) |
| Italy | 2 | 2 | 1 | 1 | 0.200 (0.007) |
| Peru | 2 | 2 | 3 | 3 | 0.110 (0.011) |
| Portugal | 2 | 1 | 1 | 1 | 0.160 (0.012) |
| Turkey | 5 | 1 | 1 | 2 | 0.160 (0.011) |
| United Kingdom | 3 | 1 | 1 | 1 | 0.140 (0.013) |
| United States | 3 | 2 | 1 | 1 | 0.100 (0.003) |

TABLE S5

*Average estimates ($10^3\times$ the standard errors) of the vaccine protection rates $1-\varphi\kappa$ and $1-\kappa$ for the partial and full vaccination respectively, with different choices of $\mathcal{S}_2$ under different parameter settings based on 100 simulations. The true values of the population size $M$, the diagnosis rate $\alpha$, the vaccine protection rates $1-\varphi\kappa$ for the partial vaccination and $1-\kappa$ for the full vaccination are $5\times10^8$, 0.15, 0.75 and 0.9, respectively.*

| $\mathcal{S}_2$ | Constant $\beta_t^{\mathrm{I}_p}$ | | Increasing $\beta_t^{\mathrm{I}_p}$ | | Decreasing $\beta_t^{\mathrm{I}_p}$ | |
|---|---|---|---|---|---|---|
| | $1-\hat{\varphi}\hat{\kappa}$ | $1-\hat{\kappa}$ | $1-\hat{\varphi}\hat{\kappa}$ | $1-\hat{\kappa}$ | $1-\hat{\varphi}\hat{\kappa}$ | $1-\hat{\kappa}$ |
| $[T_1+1, T_1+31]$ | 0.656 (2.4) | 0.859 (1) | 0.666 (2.2) | 0.867 (0.9) | 0.63 (2.5) | 0.846 (1) |
| $[T_1+2, T_1+32]$ | 0.663 (2.3) | 0.857 (1) | 0.672 (2.1) | 0.864 (0.9) | 0.662 (2.3) | 0.853 (1) |
| $[T_1+3, T_1+33]$ | 0.67 (2.2) | 0.857 (1) | 0.671 (2.1) | 0.863 (0.9) | 0.672 (2.2) | 0.861 (1) |
| $[T_1+4, T_1+34]$ | 0.69 (2.1) | 0.866 (0.9) | 0.683 (2) | 0.862 (0.9) | 0.685 (2.1) | 0.874 (0.8) |
| $[T_1+5, T_1+35]$ | 0.691 (1.9) | 0.87 (0.8) | 0.7 (1.6) | 0.872 (0.7) | 0.703 (1.7) | 0.882 (0.6) |
| $[T_1+6, T_1+36]$ | 0.706 (1.6) | 0.877 (0.7) | 0.699 (1.6) | 0.872 (0.7) | 0.704 (1.7) | 0.88 (0.7) |
| $[T_1+7, T_1+37]$ | 0.707 (1.5) | 0.88 (0.7) | 0.701 (1.5) | 0.875 (0.6) | 0.701 (1.7) | 0.877 (0.7) |
| $[T_1+8, T_1+38]$ | 0.707 (1.5) | 0.881 (0.6) | 0.699 (1.5) | 0.875 (0.7) | 0.7 (1.7) | 0.877 (0.7) |
| $[T_1+9, T_1+39]$ | 0.711 (1.5) | 0.882 (0.6) | 0.706 (1.5) | 0.876 (0.6) | 0.697 (1.7) | 0.877 (0.8) |
| $[T_1+10, T_1+40]$ | 0.716 (1.5) | 0.883 (0.6) | 0.71 (1.5) | 0.877 (0.6) | 0.707 (1.6) | 0.883 (0.7) |
| $[T_1+11, T_1+41]$ | 0.722 (1.4) | 0.883 (0.6) | 0.716 (1.4) | 0.88 (0.6) | 0.728 (1.2) | 0.891 (0.6) |
| $[T_1+12, T_1+42]$ | 0.731 (1.2) | 0.892 (0.5) | 0.717 (1.4) | 0.885 (0.5) | 0.729 (1.2) | 0.895 (0.5) |
| $[T_1+13, T_1+43]$ | 0.732 (1.2) | 0.891 (0.5) | 0.724 (1.2) | 0.892 (0.4) | 0.729 (1.2) | 0.894 (0.5) |
| $[T_1+14, T_1+44]$ | 0.739 (1) | 0.896 (0.4) | 0.731 (1) | 0.898 (0.4) | 0.731 (1.2) | 0.897 (0.4) |
| $[T_1+15, T_1+45]$ | 0.743 (0.9) | 0.899 (0.3) | 0.733 (1) | 0.901 (0.3) | 0.74 (0.9) | 0.903 (0.3) |
| $[T_1+16, T_1+46]$ | 0.745 (0.8) | 0.901 (0.3) | 0.737 (0.9) | 0.902 (0.3) | 0.743 (0.8) | 0.904 (0.3) |
| $[T_1+17, T_1+47]$ | 0.744 (0.8) | 0.903 (0.3) | 0.737 (0.8) | 0.902 (0.3) | 0.743 (0.8) | 0.903 (0.3) |
| $[T_1+18, T_1+48]$ | 0.744 (0.8) | 0.904 (0.3) | 0.737 (0.8) | 0.903 (0.3) | 0.743 (0.8) | 0.903 (0.3) |
| $[T_1+19, T_1+49]$ | 0.743 (0.8) | 0.904 (0.3) | 0.737 (0.8) | 0.901 (0.3) | 0.742 (0.8) | 0.904 (0.3) |
| $[T_1+20, T_1+50]$ | 0.743 (0.8) | 0.904 (0.3) | 0.737 (0.8) | 0.901 (0.3) | 0.741 (0.8) | 0.902 (0.3) |

TABLE S6

*Average estimates ($10^3\times$ the standard errors) of the diagnosis rate $\alpha$, the vaccine protection rates $1-\varphi\kappa$ and $1-\kappa$ for the partial and full vaccination respectively, under different parameter settings based on 100 simulations. The initial proportions of each compartment in the total population ($V_1(1)/M$, $V_2(1)/M$, $I_a(1)/M$, $I_p(1)/M$, $D(1)/M$, $R_a(1)/M$, $R_r(1)/M$, $R_d(1)/M$) are (0, 0, $8\times10^{-8}$, $4.5\times10^{-7}$, $4\times10^{-8}$, 0, $6\times10^{-8}$, 0).*

| $(\alpha, 1-\varphi\kappa, 1-\kappa)$ | $\beta_t^{\mathrm{I}_p}$ | Population size | $\hat{\alpha}$ | $1-\hat{\varphi}\hat{\kappa}$ | $1-\hat{\kappa}$ |
|---|---|---|---|---|---|
| (0.15, 0.75, 0.9) | Constant | $5\times10^8$ | 0.149 (0.7) | 0.740 (8.3) | 0.901 (3.7) |
| | | $1\times10^9$ | 0.150 (0.2) | 0.750 (1.6) | 0.901 (2.6) |
| | | $1.5\times10^9$ | 0.150 (0.1) | 0.746 (1.1) | 0.906 (2.4) |
| | Increasing | $5\times10^8$ | 0.146 (1.2) | 0.697 (17.7) | 0.890 (4.1) |
| | | $1\times10^9$ | 0.149 (0.8) | 0.725 (10.2) | 0.911 (2.9) |
| | | $1.5\times10^9$ | 0.149 (0.6) | 0.741 (6.1) | 0.910 (2.7) |
| | Decreasing | $5\times10^8$ | 0.145 (1.3) | 0.717 (17.9) | 0.888 (4.7) |
| | | $1\times10^9$ | 0.149 (0.5) | 0.745 (6.3) | 0.900 (2.5) |
| | | $1.5\times10^9$ | 0.150 (0.1) | 0.751 (1.0) | 0.897 (2.0) |
| (0.15, 0.4, 0.6) | Constant | $5\times10^8$ | 0.145 (1.4) | 0.406 (9.5) | 0.623 (4.8) |
| | | $1\times10^9$ | 0.150 (0.3) | 0.400 (1.6) | 0.606 (2.1) |
| | | $1.5\times10^9$ | 0.150 (0.3) | 0.400 (1.4) | 0.603 (1.6) |
| | Increasing | $5\times10^8$ | 0.148 (0.7) | 0.402 (3.2) | 0.617 (3.1) |
| | | $1\times10^9$ | 0.149 (0.6) | 0.399 (3.1) | 0.612 (3.1) |
| | | $1.5\times10^9$ | 0.150 (0.1) | 0.397 (1.1) | 0.606 (2.2) |
| | Decreasing | $5\times10^8$ | 0.145 (1.3) | 0.404 (8.8) | 0.618 (4.0) |
| | | $1\times10^9$ | 0.149 (0.6) | 0.392 (5.6) | 0.605 (1.8) |
| | | $1.5\times10^9$ | 0.150 (0.1) | 0.400 (0.2) | 0.601 (0.9) |

TABLE S7

*The estimated diagnosis rates, and protection rates of the partial and full vaccination in the pre-Delta, the intervening and the Delta dominated periods for the 10 countries in the sensitivity analyses with $\theta = 0.6$, $\zeta = 2$ and $\zeta = 10$.*

| Country | Diagnosis rate | | | Period | Partial vaccination | | | Full vaccination | | |
|---|---|---|---|---|---|---|---|---|---|---|
| | $\theta = 0.6$ | $\theta = 0.8$ | $\theta = 0.8$ | | $\theta = 0.6$ | $\theta = 0.8$ | $\theta = 0.8$ | $\theta = 0.6$ | $\theta = 0.8$ | $\theta = 0.8$ |
| | $\zeta = 5$ | $\zeta = 2$ | $\zeta = 10$ | | $\zeta = 5$ | $\zeta = 2$ | $\zeta = 10$ | $\zeta = 5$ | $\zeta = 2$ | $\zeta = 10$ |
| Brazil | 0.105 | 0.13 | 0.115 | Pre-Delta | 0.67 | 0.56 | 0.54 | 0.78 | 0.78 | 0.77 |
| | | | | Intervening | 0.52 | 0.52 | 0.505 | 0.68 | 0.68 | 0.67 |
| | | | | Delta dominated | 0.40 | 0.37 | 0.355 | 0.60 | 0.58 | 0.57 |
| Canada | 0.120 | 0.130 | 0.120 | Pre-Delta | 0.545 | 0.64 | 0.60 | 0.87 | 0.88 | 0.84 |
| | | | | Intervening | 0.4 | 0.45 | 0.45 | 0.76 | 0.78 | 0.78 |
| | | | | Delta dominated | 0.24 | 0.28 | 0.26 | 0.62 | 0.64 | 0.63 |
| Germany | 0.125 | 0.120 | 0.130 | Pre-Delta | 0.615 | 0.58 | 0.55 | 0.89 | 0.88 | 0.91 |
| | | | | Intervening | 0.49 | 0.55 | 0.535 | 0.66 | 0.70 | 0.69 |
| | | | | Delta dominated | 0.415 | 0.385 | 0.40 | 0.61 | 0.59 | 0.60 |
| India | 0.125 | 0.115 | 0.115 | Intervening | 0.475 | 0.445 | 0.475 | 0.65 | 0.63 | 0.65 |
| | | | | Delta dominated | 0.28 | 0.28 | 0.28 | 0.52 | 0.52 | 0.52 |
| Italy | 0.195 | 0.17 | 0.190 | Pre-Delta | 0.55 | 0.64 | 0.65 | 0.90 | 0.91 | 0.93 |
| | | | | Intervening | 0.535 | 0.595 | 0.565 | 0.69 | 0.73 | 0.71 |
| | | | | Delta dominated | 0.355 | 0.385 | 0.40 | 0.57 | 0.59 | 0.60 |
| Peru | 0.125 | 0.140 | 0.125 | Pre-Delta | 0.685 | 0.685 | 0.7 | 0.79 | 0.79 | 0.8 |
| | | | | Intervening | 0.48 | 0.48 | 0.48 | 0.74 | 0.74 | 0.74 |
| | | | | Delta dominated | 0.46 | 0.415 | 0.415 | 0.64 | 0.61 | 0.61 |
| Portugal | 0.140 | 0.175 | 0.155 | Pre-Delta | 0.505 | 0.55 | 0.535 | 0.67 | 0.70 | 0.69 |
| | | | | Intervening | 0.475 | 0.38 | 0.36 | 0.65 | 0.69 | 0.68 |
| | | | | Delta dominated | 0.385 | 0.34 | 0.34 | 0.59 | 0.56 | 0.56 |
| Turkey | 0.180 | 0.160 | 0.140 | Pre-Delta | 0.44 | 0.44 | 0.44 | 0.72 | 0.72 | 0.72 |
| | | | | Intervening | 0.295 | 0.22 | 0.205 | 0.53 | 0.48 | 0.47 |
| | | | | Delta dominated | 0.16 | 0.145 | 0.175 | 0.44 | 0.43 | 0.45 |
| UK | 0.160 | 0.135 | 0.125 | Pre-Delta | 0.49 | 0.505 | 0.49 | 0.66 | 0.67 | 0.66 |
| | | | | Intervening | 0.46 | 0.415 | 0.43 | 0.64 | 0.61 | 0.62 |
| | | | | Delta dominated | 0.34 | 0.295 | 0.31 | 0.56 | 0.53 | 0.54 |
| US | 0.110 | 0.110 | 0.110 | Pre-Delta | 0.525 | 0.51 | 0.525 | 0.95 | 0.93 | 0.95 |
| | | | | Intervening | 0.64 | 0.675 | 0.65 | 0.82 | 0.87 | 0.86 |
| | | | | Delta dominated | 0.48 | 0.46 | 0.44 | 0.74 | 0.73 | 0.72 |

TABLE S8

*Increases (the 95% confidence intervals) in total confirmed cases and deaths on October 31, 2021 for the 10 countries under the partial (Part) and first-dose-priority (First) scenarios.*

| Country | Confirmed cases (thousand) | | Death (thousand) | |
|---|---|---|---|---|
| | Part | First | Part | First |
| Brazil | 2771.7 (2704.5, 2853.7) | 84.5 (-11.8, 173) | 56.1 (53.3, 58.5) | -2.8 (-5.5, 0) |
| Canada | 7079.7 (6889.1, 7345.5) | 1323.3 (1240.9, 1395.7) | 54.5 (52.9, 56.8) | 9.8 (9.2, 10.5) |
| Germany | 8736.4 (8462.2, 9010.6) | 1691.7 (1552.8, 1795.1) | 35.6 (34.0, 37.3) | 5.1 (4.1, 6.0) |
| India | 2116.5 (1856.1, 2407.6) | 1319.4 (1035.9, 1538.7) | 25.6 (21.9, 29.8) | 16.0 (12.4, 19.1) |
| Italy | 2919.0 (2826.2, 3026.0) | 335.0 (283.9, 379.6) | 22.4 (21.3, 23.6) | 0.3 (-0.5, 1.1) |
| Peru | 85.6 (70.1, 102.5) | 5.5 (-8.5, 20.7) | 3.4 (2.2, 4.9) | 0.2 (-1.1, 1.2) |
| Portugal | 955.3 (886.7, 1025.7) | 553.8 (493.6, 607.6) | 4.3 (3.8, 4.7) | 1.9 (1.6, 2.3) |
| Turkey | 5940.1 (5819.7, 6058.6) | 2750.6 (2656.7, 2848.7) | 40.0 (39.0, 40.9) | 18.5 (17.6, 19.6) |
| UK | 19449.9 (19348.8, 19548.6) | 12143.0 (11938.5, 12345.5) | 61.4 (60.8, 62.0) | 35.1 (34.3, 36) |
| US | 106799.9 (106588.4, 106976.5) | 45079.9 (44743.5, 45461) | 1238.5 (1235.3, 1241.7) | 520.2 (515.4, 525.1) |
| Total | 156854.2 (155451.8, 158355.4) | 65286.7 (63925.2, 66565.6) | 1541.7 (1524.3, 1560.2) | 604.3 (587.6, 620.9) |

### S10. Supplemental Figures.

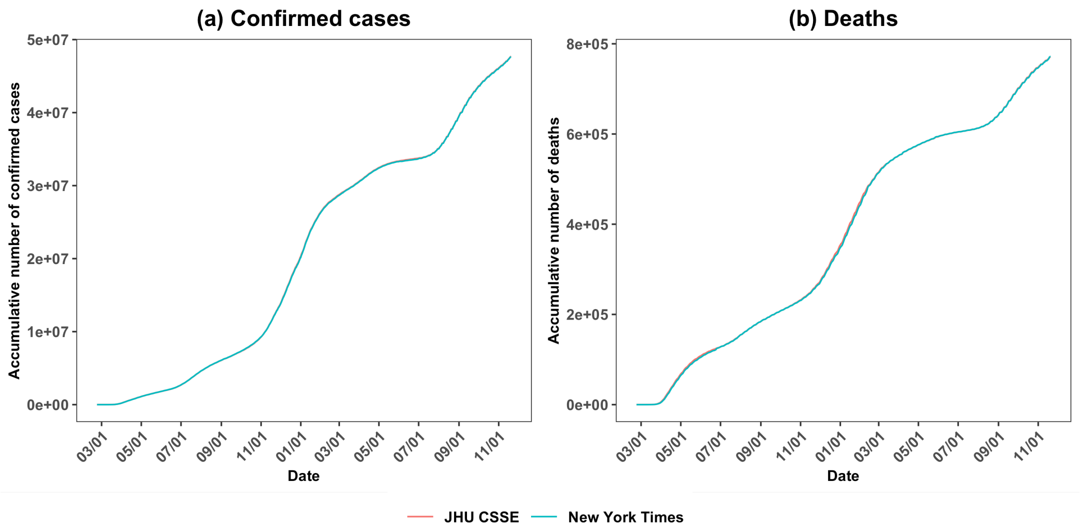


Fig S1: Curves of the accumulative number of confirmed cases and deaths from February 23, 2020 to November 20, 2021 in the US from the JHU CSSE (red) and New York Times (blue) data resources.

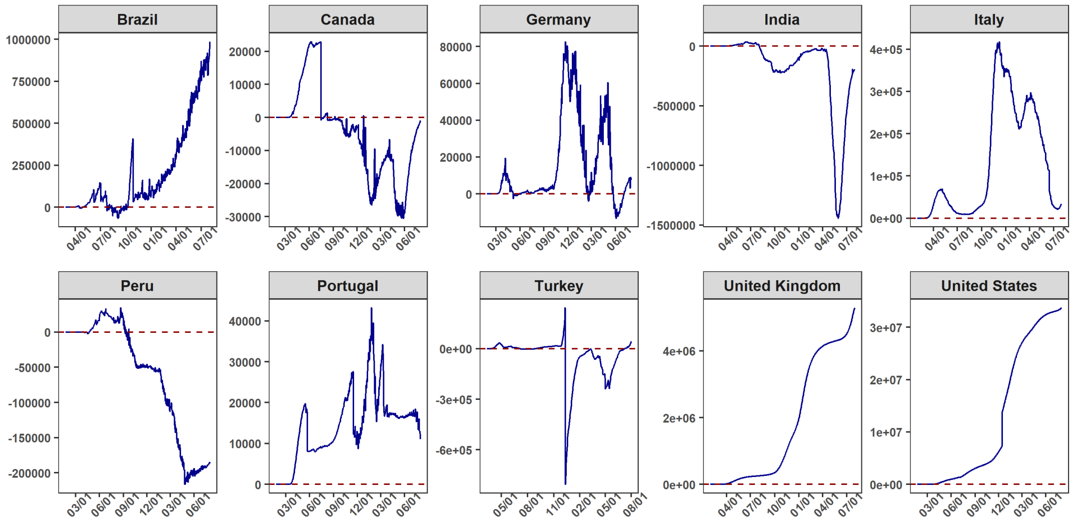


Fig S2: Curves of the imputed recovered cases minus reported recovered cases for the 10 countries from February 23, 2020 to August 4, 2021, before the date when the recovered cases were no longer reported.

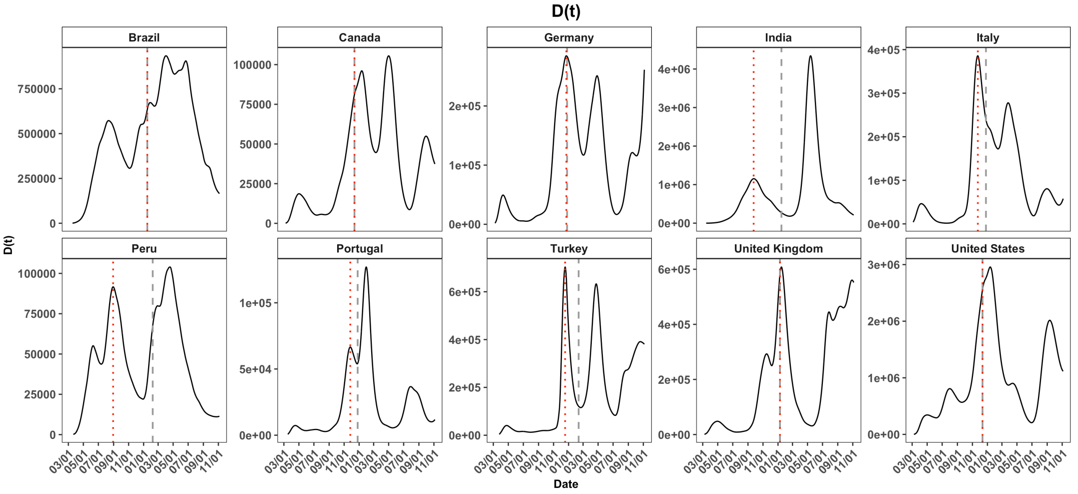


Fig S3: Curves of the diagnosed cases $D(t)$ for the 10 countries in the study period from February 23, 2020 to November 20, 2021 with the peak of the diagnosed cases in the pre-vaccine period (dotted red) and the start date of the vaccination (dashed gray), respectively.

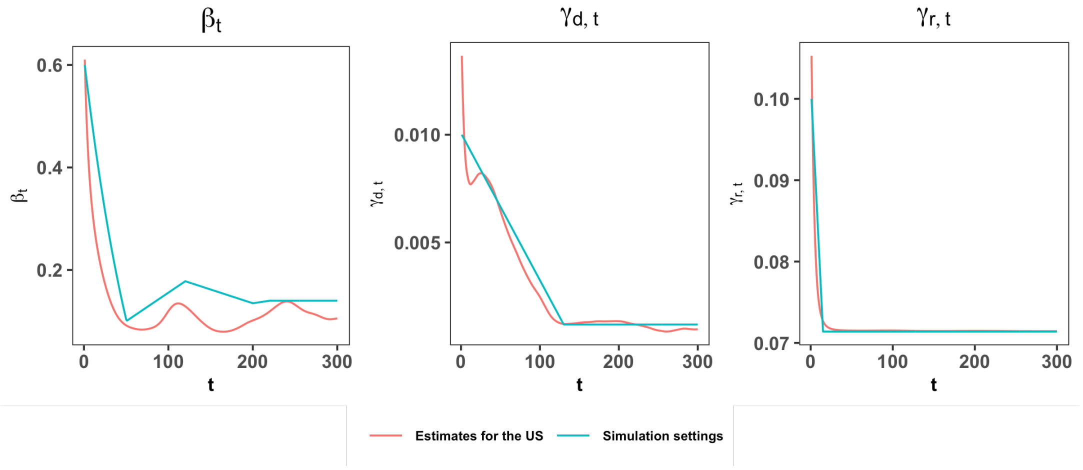


Fig S4: Curves for our simulation setting (blue) of the time-varying parameters $\beta_t^{\mathrm{I}_p}$, $\gamma_{d,t}$ and $\gamma_{r,t}$, and those for the estimates in the US (red).

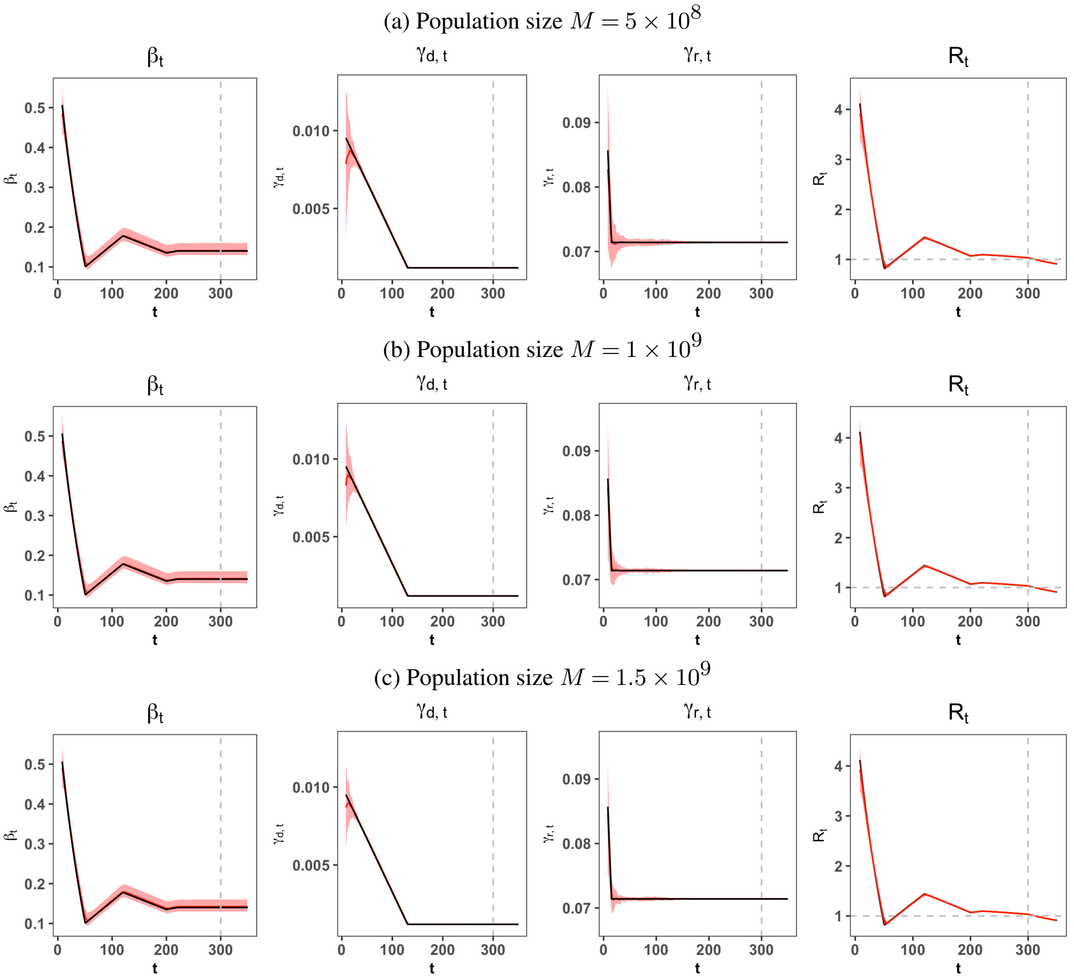


Fig S5: Curves of true (black) and estimated (red) coefficients of $\beta_t^{\mathrm{I}_p}, \gamma_{r,t}, \gamma_{d,t}$ and $R_t$ with the colored 2.5%-97.5% quantile bands for three population sizes. The gray dashed vertical line represents the start of the vaccination and the gray dashed horizontal line represents the critical value 1. The true values of infection rates after the start of the vaccination were $\beta_t^{\mathrm{I}_p} \equiv 0.14$. The true values of $\alpha$, $\varphi$ and $\kappa$ are 0.15, 1.5 and 0.4, respectively.

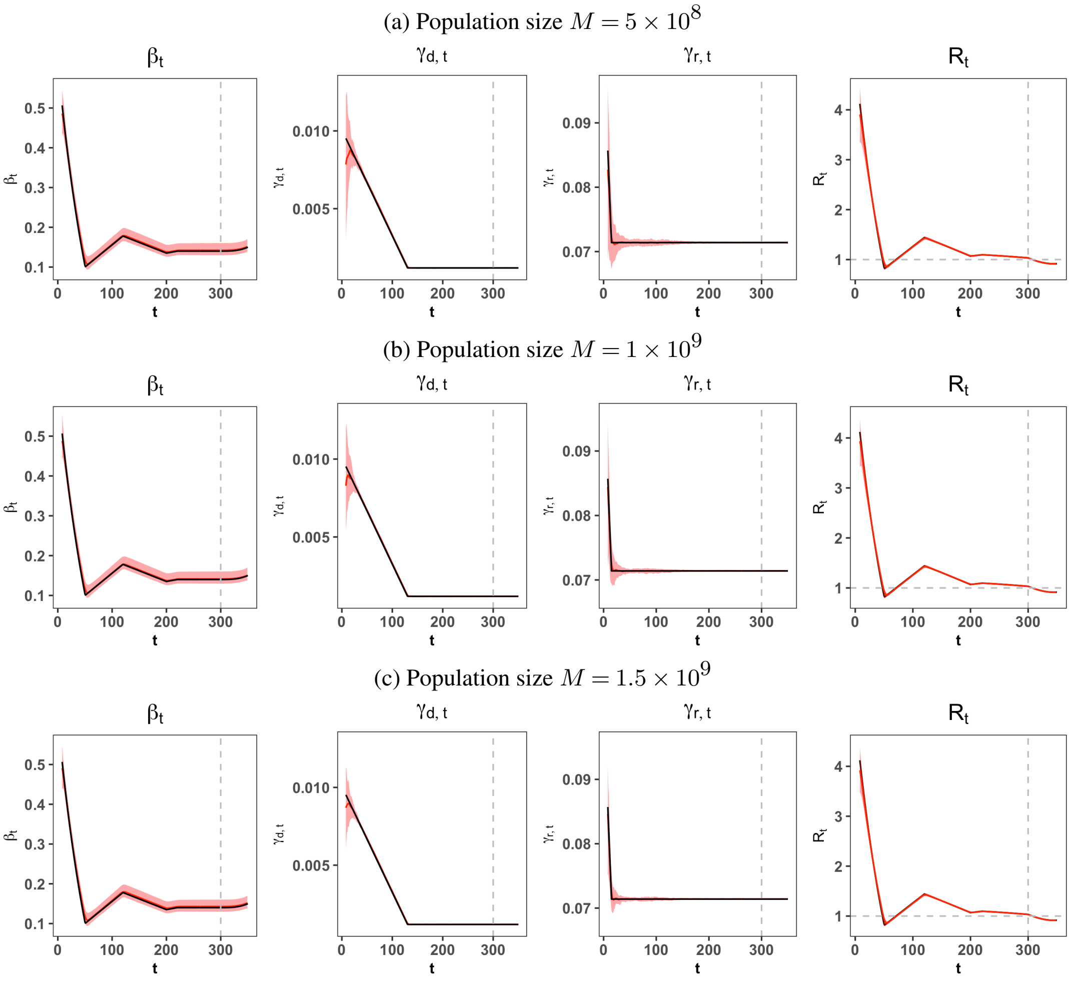


Fig S6: Curves of true (black) and estimated (red) coefficients of $\beta_t^{\mathrm{I}_p}, \gamma_{r,t}, \gamma_{d,t}$ and $R_t$ with the colored 2.5%-97.5% quantile bands for three population sizes. The gray dashed vertical line represents the start of the vaccination and the gray dashed horizontal line represents the critical value 1. The true values of infection rates after the start of the vaccination were $\beta_t^{\mathrm{I}_p} = 0.14 + 8 \times 10^{-8}(t - T_1)^3$. The true values of $\alpha$, $\varphi$ and $\kappa$ are 0.15, 2.5 and 0.1, respectively.

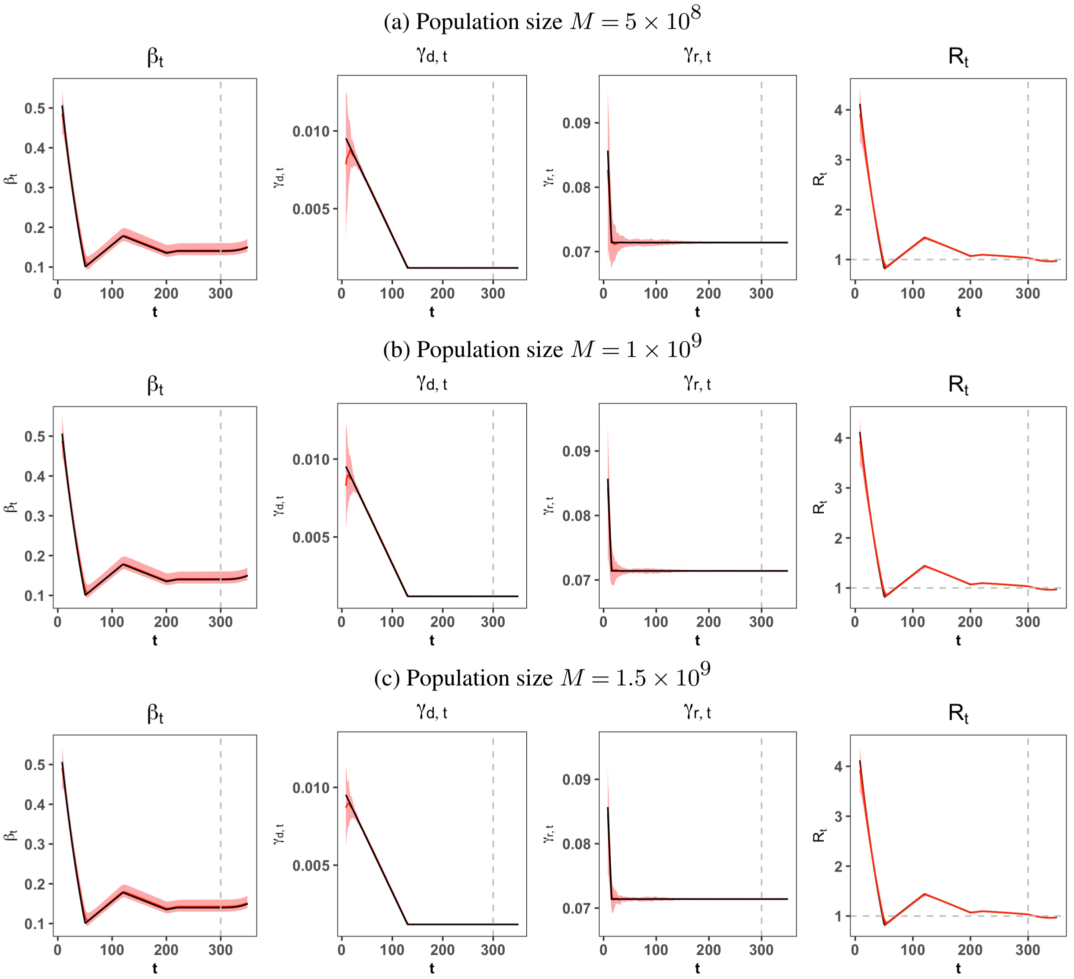


Fig S7: Curves of true (black) and estimated (red) coefficients of $\beta_t^{\mathrm{I}_p}, \gamma_{r,t}, \gamma_{d,t}$ and $R_t$ with the colored 2.5%-97.5% quantile bands for three population sizes. The gray dashed vertical line represents the start of the vaccination and the gray dashed horizontal line represents the critical value 1. The true values of infection rates after the start of the vaccination were $\beta_t^{\mathrm{I}_p} = 0.14 + 8 \times 10^{-8}(t - T_1)^3$. The true values of $\alpha$, $\varphi$ and $\kappa$ are 0.15, 1.5 and 0.4, respectively.

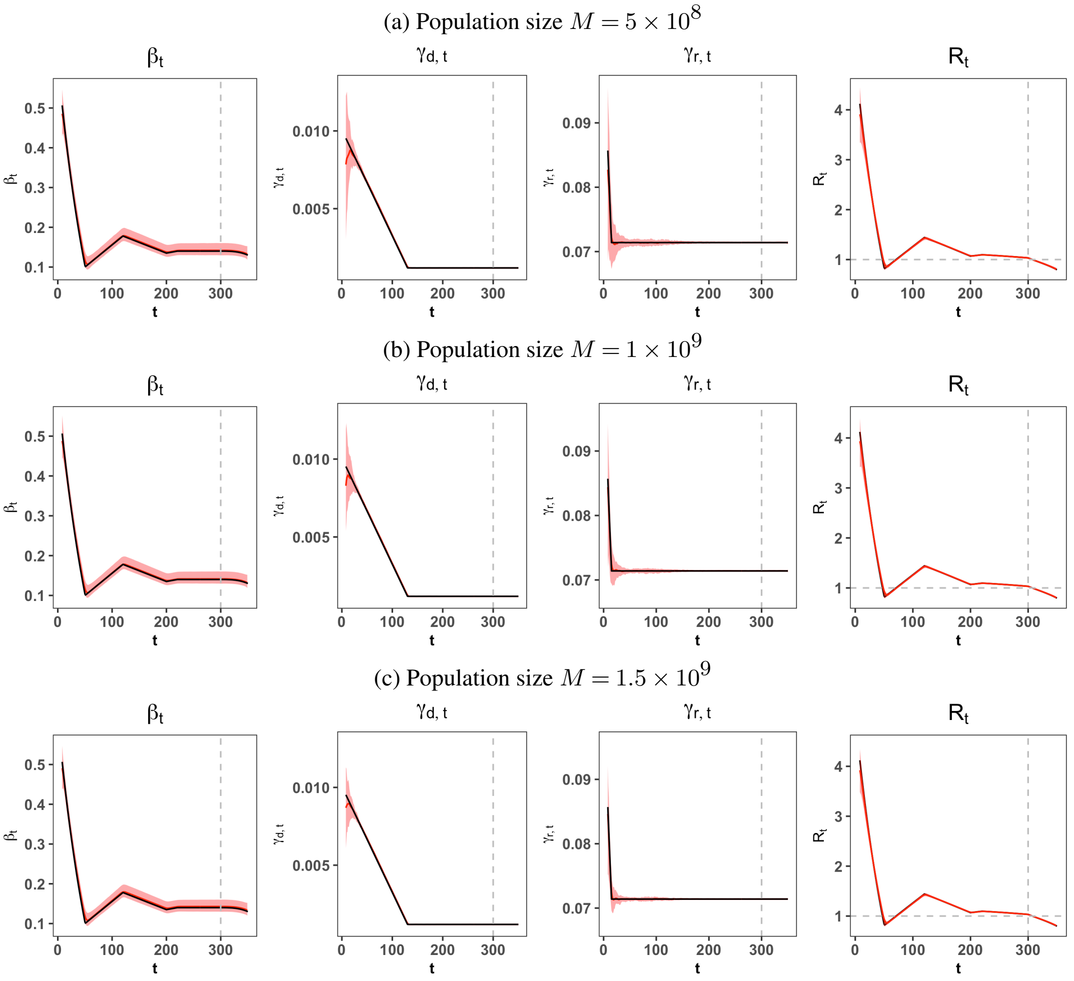


Fig S8: Curves of true (black) and estimated (red) coefficients of $\beta_t^{\mathrm{I}_p}, \gamma_{r,t}, \gamma_{d,t}$ and $R_t$ with the colored 2.5%-97.5% quantile bands for three population sizes. The gray dashed vertical line represents the start of the vaccination and the gray dashed horizontal line represents the critical value 1. The true values of infection rates after the start of the vaccination were $\beta_t^{\mathrm{I}_p} = 0.14 - 8 \times 10^{-8}(t - T_1)^3$. The true values of $\alpha$, $\varphi$ and $\kappa$ are 0.15, 2.5 and 0.1, respectively.

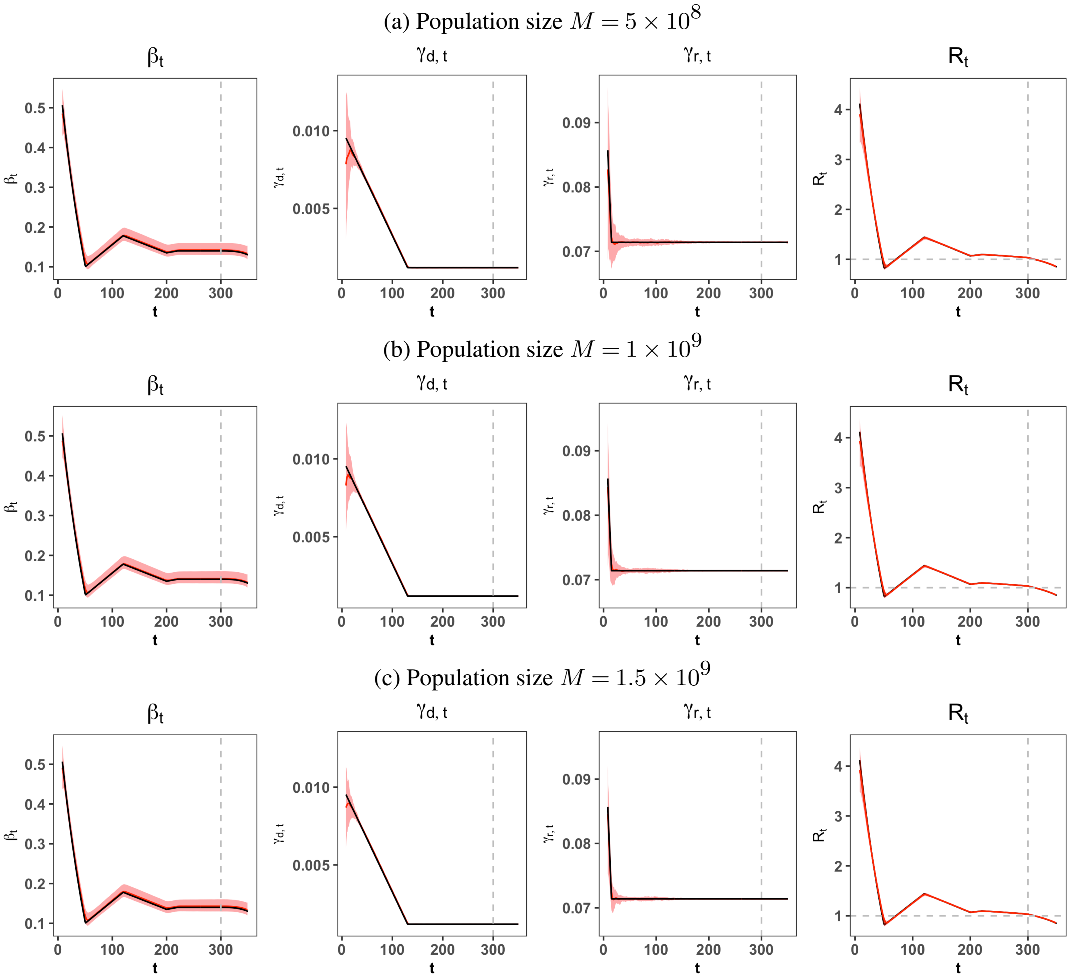


Fig S9: Curves of true (black) and estimated (red) coefficients of $\beta_t^{\mathrm{I}_p}, \gamma_{r,t}, \gamma_{d,t}$ and $R_t$ with the colored 2.5%-97.5% quantile bands for three population sizes. The gray dashed vertical line represents the start of the vaccination and the gray dashed horizontal line represents the critical value 1. The true values of infection rates after the start of the vaccination were $\beta_t^{\mathrm{I}_p} = 0.14 - 8 \times 10^{-8}(t - T_1)^3$. The true values of $\alpha$, $\varphi$ and $\kappa$ are 0.15, 1.5 and 0.4, respectively.

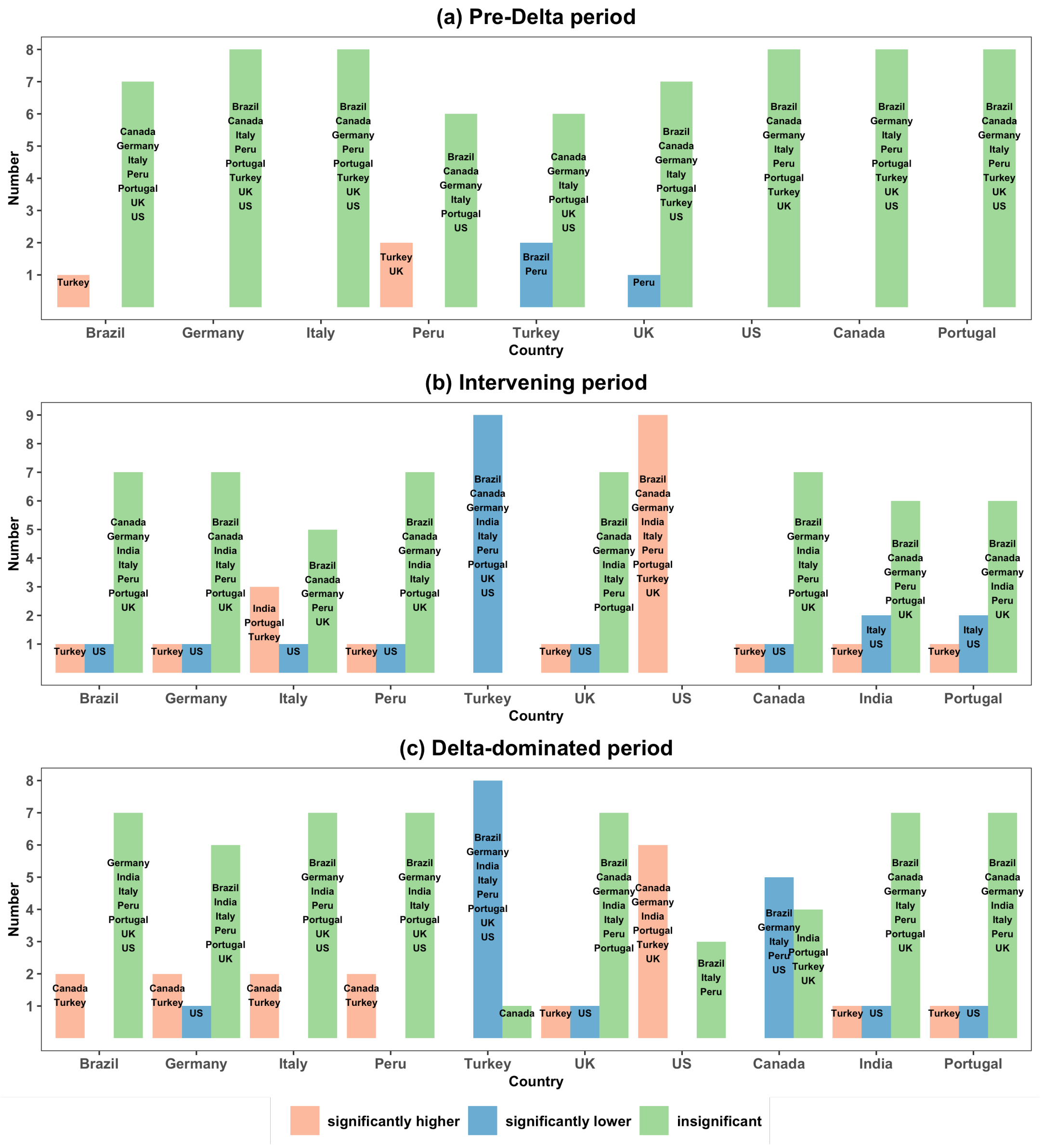


Fig S10: Frequency bar-charts on the pairwise VPR comparison of the country on the horizontal coordinate whose VPR of the partial vaccination was significantly higher than (red), significantly lower than (blue) or insignificantly different from (green) the other countries by conducting the pairwise testing on the VPRs via the bootstrap method among ten countries in the pre-Delta (a), intervening (b) and Delta-dominated (c) periods.

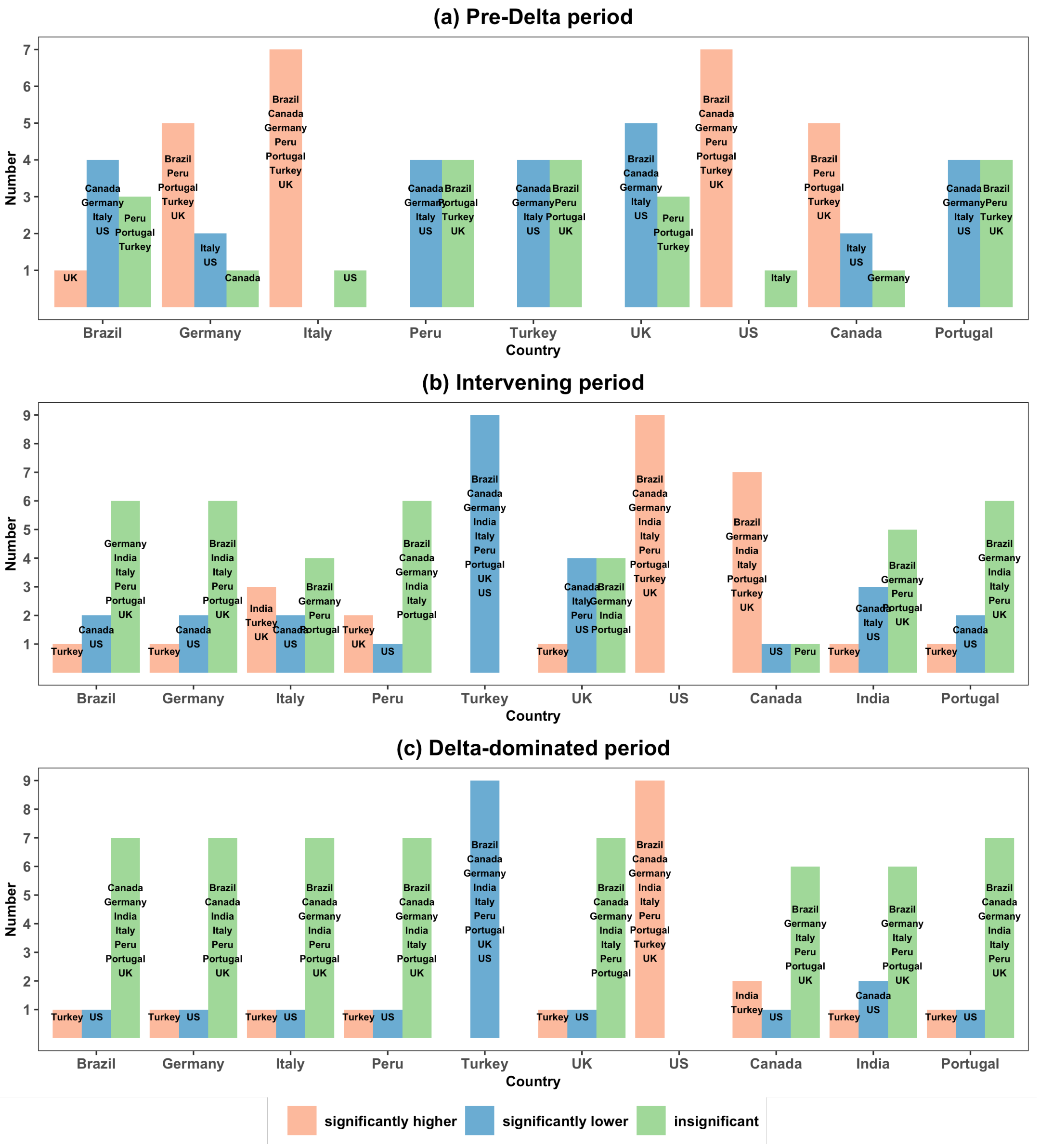


Fig S11: Frequency bar-charts on the pairwise VPR comparison of the country on the horizontal coordinate whose VPR of the full vaccination was significantly higher than (red), significantly lower than (blue) or insignificantly different from (green) the other countries by conducting the pairwise testing on the VPRs via the bootstrap method among ten countries in the pre-Delta (a), intervening (b) and Delta-dominated (c) periods.

(a) Recovery rates $\gamma_{r,t}$

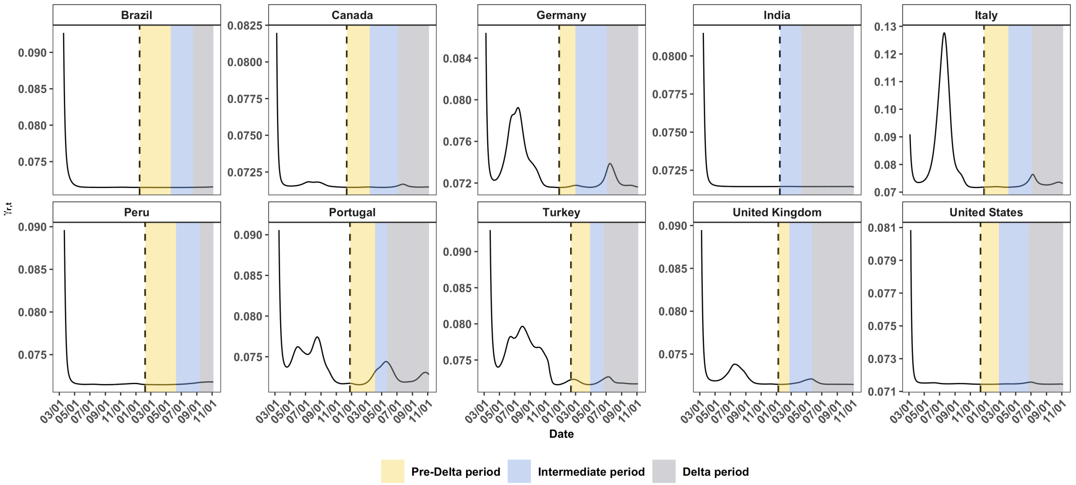


(b) Death rates $\gamma_{d,t}$

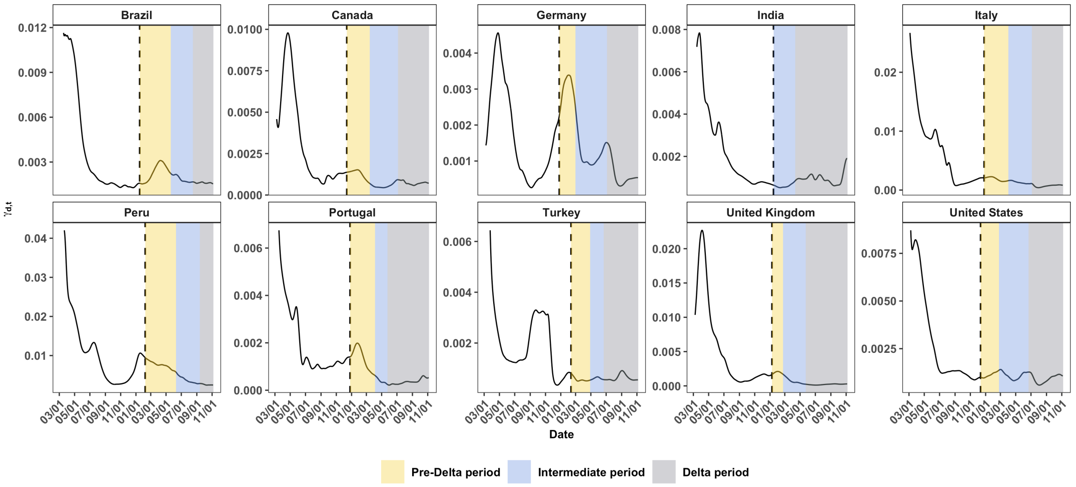


Fig S12: Curves of the estimated (a) recovery rates $\gamma_{r,t}$ and (b) death rates $\gamma_{d,t}$. The yellow, light blue and gray colored areas mark the pre-Delta, intervening and Delta periods, respectively. The black dashed vertical line represents the start of the vaccination.

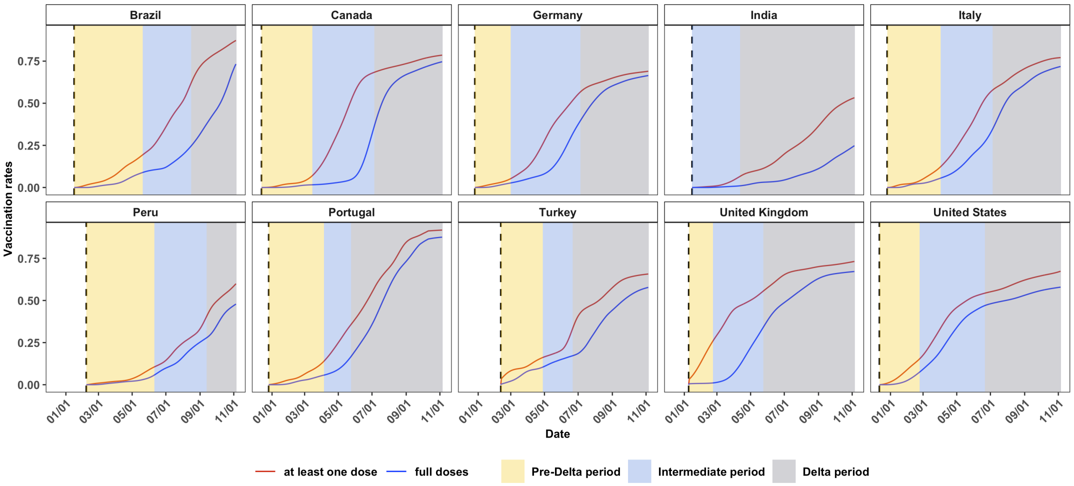


Fig S13: Curves for the rates of people receiving at least one dose $G_1(t)/M$ and full doses of the vaccines $G_2(t)/M$ in the 10 countries.

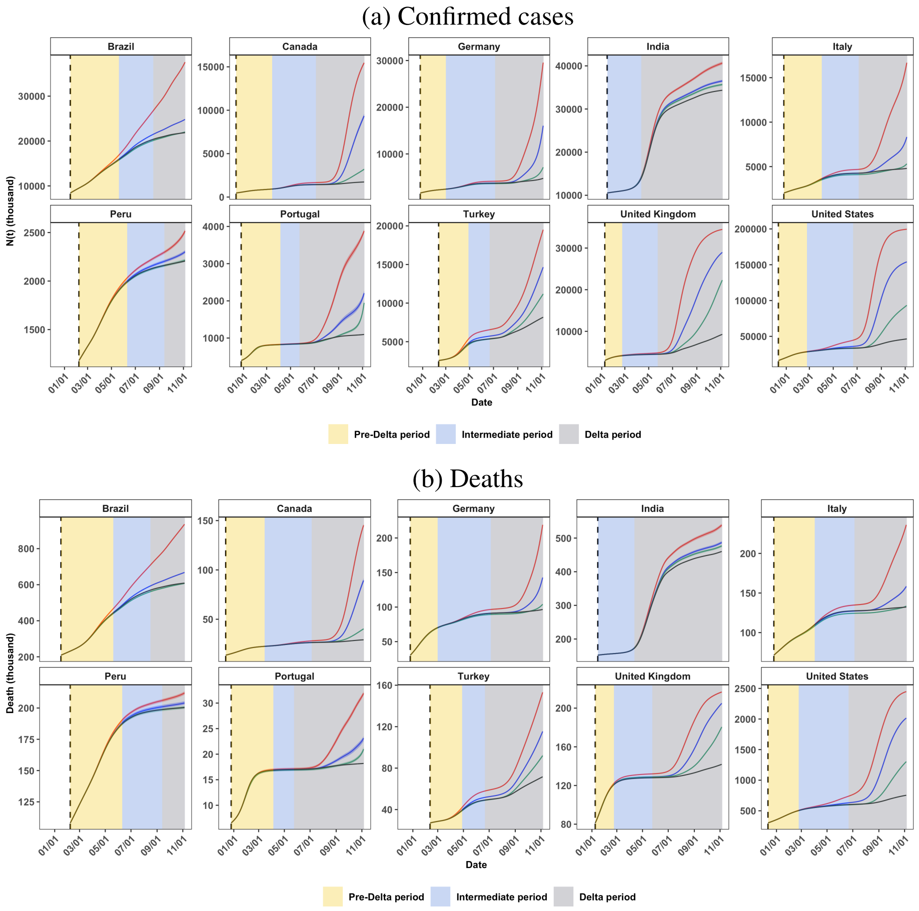


Fig S14: The actual (black), and the would-be total numbers under the no vaccination (red), the partial vaccination (blue) and the first-dose-priority (green) vaccination scenarios of (a) confirmed cases and (b) deaths. The 2.5 - 97.5% quantiles of the would-be total numbers are indicated by colored areas. The yellow, light blue and gray colored areas mark the pre-Delta, intervening and Delta periods, respectively. The black dashed vertical line represents the start of the vaccination.

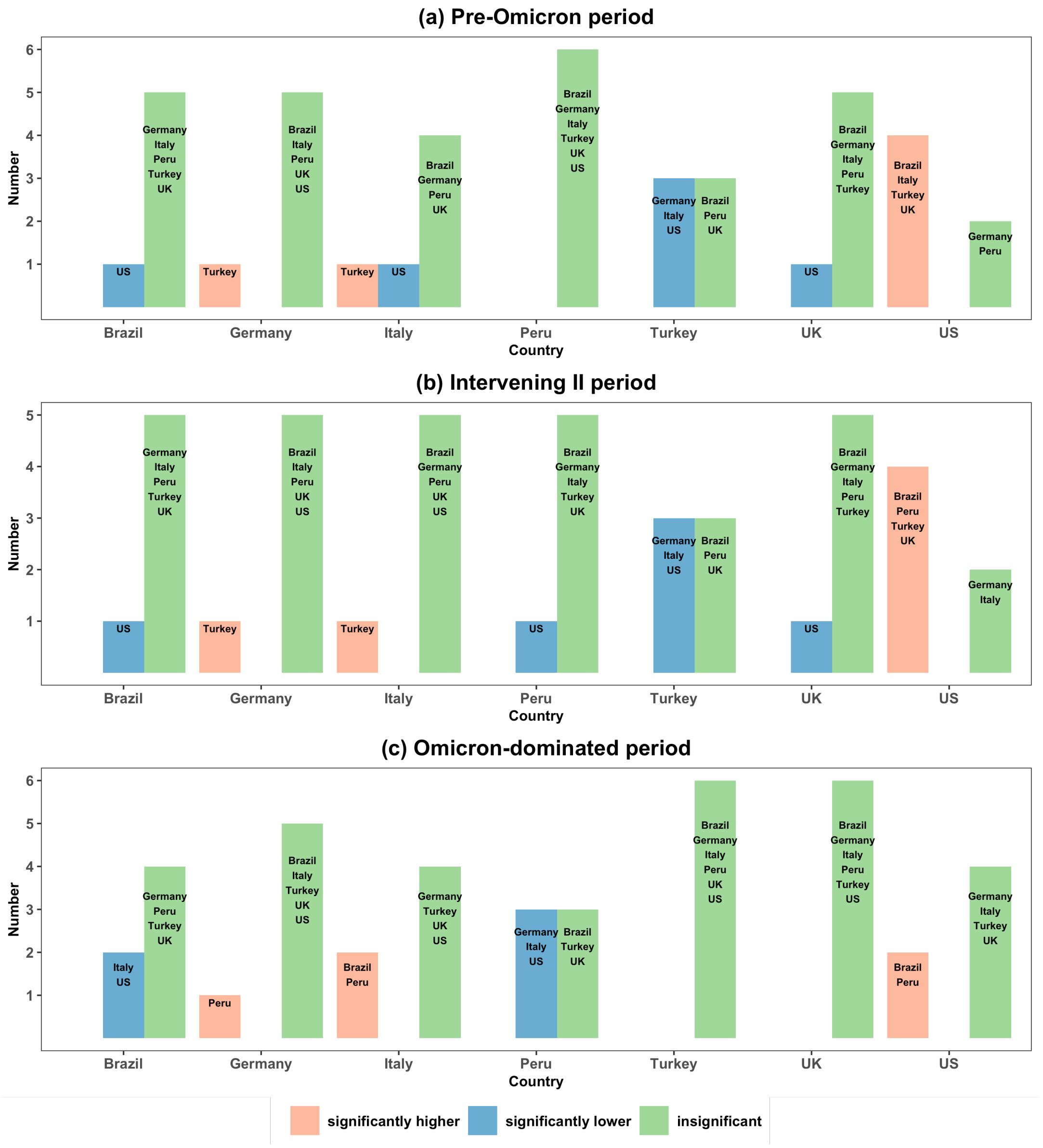


Fig S15: Frequency bar-charts on the pairwise VPR comparison of the country on the horizontal coordinate whose VPR of the partial vaccination was significantly higher than (red), significantly lower than (blue) or insignificantly different from (green) the other countries by conducting the pairwise testing on the VPRs via the bootstrap method among seven countries in the pre-Omicron (a), intervening II (b) and Omicron-dominated (c) periods.

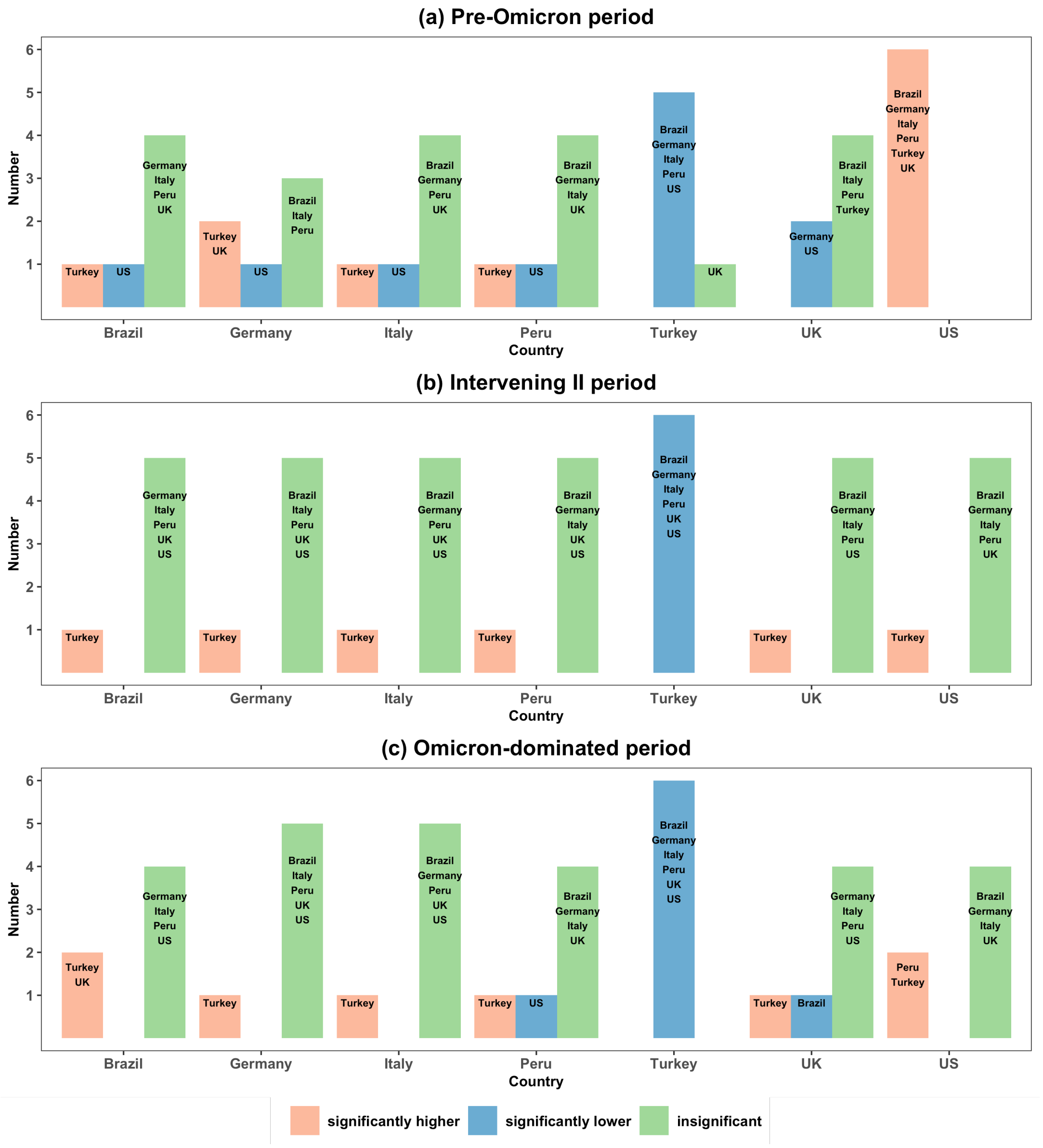


Fig S16: Frequency bar-charts on the pairwise VPR comparison of the country on the horizontal coordinate whose VPR of the full vaccination was significantly higher than (red), significantly lower than (blue) or insignificantly different from (green) the other countries by conducting the pairwise testing on the VPRs via the bootstrap method among seven countries in the pre-Omicron (a), intervening II (b) and Omicron-dominated (c) periods.

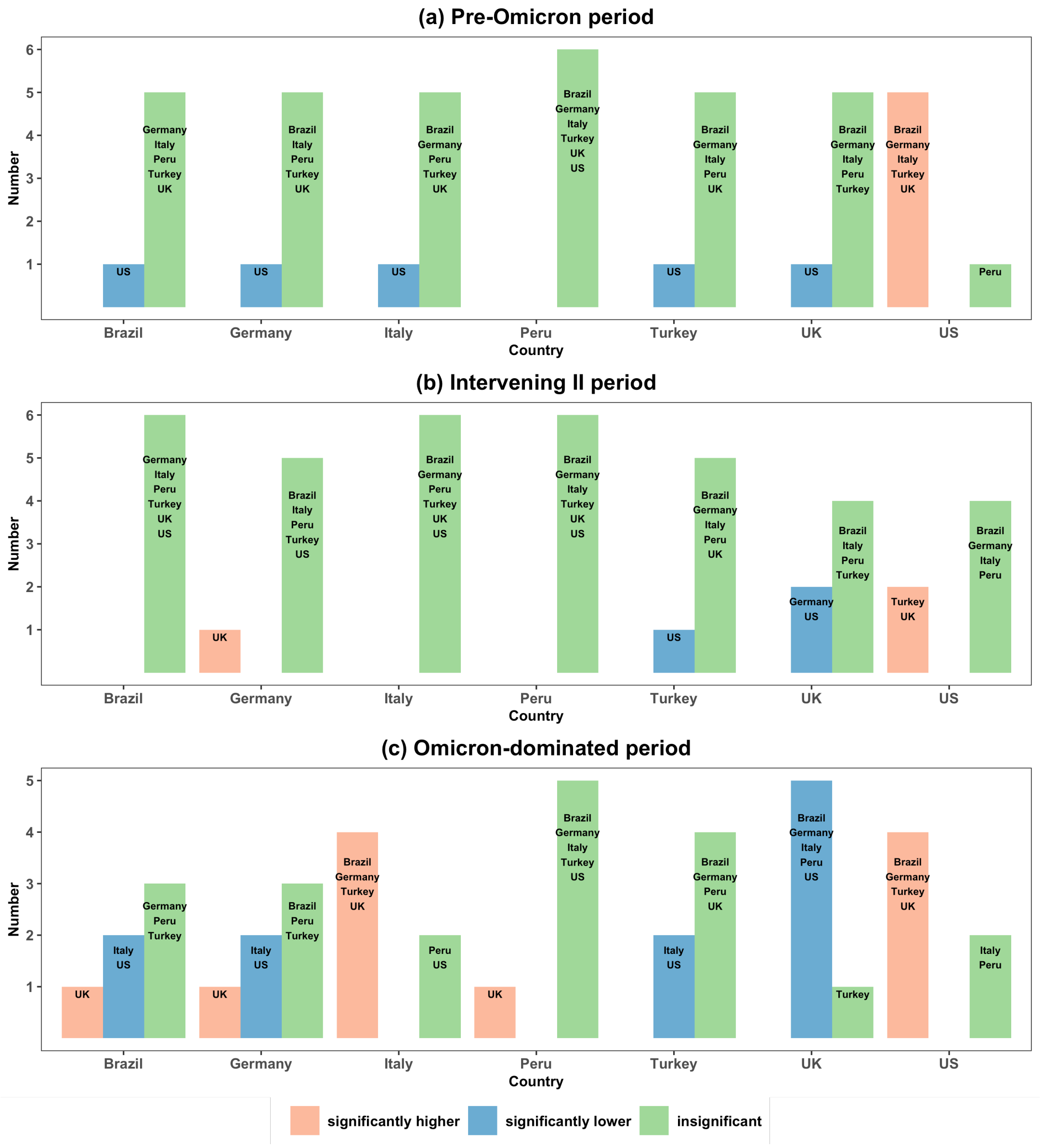


Fig S17: Frequency bar-charts on the pairwise VPR comparison of the country on the horizontal coordinate whose VPR of the booster vaccination was significantly higher than (red), significantly lower than (blue) or insignificantly different from (green) the other countries by conducting the pairwise testing on the VPRs via the bootstrap method among seven countries in the pre-Omicron (a), intervening II (b) and Omicron-dominated (c) periods.
